# Correlates of Protection and Response Durability of the RTS,S Vaccine in Four Malaria Challenge Studies

**DOI:** 10.64898/2026.09.30.26364316

**Authors:** Soo-Young Kim, Thomas Linnekin, Lindsay N. Carpp, Cynthia K. Lee, Christian F. Ockenhouse, Laina D. Mercer, Scott Gregory, Caitlyn Linde, Todd Suscovich, Tom Shneer, Kelly E. Seaton, S. Moses Dennison, Rachel L. Spreng, Sarah V. Mudrak, Laura Fontana, Drienna Holman, Solmaz Shotorbani, Jishnu Das, James E. Moon, Neville Kisalu, Lenny Moise, Galit Alter, Georgia D. Tomaras, Ollivier Hyrien

## Abstract

RTS,S/AS01 (RTS,S) is one of two WHO-recommended malaria vaccines for children in moderate-to-high transmission settings. Yet, no validated immune correlate of protection (CoP) has been established for this vaccine. We selected 16 biomarkers to further assess as candidate CoPs, based largely on their ability to well-predict post-challenge outcomes in a previous integrative correlates analysis of two phase 2a controlled human malaria infection (CHMI) studies, Malaria-068 and Malaria-071. We evaluated the predictive power of these 16 biomarkers in two additional phase 2a CHMI studies, Malaria-092 and Malaria-102, to confirm these candidate CoPs in new malaria vaccine trials. In MAL092, malaria-naïve volunteers were challenged three months after a series of two or three doses of RTS,S/AS01, and in MAL102, a subset were boosted nine months later and re-challenged after three weeks. The 16 biomarkers included *Plasmodium falciparum* circumsporozoite protein (CSP)-specific IgG antibodies targeting the CSP NANP-repeat region (CSP IgG ELISA), NANP repeat-specific antibody binding to human Fc gamma receptors (FcγRs), CSP-specific antibody Fc effector functions, total NANP6-specific serum Ig measured by biolayer interferometry (BLI), and NANP6-specific IgG1 antibodies measured by binding antibody multiplex assay. Immunogenicity analyses confirmed that the vaccine regimens elicited robust immune responses, including multiple biomarkers not previously reported on, in both trials. In MAL092, participants clustered into two distinct immunogenicity profiles that overlapped with malaria protection status. In univariate logistic regression analyses, most biomarkers were significantly associated with protection when measured on the day of challenge in both trials. Notably, the same three biomarkers ranked as top correlates in both trials (FCGR2AH NANP6, FCGR2AR NANP6, and FCGR2B NANP6; all adjusted p-values <0.001), with similar effect sizes across studies. Despite these strong associations, no biomarker demonstrated significantly better individual-level predictive performance than CSP IgG ELISA. In MAL092, CSP IgG ELISA achieved an area under the curve (AUC) [95% confidence interval (CI)] of 0.74 (0.63, 0.84), compared with AUCs of 0.76 - 0.78 for the top five biomarkers, all of which reflected NANP6-specific FcγR binding (all adjusted p-values > 0.37 versus CSP IgG ELISA). Predictive models trained on data from two prior CHMI studies and applied to MAL092 and MAL102 identified a model based on FCGR2B NANP6 (validation AUC = 0.77 in MAL092; 0.80 in MAL102) as the best-performing univariate model for both studies. The best-performing multivariate predictive model for MAL092 was based on BLI NANP6 Dissociation AUC and ADCP NANP6 (validation AUC = 0.76), and in MAL102, it was based on CSP IgG ELISA, BLI NANP6 Dissociation AUC, and ADCP NANP6 (validation AUC = 0.82). The next approximately 10 highest-ranked models, however, had comparable performance with overlapping 95% CIs, and none were clearly superior to CSP IgG ELISA (validation AUCs 0.74 and 0.71 in MAL092 and MAL102, respectively). Collectively, these findings suggest that the majority of the previously identified candidate CoPs also hold up well in two additional malaria vaccine CHMI trials and underscore the continued utility of the CSP IgG ELISA readout as a robust marker of protection. Further studies are needed to determine whether these candidate CoPs hold in malaria-endemic field settings. Identifying validated CoPs that predict malaria vaccine efficacy would promote the development and approval of more effective and durable next-generation vaccines. These advances could also inform vaccine strategies for other infectious diseases.

## Introduction

Malaria remains a formidable global health challenge, responsible for an estimated 263 million cases and nearly 600,000 deaths annually (1). It disproportionately affects people living in regions with high transmission rates of *Plasmodium* parasites (the causative agents of malaria), particularly sub-Saharan Africa, where children under the age of five are particularly vulnerable to severe disease due to their underdeveloped immunity (2, 3). Adults are also affected, contributing to poverty, lost productivity, and strain on health care systems (4–6). While vector control strategies such as insecticide-treated bed nets and indoor residual spraying can significantly reduce transmission (7), additional tools are urgently needed. Vaccines that are safe, efficacious, and durable in children and adults offer a long-term, cost-effective strategy to complement existing interventions to curtail transmission across entire communities (8, 9).

Significant progress has been made in the development of vaccines targeting pre-erythrocytic *Plasmodium falciparum* (*P. falciparum*) parasites (10, 11). Two such vaccines — RTS,S/AS01 (RTS,S) and R21/Matrix-M (R21) — have demonstrated protective efficacy against clinical malaria, and have both been recommended by the World Health Organization (WHO) for the prevention of *P. falciparum* malaria in children living in malaria-endemic areas (12). RTS,S is a recombinant protein vaccine targeting the central Asn-Ala-Asn-Pro (NANP) repeat and C-terminal regions of the *P. falciparum* circumsporozoite (CSP) protein and adjuvanted with AS01 (13–15). CSP-targeting antibody responses elicited by RTS,S vaccination have been demonstrated to block sporozoite invasion of hepatocytes (16), thereby reducing progression to blood-stage infection. WHO’s recommendation of RTS,S was based, in part, on the results of a pivotal randomized, controlled phase 3 trial in African infants and children (17–20), in which vaccine efficacy (VE) of three doses of RTS,S/AS01_E_ against clinical malaria over 12 months was estimated to be 56% in young children (five to 17 months old) (17) and 31% in infants (six to 12 weeks old) in areas of low, medium, and high transmission (18). The second vaccine, R21, also targets the sporozoite stage and includes the NANP repeat and C-terminal CSP regions (21). It was the second malaria vaccine to be recommended by the WHO (22). In a randomized phase 3 trial, estimated VE of three doses of R21/Matrix-M against clinical malaria was 68% (standard administration) over 12 months in children (five to 36 months old) in areas of low to medium transmission (23). As RTS,S and R21 have not been tested in a head-to-head trial, neither vaccine has been shown to have superior efficacy to the other, and the WHO has recommended that the choice between these two vaccines should be based on factors such as supply and affordability (24).

Both vaccines have the potential for high public health impact (25); however, neither attains the WHO preferred product characteristics for an optimal malaria vaccine in terms of efficacy and durability — a 90% reduction in blood stage infection and clinical malaria over 12 months (26, 27). Therefore, key challenges remain in achieving higher and more durable efficacy, particularly in diverse transmission settings and across age groups (9). Booster doses of RTS,S have been evaluated to address waning immunity, yet results have been mixed regarding their ability to sustain protective antibody levels (28–30). Moreover, a longitudinal qualitative evaluation study for the malaria vaccine implementation program reported strong uptake of three doses of RTS,S, yet a notable drop in adherence for the fourth dose, including among caregivers motivated to bring their children for vaccination (31). Thus, understanding and improving the longevity of antibody responses is an important goal of malaria vaccine research, with the potential to inform the development of next-generation malaria vaccines with improved immunogenicity and long-term protection (26).

Accelerating vaccine development and understanding vaccine-induced protective immunity relies on robust immune correlates of protection (CoPs), immune biomarkers whose levels reliably predict the efficacy of a vaccine against a given clinical endpoint (32–34). Immune CoPs for malaria vaccines (35–37) have been evaluated across a range of settings. These include controlled human malaria infection (CHMI) studies of malaria-naïve individuals exposed through mosquito bites (38), more recent CHMI studies of adults from endemic regions using injected sporozoites or infected red blood cells (39, 40), and field trials conducted primarily in infants and children in malaria-endemic areas (38). Although findings from a CHMI study of malaria-naïve adults may not always translate to children in a malaria-endemic area, the approach offers critical advantages. CHMI studies enable knowledge of immune biomarker levels precisely at the time of exposure, require fewer participants and small sample volumes, and may be completed faster than field studies. For these reasons, CHMI studies are an invaluable tool for identifying candidate CoPs that can then be prioritized for validation in the target population.

The present work focuses on the Malaria-092 and Malaria-102 CHMI studies (hereafter referred to as MAL092 and MAL102) and builds upon results from the earlier Malaria-068 and Malaria-071 (hereafter referred to as MAL068 and MAL071) studies. MAL068 [NCT01366534 (41)] and MAL071 [NCT01857869 (42)] were randomized, phase 2a trials in healthy, malaria-naïve adults that evaluated standard (RRR: two full doses one month apart) and modified (ARR: an alternative arm with a modified version of the RRR) RTS,S regimens. MAL071 demonstrated high efficacy using a delayed fractional third dose regimen (RRr: two full doses one month apart, followed by a fractional fifth dose six months later). In both studies, participants were challenged with *P. falciparum* sporozoites via the bite of five infected mosquitoes three weeks after the last vaccination.

MAL092 [NCT03162614 (43)] was a phase 2a, open-label, single-center clinical study designed to evaluate the safety, immunogenicity, and efficacy of RTS,S/AS01 administered using different dosing schedules in healthy malaria-naïve adults aged 18–55 years. The study included six groups: four received vaccinations at months 0, 1, and 7; one followed a 0 and 7-month schedule, and the sixth served as an infectivity control. Doses were either full or fractional (Fx) of either the adult (Adu) or pediatric (Ped) RTS,S formulation, with antigen quantity, adjuvant quantity, and volumes of each dose summarized in Supplementary Table 1A. All vaccinated participants underwent a controlled human malaria infection (CHMI) (43), but unlike the two earlier studies, the *P. falciparum* sporozoite challenge occurred three months after the last vaccine dose.

Finally, MAL102 [NCT03824236 (44)] was a follow-up booster study to MAL092, aimed at assessing the durability of protection observed with fractional dosing schedules and determining whether an additional fractional booster dose can extend protection. In this study, a subset of vaccinated participants from MAL092, regardless of their initial protection status following MAL092 CHMI, received a booster vaccination (Supplementary Table 1B) and underwent a second CHMI three to four weeks later to evaluate waning efficacy and the potential for extended protection.

Multiple studies have previously assessed immune response biomarkers as candidate CoPs in MAL068 and/or MAL071 (41, 45–53). A comprehensive assessment by Young et al. used the full repertoire of day-of-challenge immune marker data in these two studies, including antibody, cellular, and transcriptomic immune responses. In cross-study validation analyses, the level of NANP6-targeted antibody dependent cellular phagocytosis (ADCP NANP6); the levels of NANP6-specific antibody engagement of different human Fc gamma receptors (FcγRs; the present work uses “FCGR” in the biomarker nomenclature, where FCGR2 is the same as FcγRII and FCGR3 is the same as FcγRIII), particularly FCGR3A; and anti-NANP6 IgG1 antibody levels were consistently among the most predictive univariate and bivariate models (51), although biomarker ranking varied according to the specific analysis. The standard NANP-based CSP IgG ELISA readout (Pfal CSP R32LR IgG, based on binding to a CSP-derived antigen consisting of 32 NANP repeat units with a leucine-arginine tail), which has been used for more than 30 years in malaria vaccine development (54, 55), also ranked highly in some of the analyses. Another general observation was that there was no improvement in cross-study prediction using bi- or trivariate models over the best univariate model.

The primary MAL092 paper reported antibody responses and avidity indices to CSP (full-length, repeat region, C-terminal region) as measured by ELISA-based assays, as well as antibody levels to the hepatitis B surface antigen component of the vaccine (anti-HBs), from Day 1 (prevaccination) through Day 377 (43). The only antibody response reported to be significantly different between protected and unprotected participants was anti-CSP (repeat region) concentration (ELISA units/ml), with levels significantly higher in protected participants from 1 month after the final vaccination (Day 227) until the end of the study (Day 377). The primary MAL102 paper also reported antibody responses to CSP (full-length, repeat, C-terminal region) as measured by ELISA [endpoint titers (EU/ml)] and anti-HBs antibody concentrations from Day 1 (prevaccination) through Day 190 (44). All assessed CSP-specific antibody responses had comparable magnitudes between protected and non-protected participants, with no antibody response reported to be significantly different between the two groups. More recent work by Spreng et al. extended this analysis by incorporating MAL092 and MAL102 data obtained using a subset of the immune assays used in Young et al., biolayer interferometry (BLI) and binding antibody multiplex assay (BAMA) (56). A key finding from their multivariate analyses combining data across MAL068, MAL071, and MAL092 was that the most predictive multivariate model included three immune markers: serum NANP6-specific immunoglobulins (Ig) binding and avidity composite, serum N Interface Ig binding and avidity composite, and serum NANP6-specific immunoglobulin G1 (IgG1) binding measurements (56). These findings motivate the refinement of predictive models using the complete set of bioassays implemented in MAL092 and MAL102.

The present work builds upon and extends the work of Spreng et al. by focusing on a set of 16 candidate CoPs, selected based largely on their ability to well-predict post-challenge outcomes in MAL068 and MAL071 in the previous integrative correlates analysis (51). The main objective was to comprehensively further assess these candidate CoPs through immune correlates analyses of MAL092 and MAL102. This was first addressed through univariate analyses to assess the predictive power of previously identified MAL068 and MAL071 candidate CoPs in MAL092 and MAL102. In addition, multivariate analyses were conducted to refine predictive models by combining biomarkers and to identify optimal subsets by evaluating all possible pairs, triplets, and the full set of 16 biomarkers for improved prediction of protection status. To support interpretation of the correlates results, we characterized vaccine-induced immune responses by comparing pre- and post-vaccination measurements, and evaluated response durability in terms of fold-change from peak immunogenicity to the day of challenge, in MAL092 and MAL102. We also evaluated coregulation of immune responses and identified potential redundancy among the 16 readouts.

## Results

### Rich immunogenicity data from 16 biomarkers across four CHMI studies

This study considers immunogenicity data from 94 participants in MAL092 and 36 participants in MAL102. The present analysis considers 16 biomarkers that were among the highest-ranking (out of the 275 and 338 down-selected biomarkers in the MAL068 and MAL071 datasets, respectively) in predicting post-challenge outcomes within and/or across studies in an integrative correlates analysis conducted in the MAL068 and MAL071 CHMI studies (51).

Additional details on the 16 biomarkers are provided in Supplementary Table 2, and Supplementary Table 3 summarizes their predictive ability across the different analyses considered in the integrative work (51) as rationale for their inclusion in the present work. For MAL092 and MAL102, 12 of the 16 markers were not assessed in previous papers (43, 44, 56), consisting of Fc effector-related readouts (n = 4), FCGR specificity of NANP6-targeting antibodies (n = 6), an additional measurement of total NANP6-targeting IgG antibodies (Total IgG NANP6) (n=1), and an enzyme-linked immunosorbent assay (ELISA)-based measurement of IgG antibodies directed against the CSP NANP6 repeat region (hereafter, “CSP IgG ELISA”) (n=1).

### Protection from the original three-dose series did not influence protection upon re-vaccination

Vaccine efficacy (VE) estimates in all four trials are reported in Supplementary Table 4. The table also provides protection efficacy (PE) estimates against infection (*P. falciparum* parasitemia as assessed by blood smear after sporozoite challenge), across the participants from the four CHMI trials (MAL068, 071, 092, and 102) who contributed immunogenicity data to our analyses, stratified by study and vaccination arm. PE is defined here as the proportion of vaccinated participants who were not infected following challenge (details in Supplementary Table 4). This estimand differs from the VE estimates reported in the primary trial papers, both in definition and in the fact that the PE estimates were calculated only participants who consented to have their samples used in subsequent investigations, a subset of the per-protocol sets used in the primary trial papers. PE directly characterizes protection among vaccinated individuals and reflects the amount of immunogenicity data available for analysis. In this controlled challenge setting, PE provides a stable and interpretable estimand for relating immune markers to protection, while VE is reported for completeness and comparability with the primary trial results.

When pooled across arms, PE estimates were 47.8% in MAL068 (ARR and RRR arms), 77.8% in MAL071 (RRR and RRr arms), 56.4% in MAL092 (five vaccinated arms), and 52.8% in MAL102. PE estimates did not differ significantly between study arms across trials (all p-values from chi-squared test), including ARR and RRR in MAL068 (p = 0.57), MAL071 (p = 0.067), MAL092 (p = 0.064), and across the five MAL102 arms corresponding to original MAL092 assignments (p = 0.62).

We found no evidence that protection status in MAL092 was related to protection status in MAL102. Of the 35 participants from MAL092 who subsequently enrolled in MAL102 and consented to have their samples used in subsequent investigations, n = 18 (51.4%) were protected and n = 17 (48.6%) were not protected against malaria challenge in MAL092. Following re-challenge in MAL102, 17 participants (48.6%) were not protected and 18 (51.4%) were protected. Supplementary Figure 1 visualizes protection status in MAL092 vs. MAL102, confirming no apparent relationship. Estimated PE in MAL102 did not significantly differ from that in MAL092 (McNemar’s test, p = 1.0; Cramer’s V = 0.029). Moreover, protection status in MAL092 was not significantly associated with protection status in MAL102 (PE estimates in MAL102: among MAL092-protected participants: 50%, among MAL092-not protected participants, 52.9%; chi-squared test, p = 1.0). These results suggest that protection following vaccination and/or challenge in MAL092 did not measurably contribute to protection following re-vaccination in MAL102, a conclusion in agreement with the primary MAL102 paper that performed the same analysis with the entire per-protocol set (44).

### Further evidence that the MAL092 and MAL102 vaccine regimens were immunogenic

Immunogenicity has been extensively characterized for MAL068 and MAL071 (41, 42, 45–47, 49, 50, 52, 53), and thus we do not focus on this here. Longitudinal trajectories for all 16 biomarkers in MAL092 and MAL102 are shown in Figure 1. Comparison of baseline (D0) to peak immunogenicity (Day 286; i.e., the day of challenge) indicated that all five MAL092 regimens were immunogenic, with significant increase in 15 of the 16 biomarkers [Wilcoxon signed-rank test; all adjusted p < 0.10; IgG1 CSP avidity index (AI) was not measured at D0 and was therefore excluded from these comparisons]. Supplementary Figure 2 shows individual-level antibody trajectories from Day 226 onward (through Day 376 for the first five biomarkers shown, and through Day 286 for the others) for MAL092 participants, where the markers exhibited log-linear decay.

**Figure 1.**
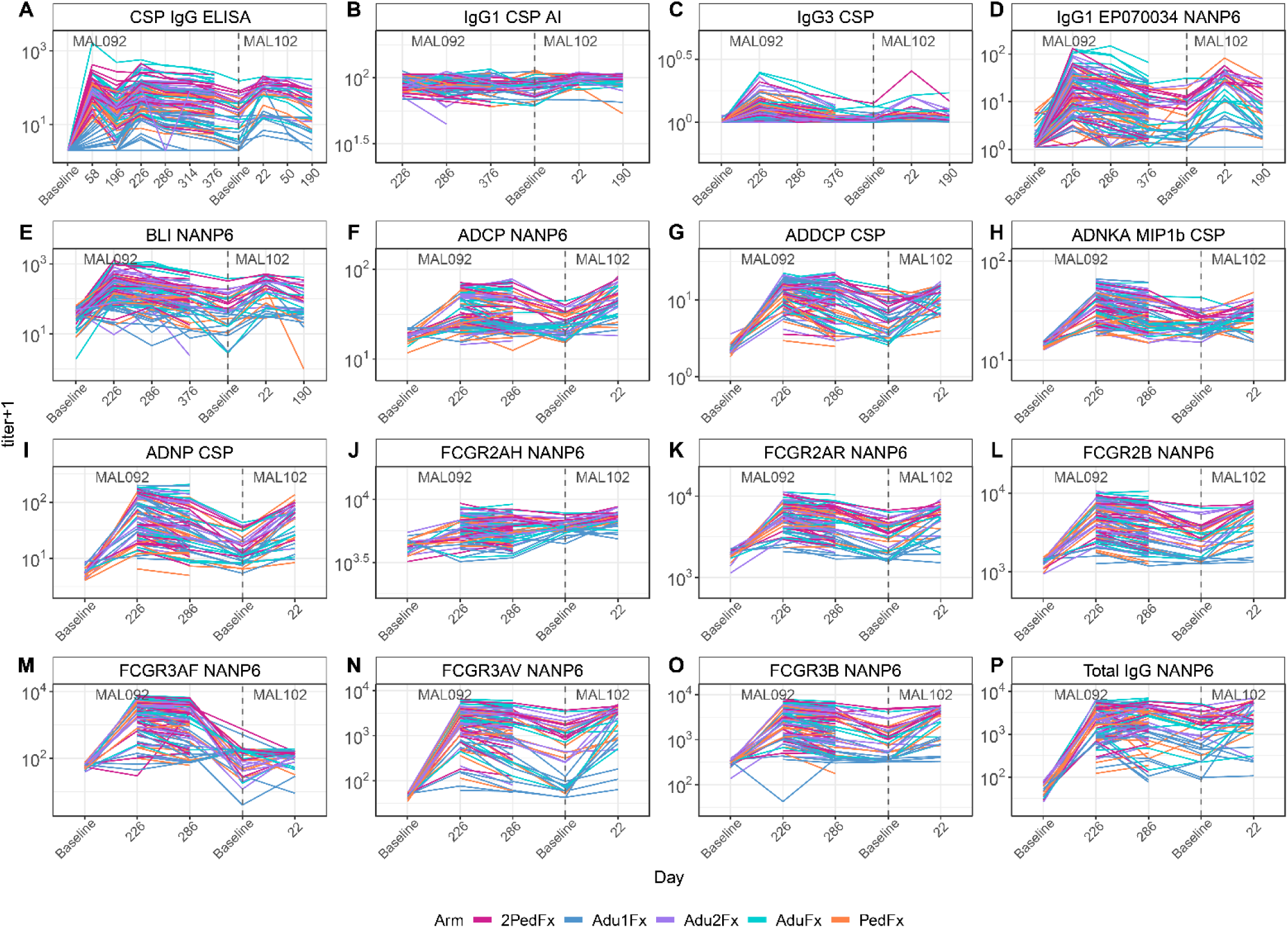
Longitudinal levels in MAL092 and in MAL102 of the 16 biomarkers considered in this study. Biomarker levels are plotted over time (each line = one participant) and colored by enrollment arm, and the x-axis indicates the study visit (in Day post baseline in MAL092 or MAL102, where baseline for MAL102 is designated with a vertical dashed line). Values were missing for FCGR3B NANP6 and ADCP NANP6, where only 22 observations were available at Day 0 but 94 observations were available at Days 226 and 286. Each dot represents one study participant. The y-axis of each plot represents the measured titer (log10-transformed after having added 1). ADCP, antibody-dependent cellular phagocytosis; ADDCP, antibody dependent dendritic cell phagocytosis; ADNKA, antibody-dependent natural killer cell activation; ADNP, antibody-dependent neutrophil phagocytosis; Adu, Adult; AI, avidity index; BLI, bio-layer interferometry; CSP, circumsporozoite protein; ELISA, enzyme-linked immunosorbent assay; Fx, fractional; IgG1, immunoglobulin G1; FCGR, Fc gamma receptor; IgG3, immunoglobulin G3; Ped, pediatric. Biomarker nomenclature: BLI NANP6: BLI NANP6 Dissociation AUC. ELISA: CSP IgG ELISA.

Focusing on the MAL092 day of challenge, Figure 2 shows distributions of the 16 biomarkers by MAL092 study arm (AduFx = full adult doses at Months 0 and 1 and 1/5^th^ of an adult dose at Month 7; Adu1Fx = full adult dose at Month 0 and 1/5^th^ of an adult dose at Month 7; Adu2Fx = full adult dose at Month 0 and 1/5^th^ adult doses at Months 1 and 7; PedFx = pediatric doses at Months 0 and 1 and 1/5^th^ of a pediatric dose at Month 7; and 2PedFx = double pediatric doses at Months 0 and 1 and a double 1/5^th^ pediatric dose at Month 7; details of each study arm are provided in Supplementary Table 1).

**Figure 2.**
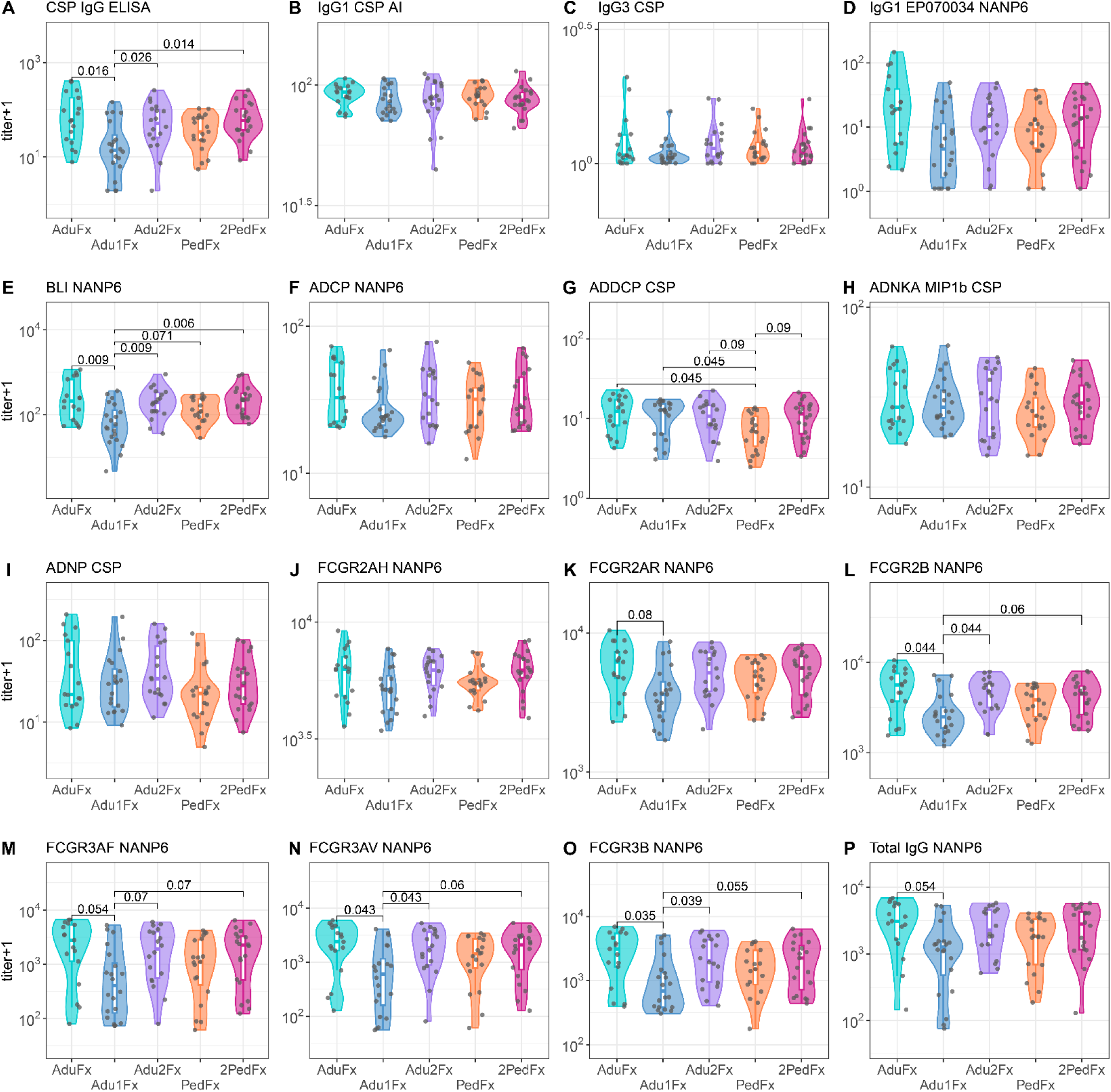
Distributions of MAL092 day-of-challenge levels for the 16 biomarkers considered in this study, plotted by MAL092 study arm. Each dot represents one study participant. In each plot, adjusted p-values from pairwise group comparisons (Wilcoxon rank-sum test) are included in each plot when they were smaller than 0.10. P-value adjustment was carried out separately for each biomarker using the Benjamini-Hochberg procedure. ADCP, antibody-dependent cellular phagocytosis; ADDCP, antibody dependent dendritic cell phagocytosis; ADNKA, antibody-dependent natural killer cell activation; ADNP, antibody-dependent neutrophil phagocytosis; Adu, Adult; AI, avidity index; BLI, bio-layer interferometry; CSP, circumsporozoite protein; ELISA, enzyme-linked immunosorbent assay; Fx, fractional; IgG1, immunoglobulin G1; FCGR, Fc gamma receptor; IgG3, immunoglobulin G3; Ped, pediatric. Biomarker nomenclature: BLI NANP6: BLI NANP6 Dissociation AUC. ELISA: CSP IgG ELISA.

With statistical significance defined as an adjusted p-value from pairwise group comparisons < 0.10 (see Methods), biomarkers tended to fall into one of two distinct patterns: 1) significantly lower in the Adu1Fx arm than in the AduFx, Adu2Fx, and 2PedFx arms (5/16 biomarkers); or 2) no significant differences across arms (7/16 biomarkers). Biomarkers showing pattern #1 included CSP IgG ELISA, binding of NANP6-specific antibodies to the FCGR2B receptor (FCGR2B NANP6), the FCGR3A F158 variant (FCGR3AF NANP6), the FCGR3A V158 variant (FCGR3AV NANP6), and the FCGR3B receptor (FCGR3B NANP6) (all adjusted p ≤ 0.055; Wilcoxon rank-sum test) (Figure 2A, 2L, 2M, 2N, 2O). For BLI NANP6, levels were lower in the Adu1Fx arm compared to all other arms (all adjusted p ≤ 0.071; Wilcoxon rank-sum test) (Figure 2E). Binding of NANP6-specific antibodies to the FCGR2A R131 variant (FCGR2AR) and total IgG antibodies to NANP6 (Total IgG NANP6) were only significantly lower in the Adu1Fx vs. AduFx arms (adjusted p = 0.08 and 0.054, respectively, Wilcoxon rank-sum test) (Figure 2K, 2P). In contrast, CSP-specific antibody-dependent dendritic cell phagocytosis (ADDCP CSP) was significantly lower in the PedFx arm compared to all other arms (all adjusted p ≤ 0.09, Wilcoxon rank-sum test) (Figure 2G). No other biomarkers differed significantly between any of the MAL092 arms on the day of challenge (pattern #2: Figure 2B, 2C, 2D, 2F, 2H, 2I, 2J).

The MAL102 vaccine regimen was also immunogenic, irrespective of MAL092 arm assignment. Comparing MAL102 measurements across study visits (MAL102 baseline versus Day 22), immune responses were detected for 15 of 16 biomarkers (Wilcoxon signed-rank test; all adjusted p < 0.05), with the exception of FCGR3AF NANP6 (adjusted p = 0.41). Supplementary Figure 3 shows day-of-challenge distributions for the 16 biomarkers in MAL102, by MAL092 study arm. Overall, responses were highest in the 2PedFx arm and lowest in the Adu1Fx arm, although between arm differences reached statistical significance for only two markers, FCGR2AR NANP6, FCGR2B NANP6 and FCGRAV NANP6 (adjusted p = 0.048, 0.048 and 0.076, respectively, Wilcoxon rank-sum test)]. Exceptions to this trend were observed for ADDCP CSP and CSP-specific antibody-dependent natural killer cell activation (ADNKA MIP1b CSP).

### Multivariate analysis reveals two immune response profiles in MAL092 participants

To identify patterns in the heterogeneous immune responses measured by the 16 day-of-challenge biomarkers in MAL092, dimensionality reduction was performed using *t*-distributed Stochastic Neighbor Embedding (*t*-SNE) (57). The analysis revealed that vaccine recipients generally grouped into three primary immune response profiles (Supplementary Figure 4). One group included participants with consistently high biomarker levels (upper right, magenta cluster); another included participants with consistently low biomarker levels (lower left, blue cluster); and the third included participants with intermediate and heterogeneous responses, showing relatively higher or lower levels depending on the specific biomarker (central region).

Overlaying protection status onto the *t*-SNE map suggested that participants protected against challenge in MAL092 tended to localize in the upper right region of the map (high immune responses), whereas non-protected participants tended to cluster in the lower left region (low immune responses) (Figure 3A). Protected and non-protected participants were significantly differently distributed (p = 0.001). These patterns suggest that some of the 16 biomarkers were associated with protection status. Similarly, projection of study arm onto the *t*-SNE map indicated differences in immune response profiles between vaccine regimens (Figure 3B). For example, participants in the Adu1Fx arm tended to cluster in the lower left region, whereas participants in the 2PedFx arm tended to cluster in the upper right region, consistent with the immunogenicity differences observed in Supplementary Figure 3. The observation of different vaccine regimens appearing to have different immune response profiles was confirmed, with at least one study arm group differing significantly from the others (p = 0.012).

**Figure 3.**
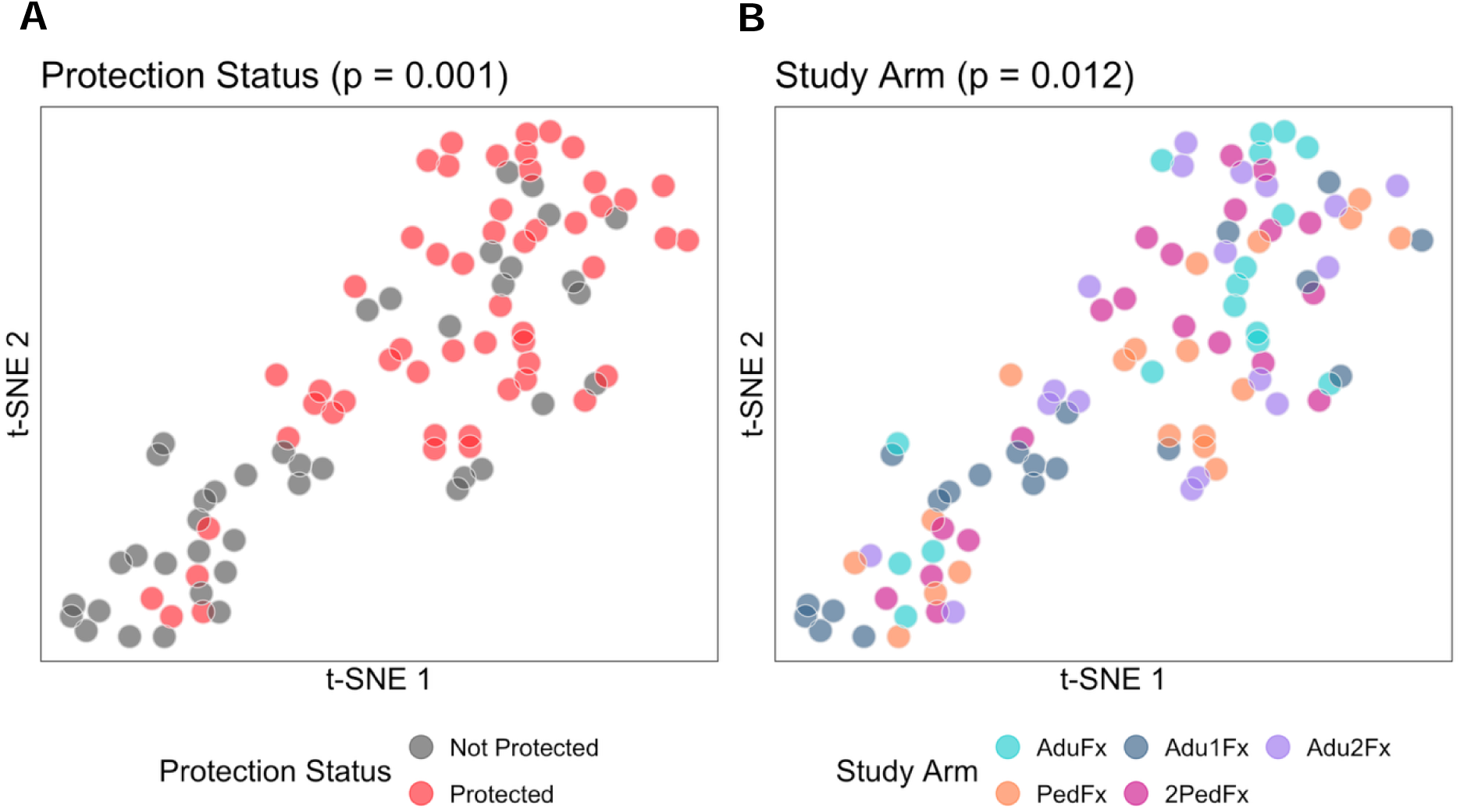
t-SNE map built using the day-of-challenge levels of all 16 biomarkers measured in MAL092. Each dot represents one study participant, colored by (A) protection status and (B) treatment arm. P-values are from testing whether protected and non-protected participants had significantly different distributions (left panel) and from testing whether at least one study arm group differed significantly from the others (right panel). Adu, Adult; Fx, fractional Ped, pediatric; t-SNE, t-distributed Stochastic Neighbor Embedding.

### High correlations within and between peak immunogenicity and day-of-challenge responses in MAL092

We next evaluated coordination of immune response components captured by the 16 biomarkers, assessed whether/how this coordination changed over time, and determined whether/how this coordination was linked with protection status. Supplementary Figure 5 shows Spearman’s rank correlation coefficients between day-of-challenge biomarkers for each of the four CHMI trials, stratified by protection status (all, protected, and non-protected).

The correlation matrices showed generally strong associations between biomarkers measured in MAL092, particularly within assay (third column, Supplementary Figure 5). When including all participants regardless of protection status, the CSP IgG ELISA readout in MAL092 correlated highly with BLI NANP6 Dissociation AUC (*ρ* = 0.93) and with the FCGR2AR/B and FCGR3A/B readouts (0.85 < *ρ* < 0.91). By contrast, the IgG1 CSP avidity index and IgG3 CSP concentration were weakly correlated with all other biomarkers, and most correlations were not significant after adjustment (adjusted p > 0.05).

Overall, the day-of-challenge correlation structure among the 16 biomarkers was consistent across MAL068, MAL071, MAL092, and MAL102 (Supplementary Figure 5), thereby suggesting that immune responses were similar across studies. A notable exception in MAL102 was FCGR3AF NANP6, which correlated with most other biomarkers in MAL092 (third column) but showed little to no correlation with these biomarkers in MAL102 (fourth column) (Supplementary Figure 5).

While antibody levels may be higher in protected individuals than in non-protected individuals, the overall pattern of day of challenge correlations was similar when stratified by protection status (Supplementary Figure 5), indicating that the global coordination of immune responses was largely preserved in both protected and non protected participants. FCGR3AF NANP6 was a potential exception in MAL102, where the correlation profiles of this biomarker appeared to differ between protected and non-protected participants, with correlations absent from non-protected participants but present among protected participants.

The correlations within readouts across peak immunogenicity (Day 226) and the day of challenge (Day 286) in MAL092 were generally high (Figure 4A). The overall correlation structures were similar at both time points, with correlation coefficients tending to be slightly higher at peak immunogenicity. Supplementary Figure 6 shows correlations of day-of-challenge (Day 286) readouts in MAL092 versus day-of-challenge (Day 22) readouts in MAL102, where most pairs of biomarkers were positively correlated, but some pairs showed no correlation or inverse correlations. Supplementary Figure 7 shows scatterplots for individual readouts (MAL092 day-of-challenge vs. MAL102 day-of-challenge), where 10 out of the 16 biomarkers had *ρ* ≥ 0.59 and p < 0.001, and 13 of the 16 biomarkers had *ρ* ≥ 0.38 and p < 0.05. The scatterplots suggest that the immune responses were consistently stronger in MAL102 as compared to MAL092, as evidenced by the majority of the points falling above the line of identity for all 5 of the 6 readouts (FCGR3AF NANP6 being the exception). Cumulatively, these results suggest that biomarkers measured at peak immunogenicity could serve as early predictors of individual-level protection status, consistent with one of the approaches used in some correlates analyses of vaccine efficacy trials for various infectious diseases (33, 58–61).

**Figure 4.**
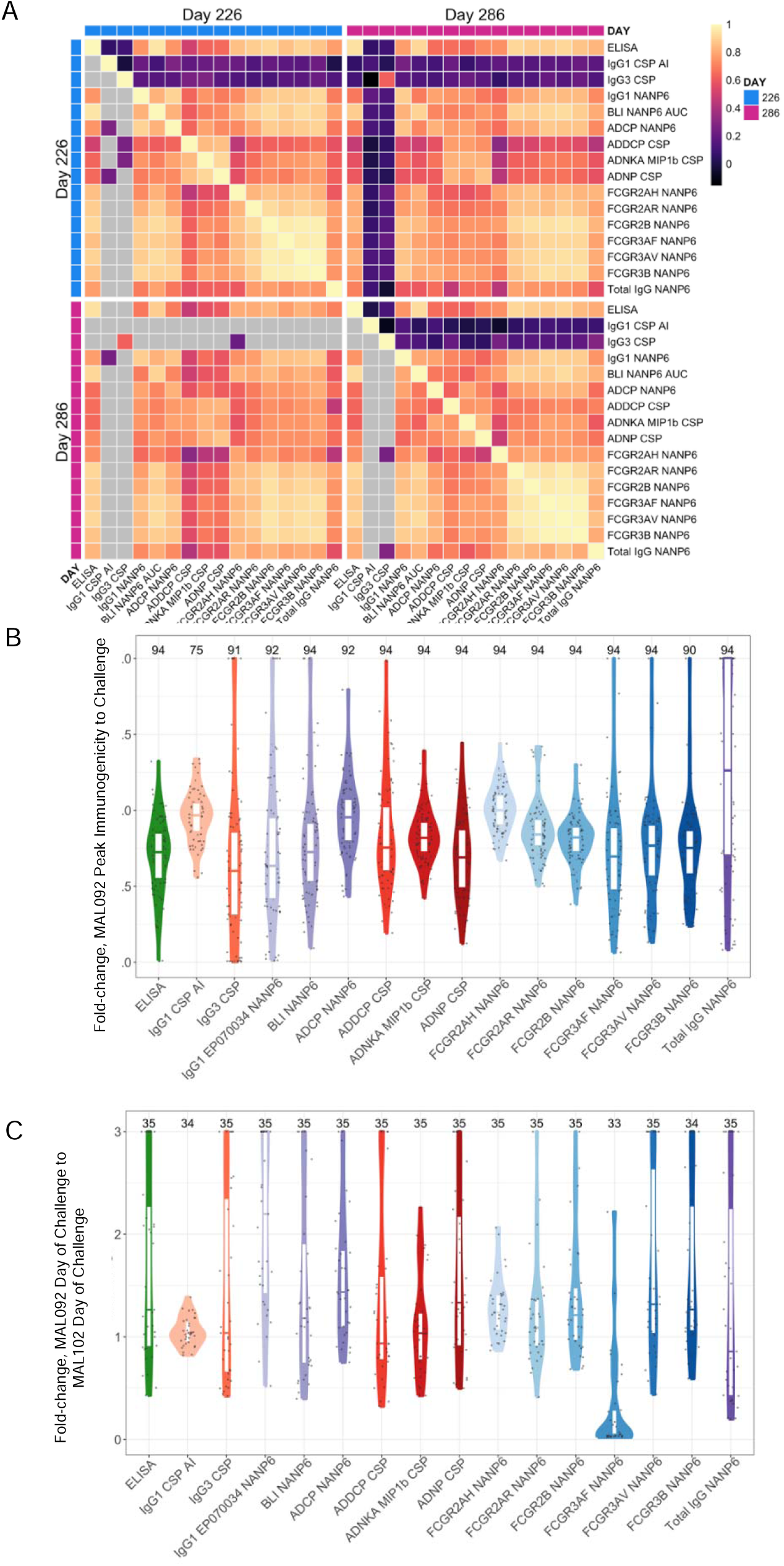
Biomarker analysis at peak immunogenicity and day of challenge in MAL092. **(A) Spearman’s rank correlation coefficients for biomarker levels at Days 226 and 286 for MAL092.** Grey cells in the lower triangle of each heatmap indicate non-significant correlation coefficients (adjusted p > 0.05). The colors in the upper triangle represent the correlation coefficient values. For significant correlations, the lower triangle uses the same color as the corresponding upper triangle. If not significant, it remains grey in the lower triangle. Day 226 = peak immunogenicity; Day 286 = day of challenge. (B) Fold-change from Day 226 (peak immunogenicity) to Day 286 (day of challenge) for each immune readout measured in the MAL092 study. Fold-change was calculated as the titer at Day 286 divided by the titer at Day 226 in MAL092 study (MAL092:Day 286/MAL092:Day 226). Each dot represents the fold-change for one study participant and a given readout. The numbers at the top of the plot correspond to the numbers of participants for whom data is plotted for each biomarker. The distribution of fold-change of each readout is described by a violin plot. Fold-changes greater than 2 were set to 2 to improve visualization. A fold change of 1 indicates no difference in antibody levels between Day 226 and Day 286. A value less than 1 indicates that antibody levels decreased over this period, while a value greater than 1 indicates that they increased over this period. (C) Fold-change from Day 286 (day of challenge in MAL092 study) to Day 22 (day of challenge in MAL102 study) for each immune readout. Fold-change was calculated as the titer at Day 22 in MAL102 divided by the titer at Day 286 in MAL092 (MAL102:Day 22/MAL092:Day 286). Fold-changes greater than 3 were set to 3 to improve data visualization. In (B) and (C), the horizontal line on each plot designates the median response. ADCP, antibody-dependent cellular phagocytosis; ADDCP, antibody dependent dendritic cell phagocytosis; ADNKA, antibody-dependent natural killer cell activation; ADNP, antibody-dependent neutrophil phagocytosis; AI, avidity index; BLI, bio-layer interferometry; CSP, circumsporozoite protein; ELISA, enzyme-linked immunosorbent assay; IgG1, immunoglobulin G1; FCGR, Fc gamma receptor; IgG3, immunoglobulin G3. Biomarker nomenclature: BLI NANP6: BLI NANP6 Dissociation AUC. ELISA: CSP IgG ELISA.

### Assessment of immune response durability identifies multiple biomarkers with slower decay rates in protected versus non-protected MAL092 participants

Since the rate at which vaccine-induced antibodies decline may influence RTS,S-mediated protection, we evaluated antibody response durability in MAL092 using the fold-change from peak immunogenicity to the day of challenge for each biomarker (Figure 4B). Median fold-change varied over biomarkers from 0.60 for IgG3 CSP to 1.26 for Total IgG NANP6. CSP IgG ELISA and BLI NANP6 Dissociation AUC had similar median fold-changes of approximately 0.75, implying a median decrease of approximately 12.5% per month from peak (assuming a constant decay rate). The most durable responses, including IgG1 CSP avidity index, ADCP NANP6, and FCGR2AH NANP6, had median fold-changes close to 1.0. Figure 4C shows an analogous plot for fold-change from Day 286 (MAL092 day of challenge) to Day 22 (MAL102 day of challenge) for each immune readout, where higher antibody responses tended to be on the day of challenge in MAL102 vs. on the day of challenge in MAL092 (i.e., most fold-changes >1), with the exception of FCGR3AF NANP6. Supplementary Tables 5 and 6 list counts and median fold-changes by protection status for the data shown in Figure 4B and 4C.

We examined whether antibody decay rates differed by protection status using linear regression models of fold-change as a function of protection status for each readout (Supplementary Table 7). Several biomarkers were associated with slower decay in protected versus non-protected participants, including CSP IgG ELISA, ADDCP CSP, FCGR2B NANP6, and FCGR3AV NANP6 (unadjusted p < 0.05); FCGR2AR NANP6 showed a borderline association (p = 0.052). These findings support evaluation of these biomarkers as candidate CoPs in field studies where transmission intensity may vary over time.

In contrast, antibody decay did not differ by protection status for the three most durable antibody responses — IgG1 CSP avidity index, ADCP NANP6, and binding of NANP6-specific antibodies to the FCGR2A H131 variant (FCGR2AH NANP6) (p > 0.32). These are functional readouts, such that antibody function may be preserved independently of titer (for avidity index at least). For these biomarkers, durability and these biomarkers themselves are unlikely to explain protection differences in MAL092. Because these readouts are strongly correlated with other biomarkers, they may still act as statistical (non-mechanistic) correlates of protection in CHMI settings.

### Correlates analysis

#### Day-of-challenge NANP6 FcγRs binding responses associate with protection

Distributions of day-of-challenge levels of the 16 biomarkers are shown by study, stratified by protection status in participants who contributed immunological data to the analyses performed here, in Figure 5 and Supplementary Figures 8 and 9 (accompanying summary statistics are provided in Supplementary Tables 8-11). Overall, biomarker levels tended to be higher in protected than non-protected participants. For some biomarkers, this was true for MAL092 only, (e.g., CSP IgG ELISA); for others, MAL102 only (e.g. ADCP NANP6); and for others, this was true in both trials (BLI NANP6, FCGR3B NANP6) (Figure 5).

**Figure 5.**
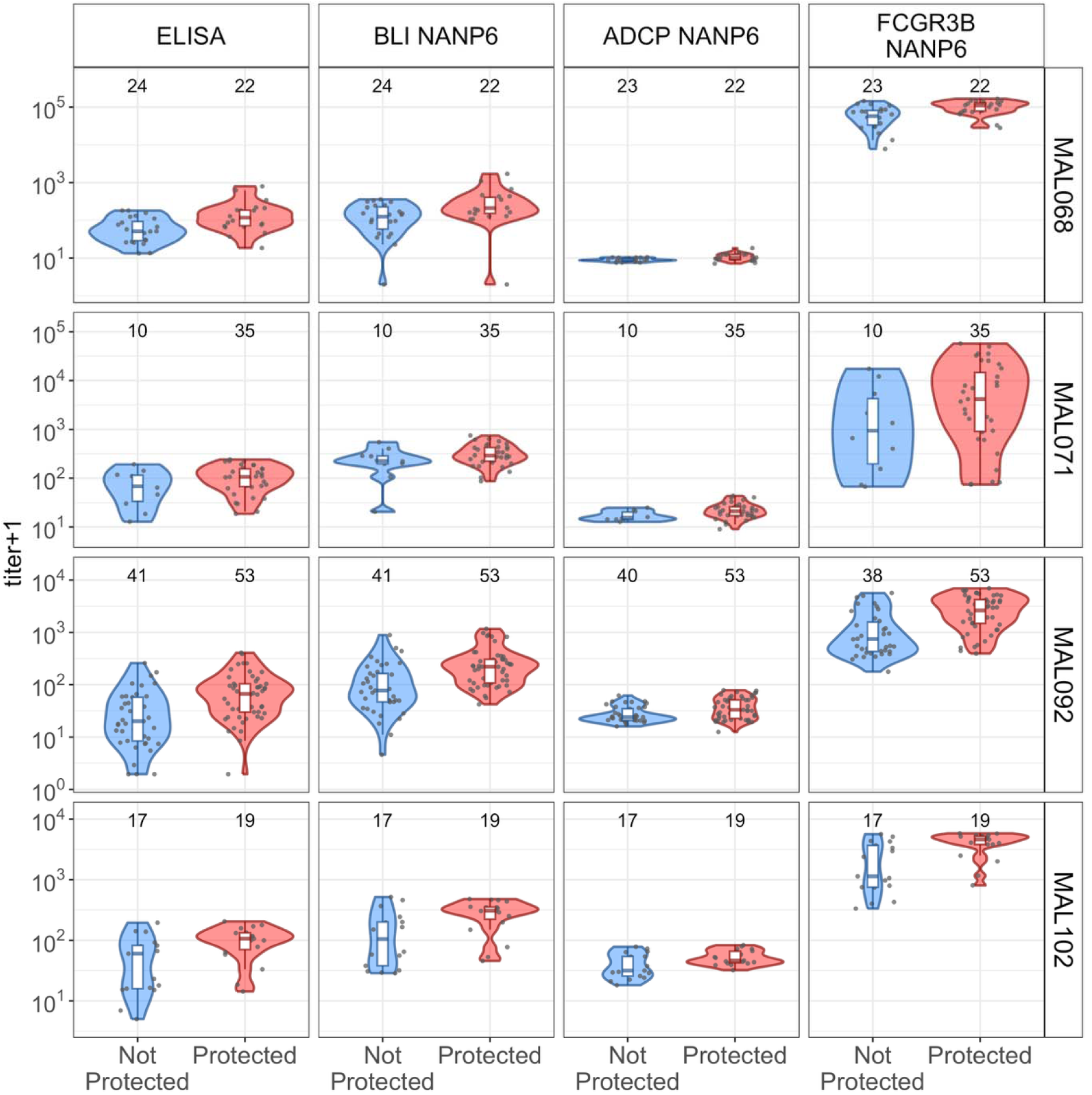
Distributions of day-of-challenge levels of selected biomarkers (ELISA, BLI NANP6, FCGR3B NANP6, and ADCP NANP6) in each of the four trials (MAL068, MAL071, MAL092, and MAL102), shown separately by protection status. Blue = not protected; red = protected. Each dot indicates one study participant. The y-axis of each plot represents the value of the measured titer. ADCP, antibody-dependent cellular phagocytosis; BLI, bio-layer interferometry; CSP, circumsporozoite protein; ELISA, enzyme-linked immunosorbent assay; FCGR, Fc gamma receptor. Biomarker nomenclature: BLI NANP6: BLI NANP6 Dissociation AUC. ELISA: CSP IgG ELISA.

Next, we fit univariate logistic regression models in MAL092 and MAL102 to evaluate associations between day-of-challenge biomarker levels and protection after CHMI. As shown in Table 1, biomarker level was significantly associated with protection status for 14 of the 16 biomarkers in MAL092 and 12 of the 16 biomarkers in MAL102 (adjusted p < 0.10). In MAL092, the largest three significant associations were observed for FCGR2AH NANP6 (coefficient: 9.73, adjusted p-value: <0.001), FCGR2AR NANP6 (6.11, <0.001), and FCGR2B NANP6 (4.91, <0.001). The same three biomarkers ranked highest in MAL102 as follows: FCGR2AH NANP6 (21.88, 0.029), FCGR2AR NANP6 (6.84, 0.029), and FCGR2B NANP6 (5.51, 0.029). For all biomarkers with significant associations, higher day-of-challenge levels were associated with a higher probability of protection.

**Table 1.** Results from univariate binary logistic regression modeling the probability of protection post-malaria challenge based on the day-of-challenge level of each biomarker. A separate logistic regression analysis was performed for each biomarker in both MAL092 (left) and MAL102 (right). For each study and biomarker, the table lists the regression coefficient measuring the association with protection status (Coefficient), the p-value of the corresponding coefficient, the adjusted p-value, and the Bayes Information Criterion (BIC). Smaller BIC values suggest better statistical models for predicting protection status. A bold adjusted p-value means that the association was significant (defined as adjusted p-value < 0.10).

|  | MAL092 |  |  |  |  | MAL102 |  |  |  |  |
| --- | --- | --- | --- | --- | --- | --- | --- | --- | --- | --- |
| Biomarker | Coefficient | Odds ratio | P-value | Adj. p-value | BIC | Coefficient | Odds ratio | P-value | Adj. p-value | BIC |
| CSP IgG ELISA | 1.85 | 6.3 | <0.001 | **<0.001** | 120.8 | 2.28 | 9.7 | 0.019 | **0.034** | 50.0 |
| IgG1 CSP AI | 1.37 | 3.9 | 0.66 | 0.66 | 137.7 | 10.58 | 39153.7 | 0.23 | 0.259 | 55.3 |
| IgG3 CSP | 4.36 | 77.9 | 0.22 | 0.23 | 136.2 | 4.68 | 107.2 | 0.34 | 0.36 | 55.9 |
| IgG1 EP070034 NANP6 | 1.84 | 6.3 | <0.001 | **<0.001** | 120.7 | 3.46 | 31.7 | 0.006 | **0.029** | 44.7 |
| BLI NANP6 Dissociation AUC | 2.61 | 13.5 | <0.001 | **<0.001** | 116.7 | 2.55 | 12.9 | 0.011 | **0.029** | 49.0 |
| ADCP NANP6 | 3.81 | 45.1 | 0.003 | **0.004** | 127.7 | 5.21 | 182.9 | 0.024 | **0.038** | 50.8 |
| ADDCP CSP | 2.15 | 8.6 | 0.017 | **0.021** | 131.9 | 5.17 | 176.4 | 0.052 | **0.069** | 52.5 |
| ADNKA MIP1b CSP | 3.69 | 40.0 | 0.013 | **0.017** | 131.2 | 3.7 | 40.3 | 0.195 | 0.24 | 55.2 |
| ADNP CSP | 1.29 | 3.6 | 0.036 | **0.041** | 133.1 | 2.48 | 12.0 | 0.029 | **0.042** | 51.4 |
| FCGR2AH NANP6 | 9.73 | 16835.4 | <0.001 | **<0.001** | 120.9 | 21.88 | 3184386000 | 0.008 | **0.029** | 46.3 |
| FCGR2AR NANP6 | 6.11 | 452.2 | <0.001 | **<0.001** | 115.5 | 6.84 | 933.7 | 0.009 | **0.029** | 46.4 |
| FCGR2B NANP6 | 4.91 | 135.3 | <0.001 | **<0.001** | 114.2 | 5.51 | 246 | 0.009 | **0.029** | 47.1 |
| FCGR3AF NANP6 | 1.68 | 5.4 | <0.001 | **<0.001** | 117.7 | -0.1 | 0.9 | 0.94 | 0.94 | 57.0 |
| FCGR3AV NANP6 | 1.99 | 7.3 | <0.001 | **<0.001** | 116.6 | 3.12 | 22.7 | 0.015 | **0.03** | 46.8 |
| FCGR3B NANP6 | 2.7 | 14.8 | <0.001 | **<0.001** | 114.7 | 3.34 | 28.3 | 0.007 | 0.029 | 46.7 |
| Total IgG NANP6 | 1.66 | 5.3 | 0.002 | **0.0032** | 126.1 | 3.11 | 22.4 | 0.013 | **0.03** | 46.6 |

ADCP, antibody-dependent cellular phagocytosis; ADDCP, antibody dependent dendritic cell phagocytosis; ADNKA, antibody-dependent natural killer cell activation; ADNP, antibody-dependent neutrophil phagocytosis; AI, avidity index; BIC, Bayes Information Criterion; BLI, bio-layer interferometry; CSP, circumsporozoite protein; ELISA, enzyme-linked immunosorbent assay; IgG1, immunoglobulin G1; FCGR, Fc gamma receptor; IgG3, immunoglobulin G3.

Figure 6 shows the predicted probabilities of protection versus day-of-challenge biomarker titer in MAL092 and in MAL102. In general, biomarker curves appeared similar between MAL092 and MAL102, with the probability of protection increasing sharply with titer for all 16 biomarkers in MAL092 and 15 of 16 biomarkers in MAL102 (FCGR3AF NANP6 being the exception). The models also estimate titer thresholds associated with high protection probabilities. For CSP IgG ELISA, which is considered as the current benchmark, the predicted probability of protection exceeded 0.80 at titers > 147 ELISA units/ml in MAL092 and > 217 ELISA units/ml in MAL102; moreover, the ADNP CSP curve appeared very similar to the CSP IgG ELISA curve in both studies. The interpretation of these curves to understand protection induced by a given vaccine regimen can be done using the distribution of antibody titers at the time of exposure. Supplementary Figure 10 shows similar patterns in MAL068 and MAL071, with 0.80-probability CSP IgG ELISA thresholds of 245 and 88 ELISA units/ml, respectively.

**Figure 6.**
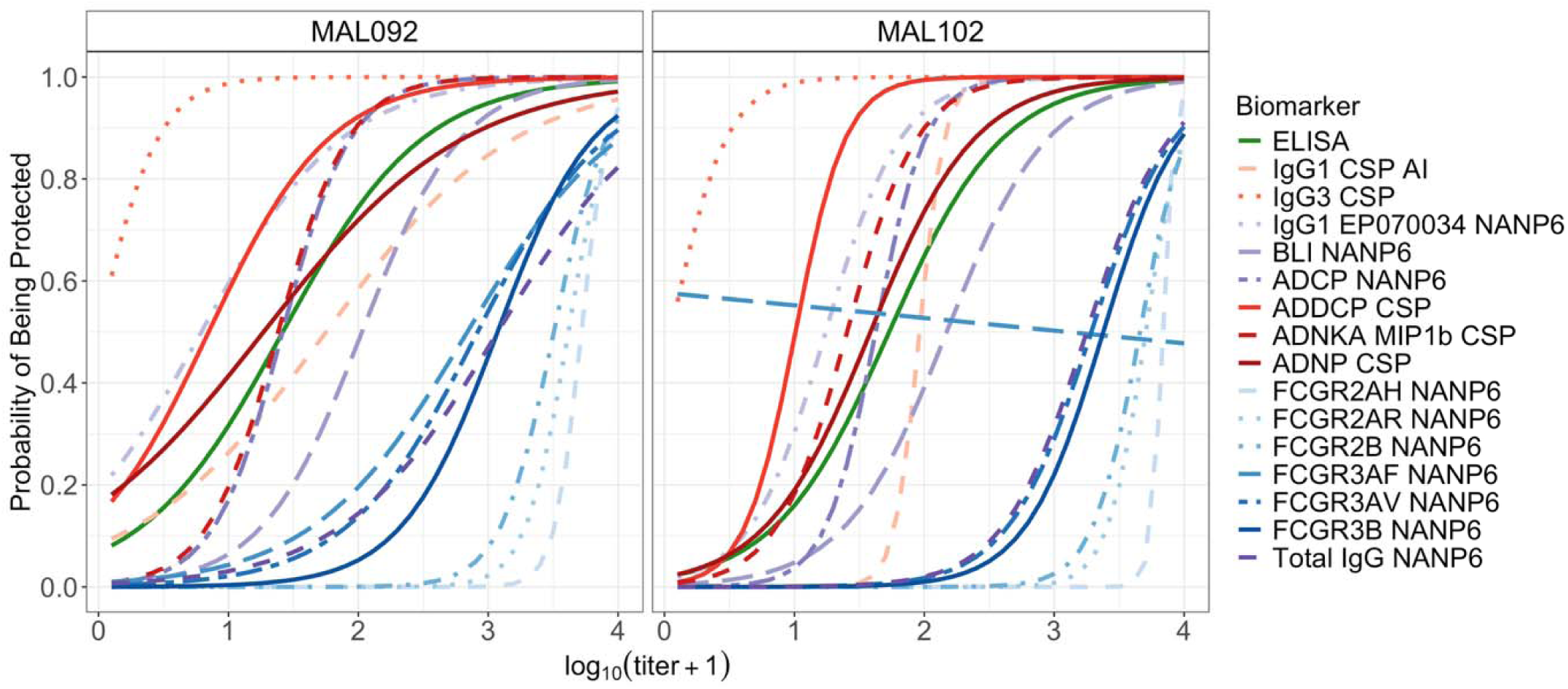
Probability of being protected as a function of the day-of-challenge level of each biomarker (one curve per biomarker) in MAL092 (left) and MAL102 (right), as predicted by the univariate logistic regression model summarized in Table 1. ADCP, antibody-dependent cellular phagocytosis; ADDCP, antibody dependent dendritic cell phagocytosis; ADNKA, antibody-dependent natural killer cell activation; ADNP, antibody-dependent neutrophil phagocytosis; AI, avidity index; BLI, bio-layer interferometry; CSP, circumsporozoite protein; ELISA, enzyme-linked immunosorbent assay; IgG1, immunoglobulin G1; FCGR, Fc gamma receptor; IgG3, immunoglobulin G3. Biomarker nomenclature: BLI NANP6: BLI NANP6 Dissociation AUC. ELISA: CSP IgG ELISA.

Altogether, these results support the majority of the biomarkers selected for the present analysis as candidate CoPs in both MAL092 and MAL102.

#### Multivariable analyses provide mixed evidence that NANP6-specific antibody levels add information beyond CSP IgG ELISA in MAL092

We fit multivariable logistic regression models (eleven prespecified models) to assess whether certain biomarkers or combinations thereof improved prediction of protection status beyond CSP IgG ELISA (i.e., significantly associated with protection status when CSP IgG ELISA was accounted for), and to identify which combinations of biomarkers were most strongly associated with protection. These analyses modeled the probability of protection after challenge in MAL092 (Supplementary Table 12) and MAL102 (Supplementary Table 13) as a function of the day-of-challenge levels of two and of three preselected biomarkers, with CSP IgG ELISA always included in all specifications. In MAL092, bivariate models suggested that non-CSP IgG ELISA biomarkers retained borderline or nominal significance after adjustment for CSP IgG ELISA, including BLI NANP6 Dissociation AUC (p = 0.051), FCGR3AF NANP6 (p = 0.054), and FCGR3AV NANP6 (p = 0.04) (Supplementary Table 12). These results suggest that certain NANP6-specific biomarkers may augment the predictive value of CSP IgG ELISA. However, in MAL102, where participants were re-challenged following a booster dose, none of the non-CSP IgG ELISA biomarkers included in a bivariate or trivariate model had a p-value < 0.05 (“Biomarker 2” column, Supplementary Table 13), suggesting that CSP IgG ELISA was the best candidate CoP in this study.

We next evaluated the predictive performance of each of the 16 biomarkers independently in each CHMI study. We used each biomarker as a predictive score, and classified participants as protected if the biomarker level exceeded a chosen threshold and as non-protected otherwise. Results are shown in Figure 7 and Supplementary Figures 11-13.

**Figure 7.**
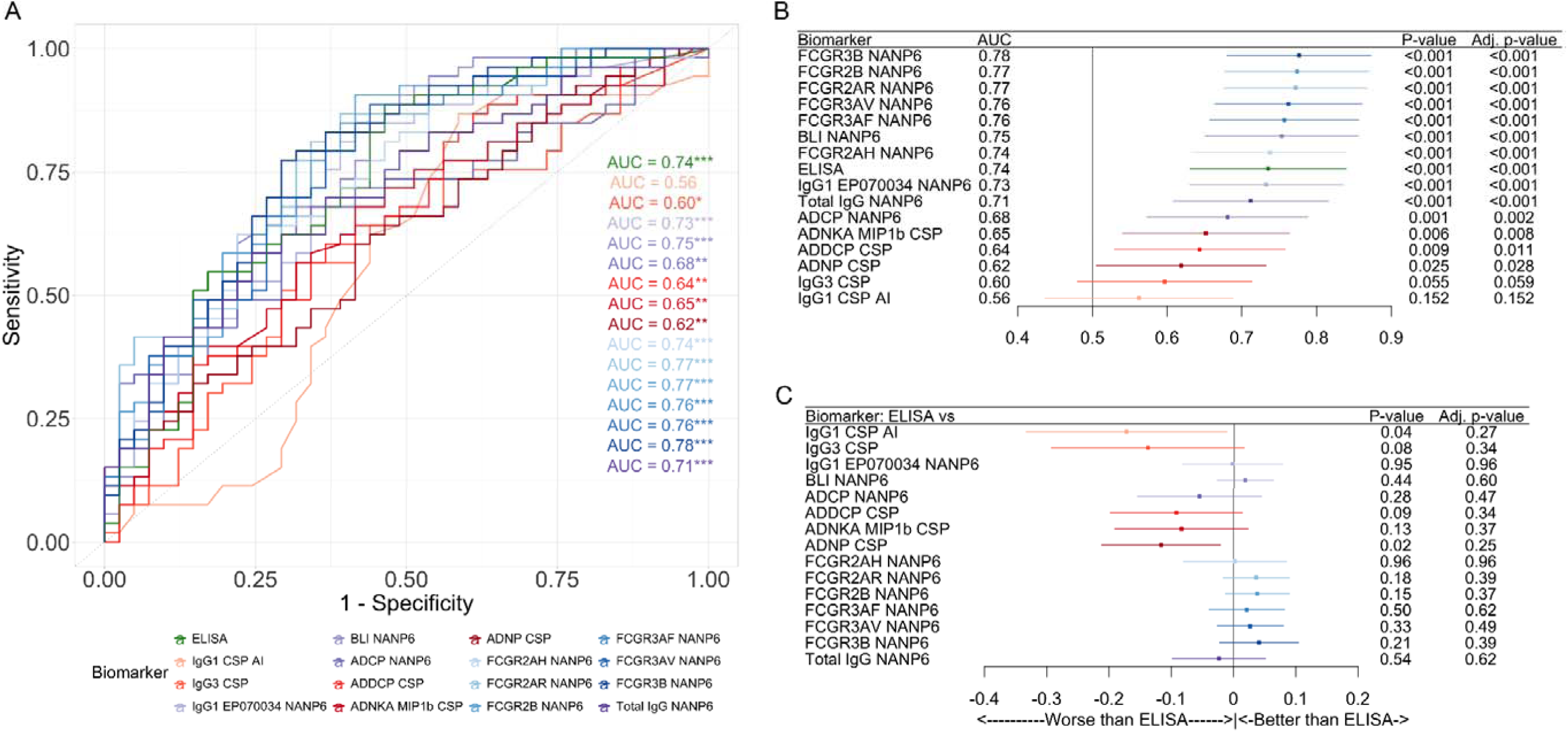
Predictive performance of each biomarker in MAL092. (A) Receiver operating characteristic (ROC) curve of each individual biomarker, and their area under the curve (AUC). (B) AUCs and 95% confidence intervals of each biomarker. A value of 0.5 for the AUC indicates no predictive power, whereas a value in (0.5; 1.0] indicates predictive performances, and perfect predictions with a value of 1.0. The horizontal line extending on either side of each dot represents the 95% confidence interval of the AUC. The p-value assesses the significance of AUC (i.e., AUC = 0.5 or AUC > 0.5). Adjusted p-values that control the false discovery rate for multiple comparisons are provided in the right column. (C) Differences between the AUC from each biomarker and the AUC from CSP IgG ELISA. A value of 0 indicates no difference in AUC, whereas a positive or negative value indicates better or worse performances for the corresponding biomarker relative to CSP IgG ELISA. The horizontal line extending on either side of each dot represents the 95% confidence interval of the difference between the AUC. The p-value assesses the significance of the difference between the AUC. Adjusted p-values that control the false discovery rate for multiple comparisons are provided in the right column of the figure. In (A-C), the color scheme distinguishes the class of antibodies targeted by the readout, with orange representing CSP, blue representing NANP6, and green indicating the CSP IgG ELISA readout. ADCP, antibody-dependent cellular phagocytosis; ADDCP, antibody dependent dendritic cell phagocytosis; ADNKA, antibody-dependent natural killer cell activation; ADNP, antibody-dependent neutrophil phagocytosis; AI, avidity index; BLI, bio-layer interferometry; CSP, circumsporozoite protein; ELISA, enzyme-linked immunosorbent assay; IgG1, immunoglobulin G1; FCGR, Fc gamma receptor; IgG3, immunoglobulin G3. Biomarker nomenclature: BLI NANP6: BLI NANP6 Dissociation AUC. ELISA: CSP IgG ELISA.

Figure 7 summarizes predictive performance in MAL092. Fourteen of the 16 biomarkers showed significantly predictive ability (adjusted p < 0.05), with AUCs ranging from 0.62 (95% CI: 0.51 – 0.73; ADNP CSP) to 0.78 (95% CI: 0.68 – 0.87; FCGR3B NANP6). The CSP IgG ELISA biomarker achieved an AUC of 0.74 (95% CI: 0.63 – 0.84). Notably, the highest AUCs were observed for FCGR NANP6 biomarkers (FCGR3B NANP6, FCGR2AH NANP6, FCGR2AR NANP6, FCGR3AV NANP6, and FCGR3AF NANP6), with AUCs ranging from 0.76 to 0.78. While these AUCs were numerically higher than that of CSP IgG ELISA, none represented a significant improvement over CSP IgG ELISA (all p values > 0.15; all adjusted p-values > 0.37). NANP6-specific biomarkers consistently outranked CSP-specific biomarkers, aligning with earlier findings from the MAL068 and MAL071 studies (51). These CSP-specific biomarkers also did not outperform the predictive ability of CSP IgG ELISA; thus, this analysis of MAL092 did not identify any single biomarker with significantly better predictive performance than CSP IgG ELISA.

Supplementary Figure 11 shows the MAL102 results, which were similar to those in MAL092, with the exception that FCGR3AF NANP6 departed from the other FCGR biomarkers in having less-predictive capacity compared to CSP IgG ELISA (albeit, again, with this difference not statistically significant). As in MAL092, no single biomarker, whether CSP- or NANP6-specific, exhibited significantly better predictive performance than CSP IgG ELISA in MAL102.

Supplementary Figures 12 and 13 present analogous results for MAL068 and MAL071. Overall trends in predictive performances were consistent across studies. Supplementary Figure 14 corroborates this by comparing the predictive performances of each biomarker in MAL092 with that in MAL102, MAL068, and MAL071.

Altogether, these multivariable analyses provide some evidence that incorporation of NANP6-specific biomarkers contributes additional predictive value beyond CSP IgG ELISA in MAL092. However, in MAL102, they showed that none of the multivariate models provided improved explanatory value compared to the model with CSP IgG ELISA alone, which was deemed the best candidate CoP. These conclusions are restricted to the eleven pre-specified models considered in the multivariable analysis.

#### Multivariate signatures did not significantly improve predictive performance over the benchmark NANP-based CSP IgG ELISA biomarker in MAL092 or MAL102

An informative biomarker combination would ideally include a small set of biomarkers that each contributes independent (non-redundant) information about protection. We therefore tested whether improved predictive performance compared to that of the CSP IgG ELISA biomarker could be achieved by incorporating additional biomarkers into the model. We trained prespecified uni-, bi- and trivariate models, as well as a full model including all 16 biomarkers, on data from MAL068 and 071 (CV-AUC of the ROC curve is reported as metric of predictive performances). Next, we evaluated these models by predicting protection in MAL092 or MAL102 (validation ROC AUC). All models included CSP IgG ELISA as a benchmark for comparison. Figure 8 shows results for MAL092. CSP IgG ELISA alone achieved an AUC of 0.74 (95% CI: 0.63 – 0.84), whereas the primary multivariate panel of 11 bi- or tri-variate models yielded AUCs ranging from 0.73 to 0.76. The substantial overlap in 95% CIs indicated that none of the 11 multivariate models provided significant improvement in predictive power compared to CSP IgG ELISA alone.

**Figure 8.**
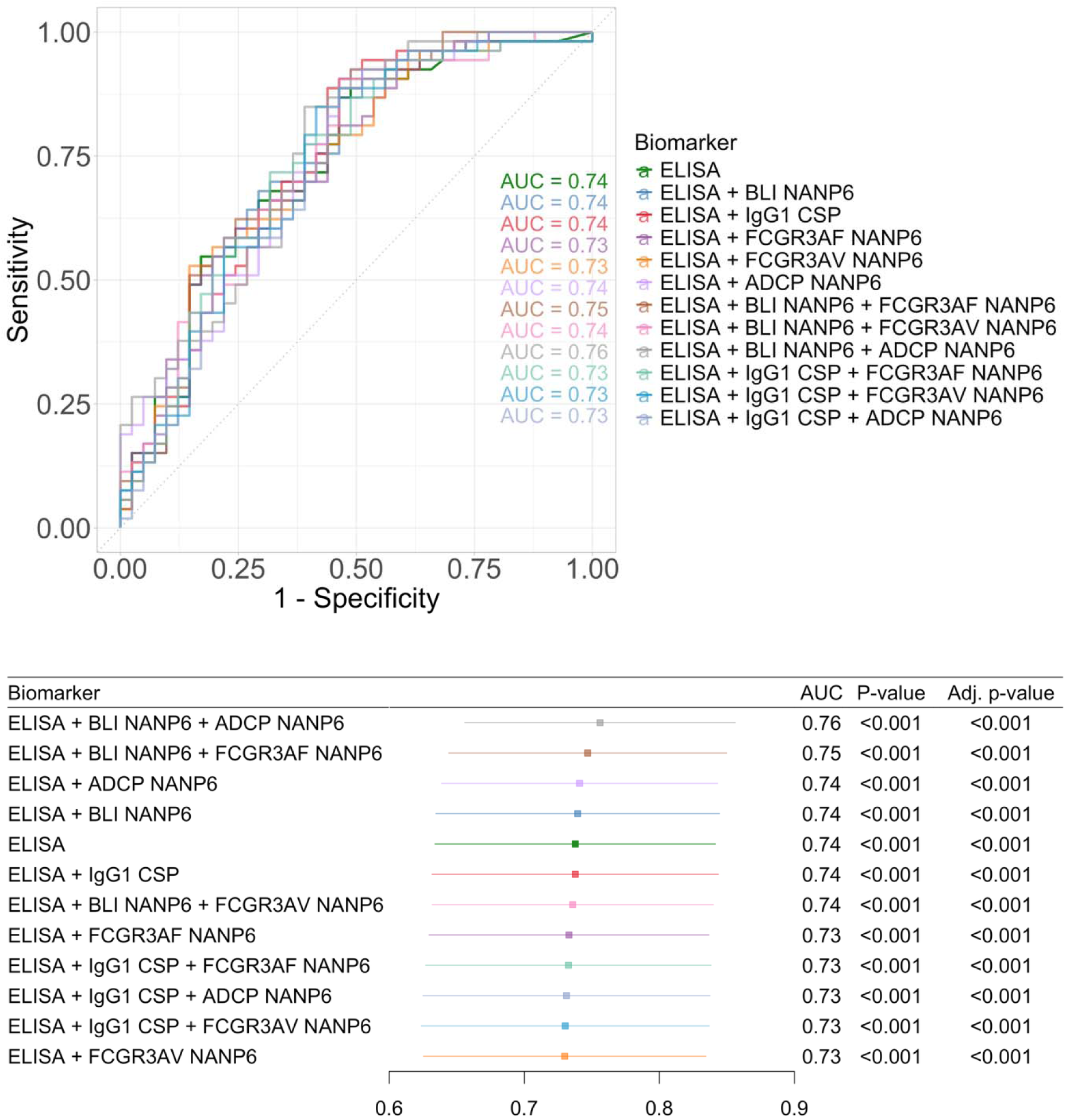
Predictive performances of the 11 prespecified multivariate signatures in MAL092. ADCP, antibody-dependent cellular phagocytosis; AUC, area under the ROC curve; BLI, bio-layer interferometry; CSP, circumsporozoite protein; ELISA, enzyme-linked immunosorbent assay; IgG1, immunoglobulin G1; FCGR, Fc gamma receptor; IgG3, immunoglobulin G3. Biomarker nomenclature: BLI NANP6: BLI NANP6 Dissociation AUC. ELISA: CSP IgG ELISA.

In MAL102, CSP IgG ELISA alone achieved an AUC of 0.71 (95% CI: 0.53 – 0.89) (Figure 9). The primary multivariate models yielded AUCs ranging from 0.71 to 0.82. Unlike in MAL092, two models stood out with higher point-estimate performance (AUC ≥ 0.80), although 95% CIs still substantially overlapped across models. These two models were CSP IgG ELISA + BLI NANP6 Dissociation AUC + ADCP NANP6 (AUC = 0.82) and CSP IgG ELISA + IgG1 CSP (AUC = 0.80).

**Figure 9.**
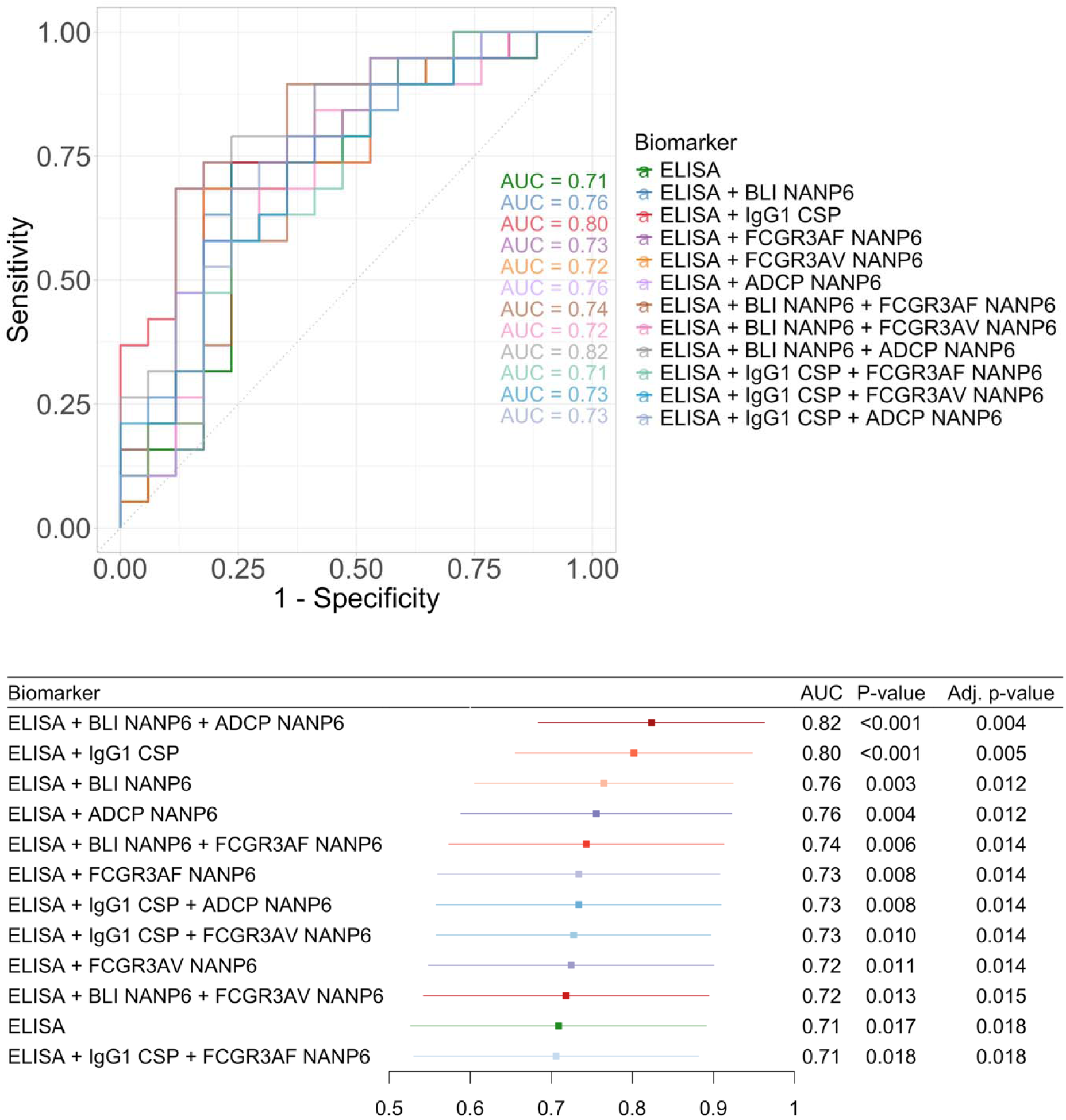
Predictive performances of the 11 prespecified multivariate signatures in MAL102. AUC = area under the ROC curve. ADCP, antibody-dependent cellular phagocytosis; AUC, area under the ROC curve; BLI, bio-layer interferometry; CSP, circumsporozoite protein; ELISA, enzyme-linked immunosorbent assay; IgG1, immunoglobulin G1; FCGR, Fc gamma receptor; IgG3, immunoglobulin G3. Biomarker nomenclature: BLI NANP6: BLI NANP6 Dissociation AUC. ELISA: CSP IgG ELISA.

Results for the secondary set of models are shown in Supplementary Figures 15 (MAL092) and 16 (MAL102). The multivariate model with the best predictive performances within the MAL068 and 071 trials included BLI NANP6 Dissociation AUC, ADCP NANP6, and FCGR3AV (CV-AUC = 0.80), while the second-best model included CSP IgG ELISA, IgG1 CSP WRAIR avidity index, and ADCP NANP6 (CV-AUC = 0.79). When the MAL068 and MAL071-trained models were applied to predict outcomes in MAL092 and MAL102, the model with the best (external) validation AUC in MAL092 was a univariate model with FCGR2B NANP6 (validation AUC = 0.77). In contrast, the model with the best validation AUC in MAL102 was a multivariate model with CSP IgG ELISA, BLI NANP6 Dissociation AUC, and ADCP NANP6 (validation AUC = 0.82). The best-performing univariate model applied to MAL102, which was also a model based on FCGR2B NANP6, yielded similar results (validation AUC = 0.80). Likewise, the best-performing multivariate model applied to MAL092 (based on BLI NANP6 Dissociation AUC and ADCP NANP6) yielded similar results (validation AUC = 0.76). In both MAL092 and MAL102, the 95% confidence intervals for the highest validation AUCs overlapped substantially, indicating no clear separation among the top-performing models and no evidence that any model outperformed CSP IgG ELISA alone.

Altogether, the multivariate models did not show significantly improved predictive performance over the benchmark NANP-based CSP IgG ELISA biomarker in MAL092 or MAL102.

## Discussion

Multiple studies highlight the critical role of CoPs across all stages of vaccine development and deployment for infectious diseases. Neutralizing antibodies against SARS-CoV-2 (62) and hemagglutination inhibition titers for influenza (63) are established CoPs for their respective vaccines, and defined antibody thresholds are widely accepted as a CoP for vaccines against measles, human papillomavirus, pneumococcal disease, meningococcal serogroups, and hepatitis B, serving as measurable benchmarks of protective immunity (32–34). During development, CoPs serve as reference points for comparing new vaccine candidates with existing ones, particularly regarding the strength and persistence of immune responses, thereby informing design and optimization strategies. CoPs support regulatory decisions by providing early indications of a vaccine’s likely effectiveness in broader populations, helping to streamline approval and strengthen confidence in its clinical value. After licensure, CoPs remain essential for evaluating real-world vaccine performance, establishing immunological targets, guiding dosing regimens, and determining the need for booster doses based on the durability of protection. In this way, well-defined CoPs act as early markers of efficacy, facilitating the faster development and approval of more effective vaccines. However, identifying CoPs for malaria vaccines is challenging due in part to heterogeneity across studies, including differences in populations, trial designs, endpoints, and a lack of standardized immunological assays (64). The biological complexity of the parasite (65, 66) compounds this complexity, as does the limited understanding of protective mechanisms, underexplored functional assays, and uncertainty in translating findings from CHMI studies in adults to endemic populations, particularly children (67). CoP discovery could advance the development, implementation, and monitoring of malaria vaccines.

Extensive research has leveraged the rich immunological data from the MAL068 and MAL071 trials to identify CoPs for the RTS,S vaccine in CHMI studies (56). While recent analyses have begun to demonstrate the translatability of these candidate CoPs to independent CHMI studies, including MAL092 and its follow-up study MAL102 (56), our work extends these findings by evaluating a broader set of 16 biomarkers. Additionally, we present longitudinal immune response data from MAL092 and the MAL102 follow-up study for these Fc effector and FcγR-binding biomarkers for the first time. Our three main findings are summarized as follows.

First, of the 16 day-of-challenge biomarkers previously identified as candidate CoPs in MAL068 and MAL071 and available for analysis in MAL092 and MAL102, 14 remained significantly predictive of protection in MAL092, and 12 remained significantly predictive of protection in MAL102. Across these sets of 14 and 12, 10 biomarkers were significantly predictive of protection in both studies (CSP IgG ELISA, IgG1 NANP6, BLI NANP6 Dissociation AUC, ADCP NANP6, ADDCP CSP, ADNP CSP, FCGR2AH NANP6, FCGR2AR NANP6, FCGR2B NANP6, FCGR3AV NANP6, FCGR3B NANP6, and Total IgG NANP6). Given that biomarkers were included in the present analysis based largely on performing well in cross-study analysis in MAL068 and MAL071, our work demonstrates high consistency in performance for these 10 biomarkers across all four studies. This result is particularly notable in MAL092 where CHMI occurred three months after the last vaccination rather than the usual three to four weeks, extending evidence for these candidate CoPs to a later post-vaccination timepoint. The biomarkers showing the strongest predictive performance were predominantly Fc effector-related or involved FcγR specificity of NANP6-targeting antibodies. In both MAL092 and MAL102, the top-performing biomarkers (i.e., largest CV-AUC) pertained to FcγR binding, especially those reflecting binding of NANP6-targeting antibodies to the FcγRIII receptor [CD16, which preferentially engage IgG1 and IgG3 over IgG2 and IgG4 (68)] and to the FcγRII receptor [with highest specificity for IgG3 (69)]. The latter biomarkers included FcγRIIa H131 [allelic variant with higher affinity for IgG1 and IgG2 than the R131 variant (68, 69)], FcγRIIa R131, and FcγRIIb. The reproducibility of these candidate CoPs across multiple CHMI studies increases confidence in their potential and supports their prioritization in field studies. These findings confirm and extend those reported by Spreng et al. in MAL092, who identified five day-of-challenge biomarkers predictive of protection (serum Ig CSP, NANP6, N Interface AUC_diss_, and IgG1 CSP and NANP6 334-hIgG1 equivalent concentration) (56). Consistent with Spreng et al, we also found no evidence that IgG1 CSP avidity index or IgG3 CSP was associated with protection in MAL092.

Second, no single candidate CoP significantly outperformed the standard CSP IgG ELISA biomarker at predicting infection post CHMI in MAL092 and MAL102, and adding biomarkers did not significantly improve prediction beyond CSP IgG ELISA alone. MAL092 multivariable models suggested independent associations for FCGR3AV NANP6, and possibly for BLI NANP6 Dissociation AUC and FCGR3AF NANP6. However, after adjusting for CSP IgG ELISA, this finding was not replicated in MAL102 (in which a subset of MAL092 participants were re-challenged after a booster dose). Predictive-performance patterns were consistent across all four CHMI studies: NANP6-specific biomarkers generally outperformed CSP-specific biomarkers, yet no individual biomarker significantly outperformed CSP IgG ELISA (e.g., in MAL092, all p > 0.15). While Young et al. did not perform direct comparisons between other top-performing biomarkers and CSP IgG ELISA, CSP IgG ELISA was identified as a top-performing biomarker in multiple analyses in Young et al. (summarized in Supplementary Table 3 in the present work); therefore, our results are largely consistent with the MAL068 and MAL071 CHMI prediction results. The anti-CSP ELISA assay using the R32LR coating antigen offers significant practical advantages over more resource-intensive antibody effector function assays. This assay has been used in validation studies submitted to the European Medicines Agency to evaluate lot-to-lot consistency and co-administration with other childhood vaccines for RTS,S (64).

Third, four biomarkers exhibited slower decay in protected versus non-protected MAL092 participants: ELISA-measured IgG antibodies directed against the CSP NANP6 repeat region (CSP IgG ELISA), NANP6-specific antibody engagement of the FcγRIIb receptor, NANP6-specific antibody engagement of the FcγRIIIa F158V receptor, and CSP-specific antibody-dependent dendritic cell phagocytosis (ADDCP CSP). Since infection exposure in field settings occurs over extended and heterogeneous time windows, and transmission intensity can vary over time, biomarkers more durable in protected individuals may better translate as correlates of protection in field studies: slower waning increases the likelihood that the biomarker level at the time of exposure remains close to the level measured at the intended correlate timepoint. Conversely, biomarkers that decay rapidly can still be statistically useful for predicting participant-level outcomes within a fixed or short follow-up interval (as in CHMI), even if their predictive value attenuates when the time between measurement and exposure increases. Additional studies using blood samples from field trials would be needed to test these hypotheses.

This work has several limitations common to CHMI correlates analyses. First, findings from CHMI studies in malaria-naïve volunteers may not generalize to populations living in malaria-endemic regions where prior exposure, host immunity, and transmission intensity vary. Second, the modest sample sizes (MAL092: n = 94; MAL102: n = 36) limit power to detect moderate differences in predictive performance when comparing individual biomarkers or multivariate signatures against the benchmark CSP IgG ELISA biomarker. Third, our analyses solely focused on humoral biomarkers; however, RTS,S was also designed to elicit cellular responses (13) and some evidence suggests that cellular immunity from RTS,S contributes to protection (70). Future work integrating humoral and cellular data, through multivariate analyses and systems immunology, may better delineate their relative contributions and interplay and could potentially improve the predictive power of immune response biomarkers beyond that of the current CSP IgG ELISA benchmark.

## Methods

### Ethics Statements

#### MAL068

The MAL068 study was undertaken in accordance with the provisions of the International Conference on Harmonization and Good Clinical Practice guidelines. The MAL068 study protocol was approved by the Walter Reed Army Institute of Research (WRAIR) Institutional Review Board (IRB) and PATH-Malaria Vaccine Initiative’s Western IRB.

#### MAL071

The MAL071 study was undertaken in accordance with International Council for Harmonization of Technical Requirements for Pharmaceuticals for Human Use guidelines and good clinical practice. The MAL071 study protocol was approved by the WRAIR Institutional Review Board and the Western Institutional Review Board. After publication of the primary MAL071 paper (42), one participant withdrew consent for their data to be used in further publications, and their data are not analyzed in the present manuscript.

#### MAL092

The MAL092 study was conducted according to the protocol and in compliance with International Conference on Harmonization/Good Clinical Practice guidelines. The MAL092 study protocol was approved by the WRAIR Institutional Review Board.

#### MAL102

The MAL102 follow-up/extension study was conducted according to the protocol and in compliance with International Conference on Harmonization/Good Clinical Practice guidelines. The MAL102 study protocol was approved by the WRAIR Institutional Review Board. In each of the four studies, all participants provided written informed consent before study procedures were initiated.

### Study Samples

Out of 67 participants originally enrolled in MAL068 (41) and 63 in MAL071 (42), 46 and 45 participants, respectively, who had consented to their samples being used in subsequent investigations were included.

Out of 130 participants originally enrolled in MAL092 (43), 94 participants who had previously provided consent for future use of their samples for research were included in this analysis. For the MAL102 (44) analysis, serum samples from 36 participants from MAL092 who reenrolled in MAL102 and consented to their samples being used in subsequent investigations were included.

## Laboratory Methods

### ELISA

Binding of *P. falciparum* CSP R32LR IgG antibodies in vaccinee serum was measured by ELISA as described (71).

### BAMA

Binding measurements of *P. falciparum* CSP-specific or NANP6-specific antibodies in vaccinee serum were evaluated using BAMA as described previously (52). Briefly, antigen-specific antibody responses were measured by covalently coupling *P. falciparum* CSP antigens to fluorescent beads (Luminex Corporation) and incubating with vaccinee serum samples in BAMA assay diluent (Phosphate Buffered Saline, 5% normal goat serum (v/v), 0.05% Tween 20, 1% milk blotto (w/v)). Anti-human IgG1 (BioLegend, San Diego, CA, clone number 12G8G11) or anti-human IgG3 (BioLegend, San Diego, CA, clone number HP6047) were used to quantify *P. falciparum* specific IgG1 or IgG3 responses, as monoclonal antibody (mAb) equivalents. Subclass specific 334-hIgG1 or IgG3 (334-hIgG3) mAbs were titrated as standard curves in each assay, and equivalence units in vaccinee sera were calculated using 5-PL logistic regression. Avidity index (AI) was calculated as the percentage retained binding in sodium citrate buffer at pH 4.0 (CIT, Teknova, Hollister, CA) versus PBS buffer alone (i.e., AI = MFI [CIT]/MFI [PBS] × 100) by using a modified BAMA with the addition of a 15-minute incubation step after sample binding. This step contained either PBS or CIT followed by washing. All assays and data analyses were performed in a GCLP compliant laboratory with assay tracking via Levey-Jennings charts.

### BLI Avidity Assay

The BLI assay for measuring magnitude and avidity of NANP6-specific antibodies in vaccinee serum was performed as described previously using Fortebio Octet HTX and Octet Red384 instruments and biosensors (Fortebio-Biologics by Sartorius, San Jose, CA). Both data acquisition and analyses were performed with the United States Food and Drug Administration’s Title 21 Code of Federal Regulations Part 11 (FDA Title 21 CFR Part 11) compliant software versions (Data Acquisition 12.0 and Data Analysis HT 12.0 packages) (56, 72). Briefly, biotinlyated NANP6 and negative control peptide C1 (56, 72) were loaded onto streptavidin (SA) biosensors (threshold level set not to exceed Δλ = 1 nm). The 1:50 diluted vaccinee sera binding to NANP6 and C1 peptides were measured. Binding to C1 peptide was subtracted to obtain antigen specific binding time courses. The AUC of the dissociation curve, a composite measure of binding magnitude and avidity, was estimated for the specific binding time course data by using the trapezoidal rule in the R package ‘caTools’.

### Fc-mediated Effector Functional Assays

NANP6-specific ADCP activity, CSP-specific ADDCP activity, CSP-specific ADNKA activity, and CSP-specific ADNP activity in vaccinee serum were all measured as described (50).

### Fc-Binding Protein Array

Binding of NANP6-specific antibodies in vaccinee serum to human Fc receptors was measured as described (73, 74).

### Statistical Methods

The primary goals of the analysis were to characterize vaccine-induced immune responses and assess whether immune biomarkers previously identified as predictive of protection in the MAL068 and MAL071 trials remained predictive in the MAL092 and MAL102 trials. The analyses focused on 16 immune biomarkers, including the historical CSP IgG ELISA readout as a reference comparator.

Continuous variables (e.g., day-of-challenge biomarker levels) were compared between pairs of independent groups using Wilcoxon rank-sum tests. Paired continuous variables measured within study participants at two distinct visits were compared using Wilcoxon signed-rank tests. McNemar’s test was used to compare binary proportions between study visits estimated using paired binary data. Association between pairs of categorical variables was measured using the Cramer’s V statistic. Fold-change was defined as FC = Y(286)/Y(226); values >1 indicate increases and <1 indicate decreases in antibody levels between Days 226 and 286. We applied *t*-distributed stochastic neighbor embedding (*t*-SNE; `Rtsne` package) for dimensionality reduction to visually display patterns in immune responses captured by the 16 biomarkers across participants, study groups, and protection status (75). Associations between biomarkers were quantified using Spearman rank correlation coefficients. Univariate and multivariable logistic regression analyses were conducted to model the probability of being protected as a function of day-of-challenge biomarker levels. Odds ratios and their 95% confidence intervals were reported to quantify the association between biomarker levels and protection.

The ability of individual biomarkers to predict protection status following challenge was evaluated by building receiver operating characteristic (ROC) curves based on the biomarker levels measured on the day of challenge. Multivariate immune signatures of protection were trained using a super-learner approach (76) by stacking multiple classifiers including lasso/elastic net logistic regression, support vector machines, random forests, neural networks, and boosting. The method was implemented via the `SuperLearner` R package. Predictive performances of univariate and multivariate immune signatures were summarized using receiver operating characteristics (ROC) curves, their area under the curve (AUC), and associated p-values assessing the significance of the predictive performances of the immune signatures (i.e., testing the null hypothesis that AUC = 0.5 against the alternative hypothesis that AUC > 0.5). ROC curves were constructed using the `precrec` package. Where appropriate, associated p-values were computed using Mason and Graham’s method (77), calculated using the `verification` R package. To examine whether any of the other 15 biomarkers significantly outperformed CSP IgG ELISA at predicting protection status, their AUCs were also compared to the AUC of CSP IgG ELISA. When the model was trained and tested on data from the same studies, ROC curves were constructed via cross-validation. CSP IgG ELISA was compared with each of the remaining 15 biomarkers using the DeLong et al. method for ROC curves [(78), `pROC` package]. Pre-specified bivariate and trivariate combinations of biomarkers (each including CSP IgG ELISA titer) were evaluated for their ability to improve predictive performances over CSP IgG ELISA alone. Predictive models were trained on MAL068 and MAL071 data via 10-fold cross-validation, with data splitting stratified by protection status, and validated on MAL092 and MAL102 data.

Confidence intervals and hypothesis tests were two-sided, except for tests assessing the significance of the AUC of ROC curves. P-values less than 0.05 were treated as significant. Where appropriate, p-values were also adjusted for multiple comparisons by controlling the false discovery rate (FDR) using the Benjamini-Hochberg procedure. Adjusted p-values smaller than 0.10 were considered statistically significant.

All statistical analyses were conducted using R, version 4.4.3 (79).

#### Assay readouts

All 16 assay readouts were calculated identically to what was done for the integrative analysis of the MAL068 and MAL071 studies (51), except for three: IgG1 EP070034 NANP6 concentration, IgG3 CSP WRAIR concentration, and BLI NANP6 dissociation AUC. These three readouts were used in place of IgG1 EP070034 NANP6 magnitude, IgG3 CSP WRAIR magnitude, and BLI NANP6 score, respectively. Their calculations were modified to reduce inter-run variability and potentially improve prediction of protection status against malaria challenge.

#### Handling of missing values

Imputation of missing values was performed using the preProcess function and the bagImpute method implemented in the caret R package (80).

## Data Availability

Qualified researchers can request access to anonymized participant-level data. Further details on GSK data sharing criteria, eligibility requirements, and process for requesting access can be found at https://vivli.org/.

## Acknowledgments

We thank the participants in the MAL068, MAL071, MAL092, and MAL102 trials; Erik Jongert (GSK) and Ulrike Wille-Reece (PATH’s Center for Vaccine Innovation and Access, current affiliation: BioNTech) for contributing to study concept and design and critically reviewing the manuscript; M. Umar Qureshi for programmatic support and helpful discussions of the data generated by the Antibody Dynamics platform of the Global Health-Discovery Collaboratory (GHDC) at Duke University; and Sheetij Dutta for provision of full-length CSP protein for the BAMA and BLI assays. GSK had the opportunity to review this publication but the authors were solely responsible for the content.

## Disclaimers

The opinions and assertions contained herein are the private views of the authors and are not to be construed as official or as reflecting the views or position of the US Army, the US Department of War, the US Government, or The Henry M Jackson Foundation for the Advancement of Military Medicine, Inc.

