## Supplementary Material for "Correlates of Protection and Response Durability of the RTS,S Vaccine in Four Malaria Challenge Studies"

Supplementary Table 1. Vaccination schedules in the MAL092 and MAL102 trials.

| A. **Vaccination schedule in the MAL092 trial (1)** | | | | |
| --- | --- | --- | --- | --- |
| **Group** | **Product** | **Month 0** | **Month 1** | **Month 7** |
| AduFx | RTS,S/AS01_B_ | Full^1^ | Full | 1/5^th2^ |
| Adu1Fx | RTS,S/AS01_B_ | Full | - | 1/5^th^ |
| Adu2Fx | RTS,S/AS01_B_ | Full | 1/5^th^ | 1/5^th^ |
| PedFx | RTS,S/AS01_E_ | Full^3^ | Full | 1/5^th4^ |
| 2PedFx | RTS,S/AS01_E_ | Double^5^ | Double | Double 1/5^th6^ |
| Inf. Ctrl | none |  |  |  |
| **B. Vaccination schedule in the MAL102 trial (2)** | | | | |
| **Group** | **Product** | **Day 1**^7^ |  |  |
| P-Fx | RTS,S/AS01_E_ | Fractional^8^ |  |  |
| NP-Fx | RTS,S/AS01_E_ | Fractional^8^ |  |  |
| Inf. Ctrl |  |  |  |  |

**^1^** Full dose of the adult (Adu) formulation of RTS,S/AS01_B_: 50 µg of RTS,S, along with AS01_B_, an adjuvant system containing 50 µg of Monophosphoryl Lipid A, 50 µg of QS-21 (*Quillaja saponaria* Molina, fraction 21; licensed by GSK from Antigenics, a wholly owned subsisdiary of Agenus), and liposome in a 0.5-mL dose.

^2^1/5^th^ dose (Fx = fractional) of the adult formulation of RTS,S/AS01_B_: 10 µg of RTS,S containing 10 µg of Monophosphoryl Lipid A, 10 µg of QS-21, and liposome in a 0.1-mL dose.

^3^ Full dose of the pediatric (Ped) formulation of RTS,S/AS01_E_: 25 µg of RTS,S and an adjuvant system AS01_E_, (25 µg of MPL, QS-21, and liposomes) in a 0.5-mL dose.

^4^ 1/5^th^ dose of the pediatric formulation of RTS,S/AS01_E_: 5 µg of RTS,S and an adjuvant system AS01_E_, (5 µg of MPL, QS-21, and liposomes) in a 0.1-mL dose.

^5^ Double dose of the pediatric formulation of RTS,S/AS01_E_: 50 µg of RTS,S and an adjuvant system AS01_E_, (50 µg of MPL, QS-21, and liposomes) in a 1-mL dose.

^6^ Double 1/5^th^ dose of the pediatric formulation of RTS,S/AS01_E_: 10 µg of RTS,S and an adjuvant system AS01_E_, (10 µg of MPL, QS-21, and liposomes) in a 0.2-mL dose.

^7^ Day 1 = 12 months post-receipt of final vaccination in MAL092.

^8^ Fractional dose of RTS,S/AS01_E_: RTS,S/AS01_E_ contains 25 µg RTS,S and the AS01_E_ Adjuvant System, which consists of 25 µg MPL, 25 µg QS-21, and liposomes in a 0.5-mL dose. A fractional dosage of 0.1 mL was withdrawn from the reconstituted vaccine vial, and the administered dose was made up of 5 µg RTS,S in the AS01_E_ Adjuvant System (containing 5 µg MPL, 5 µg QS-21 and liposomes).

**Description of MAL092 groups:**

**AduFx:** Active Comparator. Healthy adult participants, between, and including, 18 and 55 years of age, who received RTS,S/AS01B full dose at Month 0 and Month 1 and 1/5th of RTS,S/AS01B full dose at Month 7, and underwent sporozoite challenge.

**Adu1Fx:** Active Comparator. Healthy adult participants, between, and including, 18 and 55 years of age, who received RTS,S/AS01B full dose at Month 0 and 1/5th of RTS,S/AS01B full dose administered at Month 7, and underwent sporozoite challenge.

**Adu2Fx:** Active Comparator. Healthy adult participants, between, and including, 18 and 55 years of age, who received RTS,S/AS01B full dose at Month 0 and 1/5th of RTS,S/AS01B full dose at Month 1 and Month 7, and underwent sporozoite challenge.

**2PedFx:** Experimental. Healthy adult participants, between, and including, 18 and 55 years of age, who received RTS,S/AS01E double dose at Month 0 and Month 1, and 1/5th of double dose RTS,S/AS01E at Month 7, and underwent sporozoite challenge.

**PedFx:** Experimental. Healthy adult participants, between, and including, 18 and 55 years of age, who received RTS,S/AS01E full dose at Month 0 and Month 1, and 1/5th of RTS,S/AS01E full dose at Month 7, and underwent sporozoite challenge.

**Inf. Ctrl (Infectivity Control):** Control group. Healthy participants, between and including 18 and 55 years of age, who did not receive any immunization but underwent sporozoite challenge.

**Description of MAL102 groups:**

**P-Fx:** Healthy adult participants, between and including 18 and 55 years of age, vaccinated with RTS,S/AS01 vaccine (different doses/formulations) and protected (P) following the first challenge at the time of the MAL092 study (NCT03162614), who received, in MAL102, a fractional (Fx) booster dose of RTS,S/AS01E 12 months after completion of the vaccination course in the MAL092 study (“Day 1” in table panel B), and underwent sporozoite challenge.

**NP-Fx:** Healthy adult participants, between and including 18 and 55 years of age, vaccinated with RTS,S/AS01 vaccine (different doses/formulations) and not protected (NP) following the first challenge at the time of the MAL092 study (NCT03162614), who received, in MAL102, a fractional (Fx) booster dose of RTS,S/AS01E 12 months after completion of the vaccination course in the Malaria-092 study, and underwent sporozoite challenge.

**Inf. Ctrl:** No intervention. Healthy adult participants, between and including 18 and 55 years of age, who did not receive, in MAL102, any immunization but underwent sporozoite challenge.

Supplementary Table 2. Numbers of participants, by study and biomarker, whose day-of-challenge biomarker levels were analyzed in this work.

| **Biomarker** | **Reporting Name** | **Laboratory** | **MAL068 (n = 46)** | **MAL071 (n = 45)** | **MAL092 (n = 94)** | **MAL102 (n = 36)** |
| --- | --- | --- | --- | --- | --- | --- |
| P falciparum CSP R32LR IgG Antibody^1^ | CSP IgG ELISA | Centre for Vaccinology (CEVAC), Ghent University, Belgium | 46 | 45 | 94 | 36 |
| IgG1 CSP WRAIR Avidity Index | IgG1 CSP AI^2^ | Georgia Tomaras (Duke University, Durham, NC, USA) | 32 | 45 | 91 | 36 |
| IgG3 CSP WRAIR Concentration | IgG3 CSP^2^ | Georgia Tomaras (Duke University, Durham, NC, USA) | 45 | 45 | 94 | 36 |
| IgG1 EP070034 NANP6 Concentration | IgG1 EP070034 NANP6^2^ | Georgia Tomaras (Duke University, Durham, NC, USA) | 43 | 40 | 94 | 36 |
| BLI NANP6 Dissociation AUC | BLI NANP6 Dissociation AUC^2^ | Georgia Tomaras (Duke University, Durham, NC, USA) | 46 | 45 | 94 | 36 |
| ADCP NANP6 | ADCP NANP6 | Galit Alter (SeromYx, Ragon Institute of MGH, MIT, and Harvard, Cambridge, MA, USA) | 45 | 45 | 93 | 36 |
| ADDCP CSP | ADDCP CSP | Galit Alter (SeromYx, Ragon Institute of MGH, MIT, and Harvard, Cambridge, MA, USA) | 45 | 45 | 94 | 36 |
| ADNKA MIP1b CSP | ADNKA MIP1b CSP | Galit Alter (SeromYx, Ragon Institute of MGH, MIT, and Harvard, Cambridge, MA, USA) | 45 | 45 | 94 | 36 |
| ADNP CSP | ADNP CSP | Galit Alter (SeromYx, Ragon Institute of MGH, MIT, and Harvard, Cambridge, MA, USA) | 45 | 45 | 94 | 36 |
| FCGR2AH NANP6^3^ | FCGR2AH NANP6 | Galit Alter (SeromYx, Ragon Institute of MGH, MIT, and Harvard, Cambridge, MA, USA) | 45 | 45 | 94 | 36 |
| FCGR2AR NANP6^3^ | FCGR2AR NANP6 | Galit Alter (SeromYx, Ragon Institute of MGH, MIT, and Harvard, Cambridge, MA, USA) | 45 | 45 | 94 | 36 |
| FCGR2B NANP6^3^ | FCGR2B NANP6 | Galit Alter (SeromYx, Ragon Institute of MGH, MIT, and Harvard, Cambridge, MA, USA) | 45 | 45 | 94 | 36 |
| FCGR3AF NANP6^3^ | FCGR3AF NANP6 | Galit Alter (SeromYx, Ragon Institute of MGH, MIT, and Harvard, Cambridge, MA, USA) | 45 | 45 | 94 | 34 |
| FCGR3AV NANP6^3^ | FCGR3AV NANP6 | Galit Alter (SeromYx, Ragon Institute of MGH, MIT, and Harvard, Cambridge, MA, USA) | 45 | 45 | 94 | 36 |
| FCGR3B NANP6^3^ | FCGR3B NANP6 | Galit Alter (SeromYx, Ragon Institute of MGH, MIT, and Harvard, Cambridge, MA, USA) | 45 | 45 | 91 | 36 |
| Total IgG NANP6 | Total IgG NANP6 | Galit Alter (SeromYx, Ragon Institute of MGH, MIT, and Harvard, Cambridge, MA, USA) | 45 | 45 | 94 | 36 |

^1^ R32LR is a recombinant protein consisting of two NVDP oligopeptides and 30 NANP repeats followed by LR [NVDP(NANP)_152_LR] (3, 4).

ADCP, antibody-dependent cellular phagocytosis; ADDCP, antibody dependent dendritic cell phagocytosis; ADNKA, antibody-dependent natural killer cell activation; ADNP, antibody-dependent neutrophil phagocytosis; BLI, bio-layer interferometry; CEVAC, Center for Vaccinology; CSP, circumsporozoite protein; ELISA, enzyme-linked immunosorbent assay; FCGR, Fc gamma receptor; MGH, Massachusetts General Hospital; WRAIR, Walter Reed Army Institute of Research.

^2^ Relating nomenclature here to nomenclature in Spreng et al. (5): IgG1 CSPAI = IgG1 CSP AI (same); IgG3 CSP = IgG3 CSP (same); IgG1 EP070034 NANP6 = IgG1 NANP6 in Spreng et al; BLI NANP6 Dissociation AUC = N Interface AUC_diss_ or serum NANP6 in Spreng et al.

^3^ Binding of NANP6-specific antibodies to: the FCGR2A H131 variant (FCGR2AH), the FCGR2A R131 variant (FCGR2AR), the FCGR2B receptor (FCGR2B NANP6), the FCGR3A F158 variant (FCGR3AF NANP6), the FCGR3A V158 variant (FCGR3AV NANP6), and the FCGR3B receptor (FCGR3B NANP6).

Supplementary Table 3. Summary of previous correlates results in Young et al. (6) for the 16 biomarkers included in the present analysis.

|  |  | **Figure panel in Young et al. supporting the biomarker as a candidate CoP^1,2^** | | | | | | | |
| --- | --- | --- | --- | --- | --- | --- | --- | --- | --- |
| **Kim et al. nomenclature (Young et al. nomenclature provided in parentheses if different)** | **Original Young et al. biomarker^3^** | **3A^4^**  **(068)** | **3C**  **(071)** | **5A**  **(068 RRR)** | | **5A (068 RRR)** | | **5C (068 Full)** | **5C**  **(068 Full)** |
|  |  |  |  | Uni | | Bi^5^ | | Uni | Bi^5^ |
| CSP IgG ELISA (ELISA Pfal CSP R32LR IgG) |  | * |  |  | |  | |  |  |
| IgG1 CSP AI (IgG1 CSP WRAIR Avidity Index) |  |  |  |  | |  | |  |  |
| IgG3 CSP | IgG3 CSP WRAIR magnitude (BAMA IgG3 CSP magnitude) |  |  |  | |  | |  |  |
| IgG1 EP070034 NANP6** | IgG1 EP070034 NANP6 magnitude (BAMA IgG1 NANP6 magnitude) | * |  |  | |  | |  |  |
| BLI NANP6 Dissociation AUC** | BLI NANP6 score (BLI Serum NANP6 score) |  |  |  |  | |  | |  |
| ADCP NANP6 |  | * |  |  | |  | |  |  |
| ADDCP CSP (ADDCP Phagocytosis Score CSP) |  |  |  |  | |  | |  |  |
| ADNKA MIP1b CSP |  | * |  |  | |  | |  |  |
| ADNP CSP (ADNP Phagocytosis Score CSP) |  |  |  |  | |  | |  |  |
| FCGR2AH NANP6 (Fc Array FcGRIIAH NANP6) |  | * |  |  | |  | |  |  |
| FCGR2AR NANP6 (Fc Array FcGRIIAR NANP6) |  | * |  |  | |  | |  |  |
| FCGR2B NANP6 (Fc Array FcGRIIB NANP6) |  |  |  |  | |  | |  |  |
| FCGR3AF NANP6 (Fc Array FcGRIIIAF NANP6) |  | * |  |  | |  | |  |  |
| FCGR3AV NANP6 (Fc Array FcGRIIIAV NANP6) |  | * |  |  | |  | |  |  |
| FCGR3B NANP6 (Fc Array FcGRIIIB NANP6) |  |  |  |  | |  | |  |  |
| Total IgG NANP6 (IgG Subclass IgG NANP6) |  |  |  |  | |  | |  |  |

^1^ Figure 3A and 3C of Young et al. reported the odds ratios for protection against patent parasitemia per unit increase in the designated immune response variable on the day of challenge, for the 20 most predictive immune response variables in each data set (A, MAL068; C, MAL071). All models were univariate and were done with (A) MAL068 (ARR + RRR) or (B) MAL071 (RRR + RRr) data.

^2^ Figure 5A and 5C of Young et al. reported the top 10 cross-validated AUC scores for the top-performing univariate and 2-variate models trained on (A) the MAL068 RRR data set or (C) the entire MAL068 data set.

^3^ In the present manuscript, IgG1 EP070034 NANP6 concentration, IgG3 CSP WRAIR concentration, and BLI NANP6 dissociation AUC were used in place of IgG1 EP070034 NANP6 magnitude, IgG3 CSP WRAIR magnitude, and BLI NANP6 score, respectively (see Methods). If the biomarker differs slightly between the first and second columns due to this substitution, the green cell shading in the right-hand columns pertains to the results for the original Young et al. biomarker (second column)

^4^ An asterisk indicates that the biomarker was significantly associated with protection (p-value < 0.2 after adjustment for false discovery rate).

^5^ The cell is shaded for "bi" if the variable appears in at least one top-ranking bivariate model.

ADCP, antibody-dependent cellular phagocytosis; ADDCP, antibody dependent dendritic cell phagocytosis; ADNKA, antibody-dependent natural killer cell activation; ADNP, antibody-dependent neutrophil phagocytosis; Adu, Adult; AI, avidity index; BLI, bio-layer interferometry; CSP, circumsporozoite protein; ELISA, enzyme-linked immunosorbent assay; Fx, fractional; IgG1, immunoglobulin G1; FCGR, Fc gamma receptor; IgG3, immunoglobulin G3; Ped, pediatric. Biomarker nomenclature: BLI NANP6: BLI NANP6 Dissociation AUC. ELISA: CSP IgG ELISA. Uni: Univariate model; Bi: bivariate model.

Supplementary Table 4. Vaccine efficacy and estimated protection rates against infection (*P. falciparum* parasitemia as assessed by blood smear after sporozoite challenge), by study and by arm, as well as for all arms combined in a study.

| **Trial (Reference)** | **Arm(s)** | **Vaccine Efficacy (VE), %**  **(95% Confidence Interval)^1^** | **Protection Efficacy (PE), %^2^** |
| --- | --- | --- | --- |
| MAL068 (7) | All | 47.8 (31.2, 60.4) | 47.8 |
|  | ARR | 44.0 (20.7, 60.4) | 44.0 |
|  | RRR | 52.4 (25.4, 69.6) | 52.4 |
| MAL071 (8) | All | 78.3 (62.4, 87.4) | 77.8 |
|  | RRr | 86.7 (66.8, 94.6) | 86.2 |
|  | RRR | 62.5 (29.4, 80.1) | 62.5 |
| MAL092 (1) | All | 55.8 (45.1, 64.3) | 56.4 |
|  | AduFx | 55.0 (26.9, 72.3) | 58.8 |
|  | Adu1Fx | 28.6 (6.4, 45.5) | 30.0 |
|  | Adu2Fx | 55.0 (26.9, 72.3) | 55.6 |
|  | PedFx | 63.6 (36.8, 79.1) | 63.2 |
|  | 2PedFx | 76.2 (48.8, 88.9) | 75.0 |
| MAL102 (2)^3^ | All | 53.1 (36.8, 65.1) | 52.8 |
|  | AduFx | 36.4 (0.5, 59.3) | 28.6 |
|  | Adu1Fx | 66.7 (25.8, 85.0) | 55.6 |
|  | Adu2Fx | 71.4 (7.8, 91.1) | 66.7 |
|  | PedFx | 41.7 (5.9, 63.8) | 50.0 |
|  | 2PedFx | 57.1 (-0.8, 81.8) | 66.7 |

^1^ Vaccine efficacy (VE) is defined as 100*(1-Relative Risk), as previously reported in the primary papers (1, 2, 7, 8). VE estimates are shown for comparison to the protection efficacy (PE) estimates used in the present work.

^2^ PE is defined as PE = 100*(nP /N), where nP is the number of protected participants and N is the total number of participants.

^3^ These numbers differ slightly from the previously reported VE estimates because out of the 130 participants originally enrolled in the MAL092 study, 104 of whom were in the per-protocol set and were used for the VE calculations in the primary paper (1), 94 participants who consented to have their samples used in subsequent investigations were included in the PE calculations for this analysis. Moreover, of the 49 participants who received a fractional RTS,S/AS01 booster 12 months post-primary regimen in the MAL102 study (2), n=36 participants who consented to have their samples used in subsequent investigations were included in the PE calculations for this analysis (5). See the “Study Samples” section in Methods.

RRR: two full doses one month apart; ARR: alternative arm with a modified version of the RRR regimen; RRr: two full doses one month apart, followed by a fractional fifth dose six months later.

Adu, adult; Fx, fractional; Ped, pediatric. See Supplementary Table 1 for details.

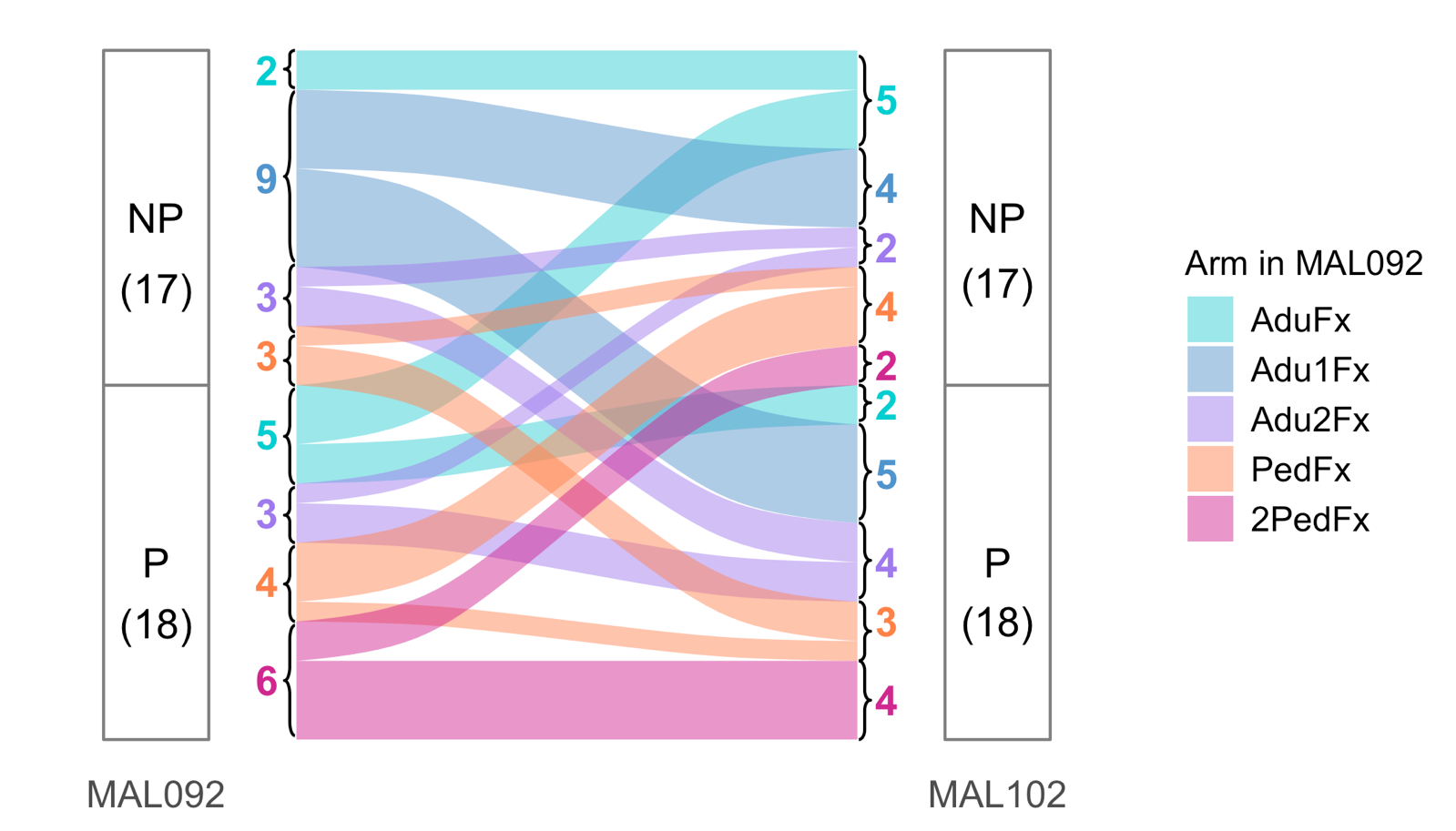

Supplementary Figure 1. Alluvial plot showing protection status in MAL092 and MAL102, restricted to the 35 participants from MAL092 who subsequently enrolled in MAL102. NP: not protected, P: protected. Numbers in parentheses and numbers by brackets represent numbers of participants. Adu, adult; Fx, fractional; Ped, pediatric. See Supplementary Table 1 for details on dose arms.

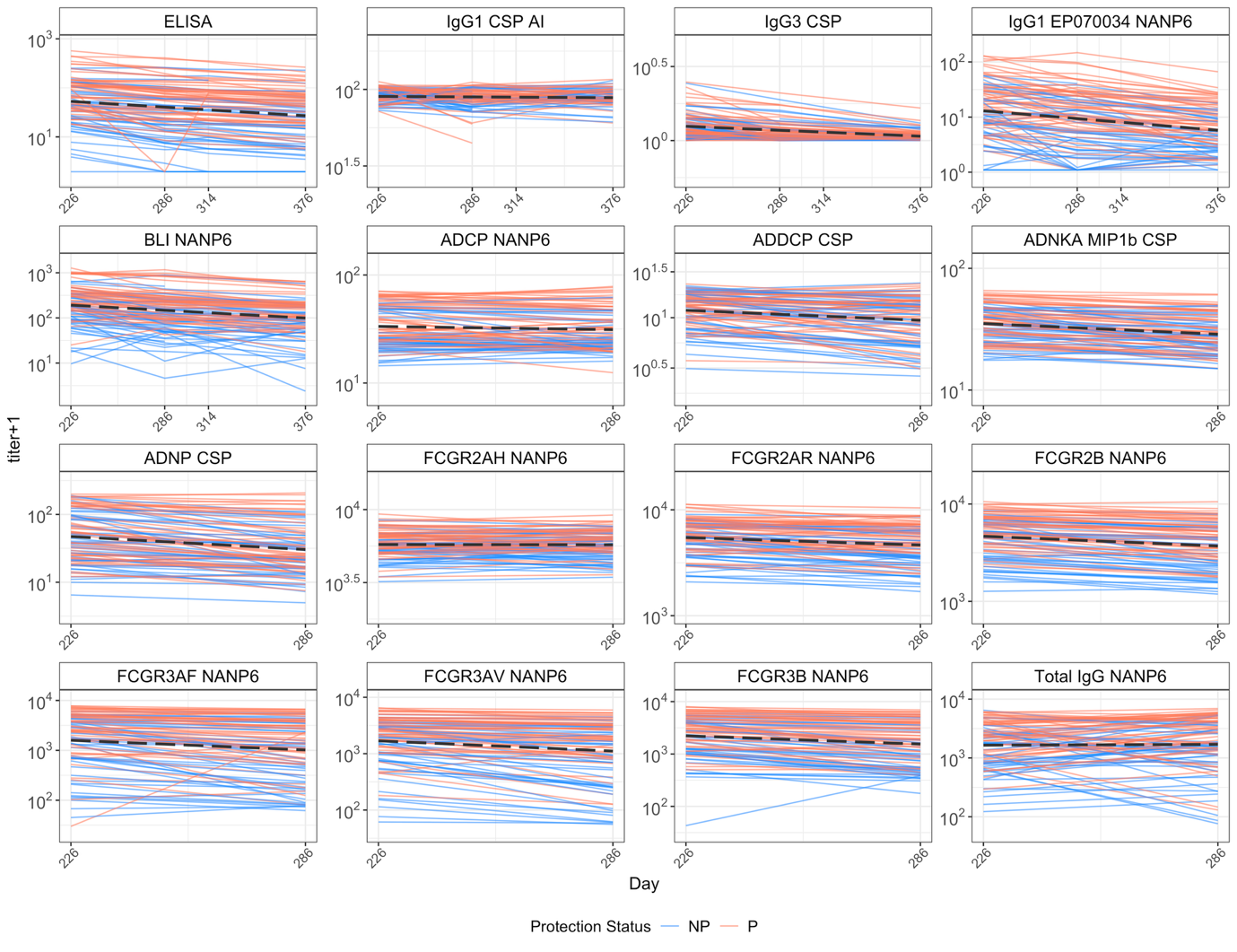

Supplementary Figure 2. Individual titer trajectories from Day 226 onward in MAL092 participants. Each line represents one participant, colored by protection status (blue = Not Protected, red = Protected). The gray line indicates the overall linear regression of titer (log scale) over time across all participants. Approximately linear trends are observed in individual trajectories, indicating log-linear decay after Day 226. ADCP, antibody-dependent cellular phagocytosis; ADDCP, antibody dependent dendritic cell phagocytosis; ADNKA, antibody-dependent natural killer cell activation; ADNP, antibody-dependent neutrophil phagocytosis; Adu, Adult; AI, avidity index; BLI, bio-layer interferometry; CSP, circumsporozoite protein; ELISA, enzyme-linked immunosorbent assay; Fx, fractional; IgG1, immunoglobulin G1; FCGR, Fc gamma receptor; IgG3, immunoglobulin G3; Ped, pediatric. Biomarker nomenclature: BLI NANP6: BLI NANP6 Dissociation AUC. ELISA: CSP IgG ELISA.

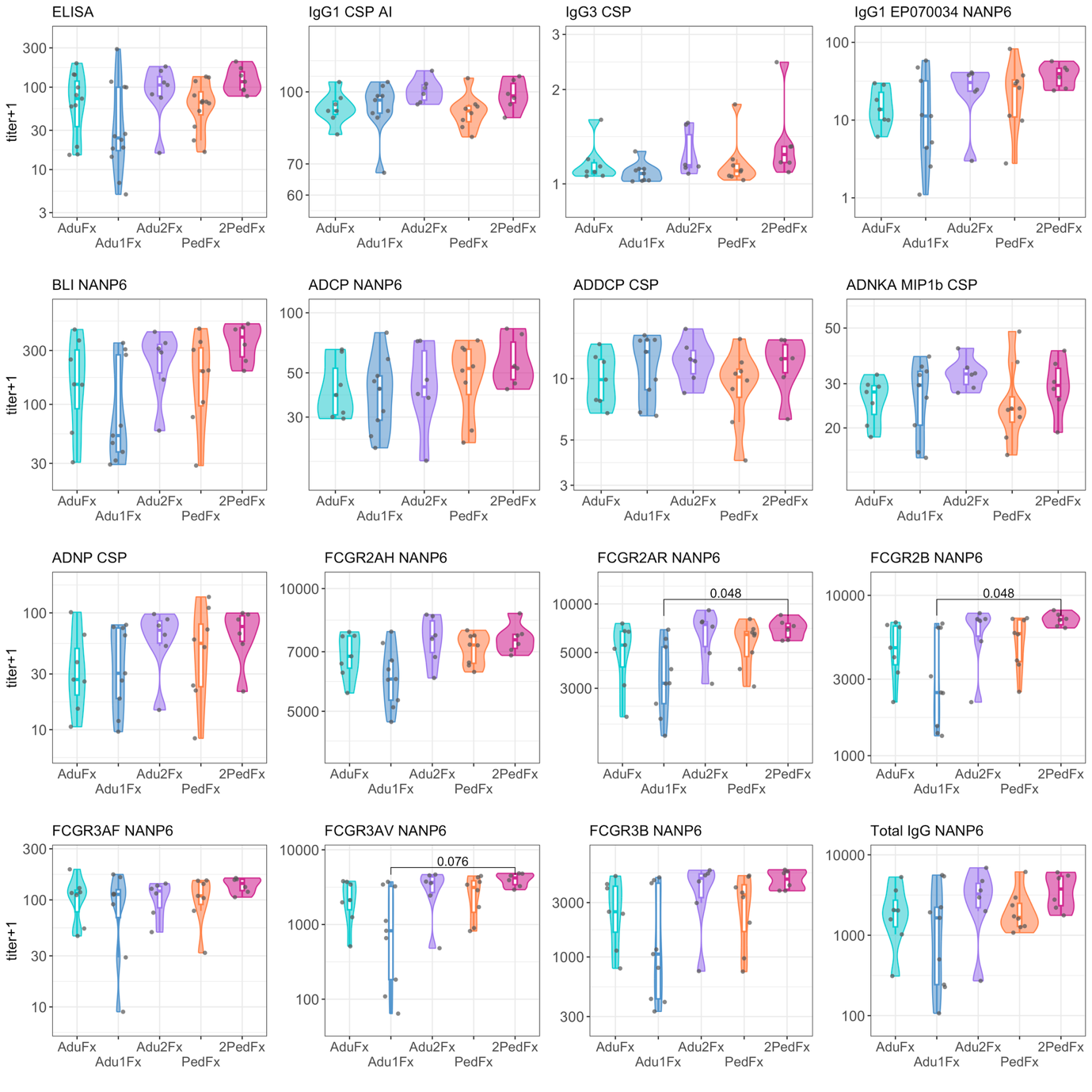
Supplementary Figure 3. Distribution of the 16 biomarkers on the day of challenge in MAL102, by study arm in MAL092. Each dot represents one study participant. In each plot, adjusted p-values from pairwise group comparisons (Wilcoxon rank sum test) are included when they were smaller than 0.10. P-value adjustment was carried out separately for each biomarker using the Benjamini-Hochberg procedure. ADCP, antibody-dependent cellular phagocytosis; ADDCP, antibody dependent dendritic cell phagocytosis; ADNKA, antibody-dependent natural killer cell activation; ADNP, antibody-dependent neutrophil phagocytosis; Adu, Adult; AI, avidity index; BLI, bio-layer interferometry; CSP, circumsporozoite protein; ELISA, enzyme-linked immunosorbent assay; Fx, fractional; IgG1, immunoglobulin G1; FCGR, Fc gamma receptor; IgG3, immunoglobulin G3; Ped, pediatric. Biomarker nomenclature: BLI NANP6: BLI NANP6 Dissociation AUC. ELISA: CSP IgG ELISA.

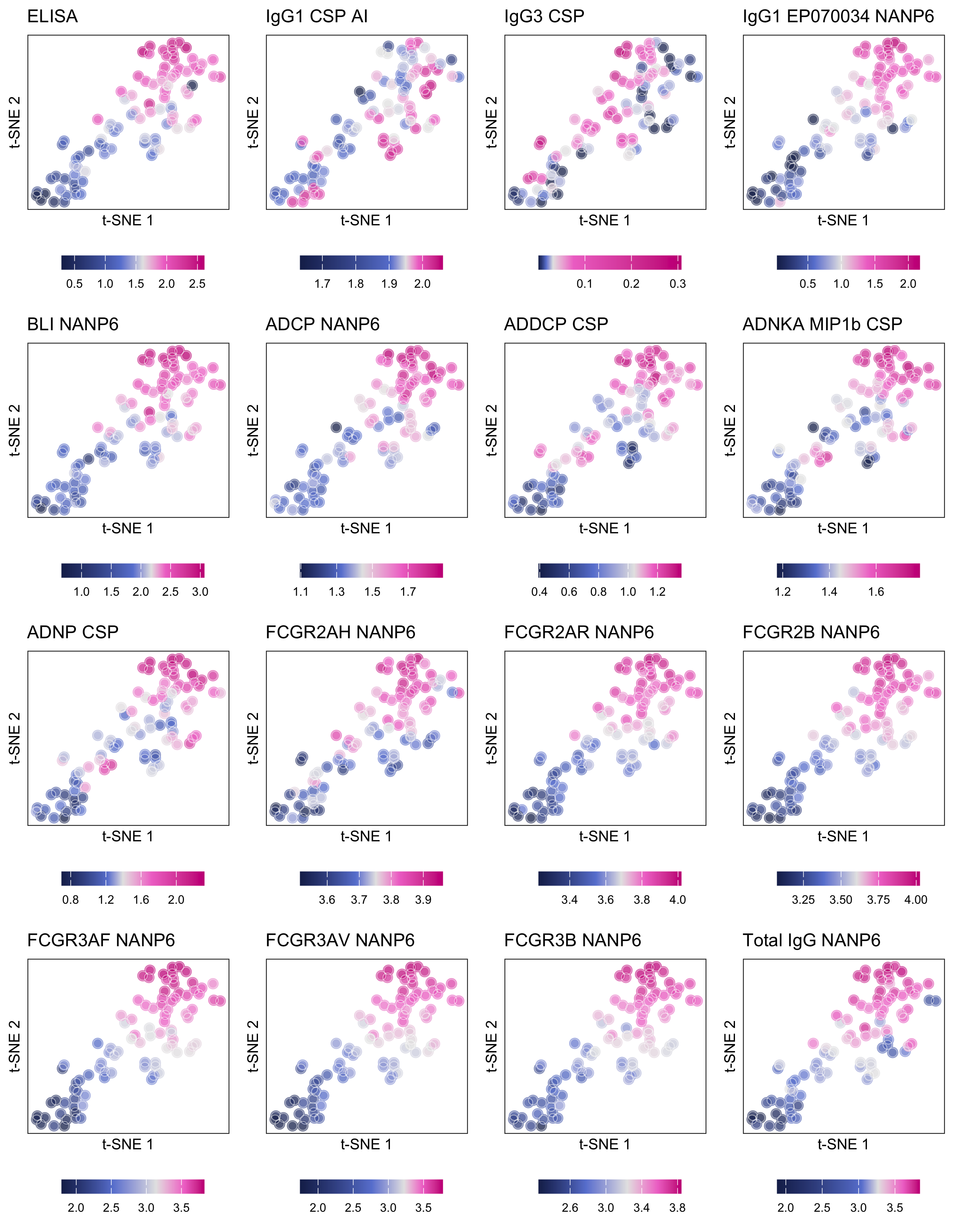

Supplementary Figure 4. *t*-SNE map displaying the level of the 16 biomarkers measured on the day of challenge in MAL092. Each dot represents one study participant from MAL092, and each of the 16 panels displays the level of one of the 16 biomarkers at the participant level using the color of the corresponding dots. Biomarker levels were normalized to center about 0 and have unit standard deviation (see plot-specific colorbar). ADCP, antibody-dependent cellular phagocytosis; ADDCP, antibody dependent dendritic cell phagocytosis; ADNKA, antibody-dependent natural killer cell activation; ADNP, antibody-dependent neutrophil phagocytosis; AI, avidity index; BLI, bio-layer interferometry; CSP, circumsporozoite protein; ELISA, enzyme-linked immunosorbent assay; IgG1, immunoglobulin G1; FCGR, Fc gamma receptor; IgG3, immunoglobulin G3. Biomarker nomenclature: BLI NANP6: BLI NANP6 Dissociation AUC. ELISA: CSP IgG ELISA.

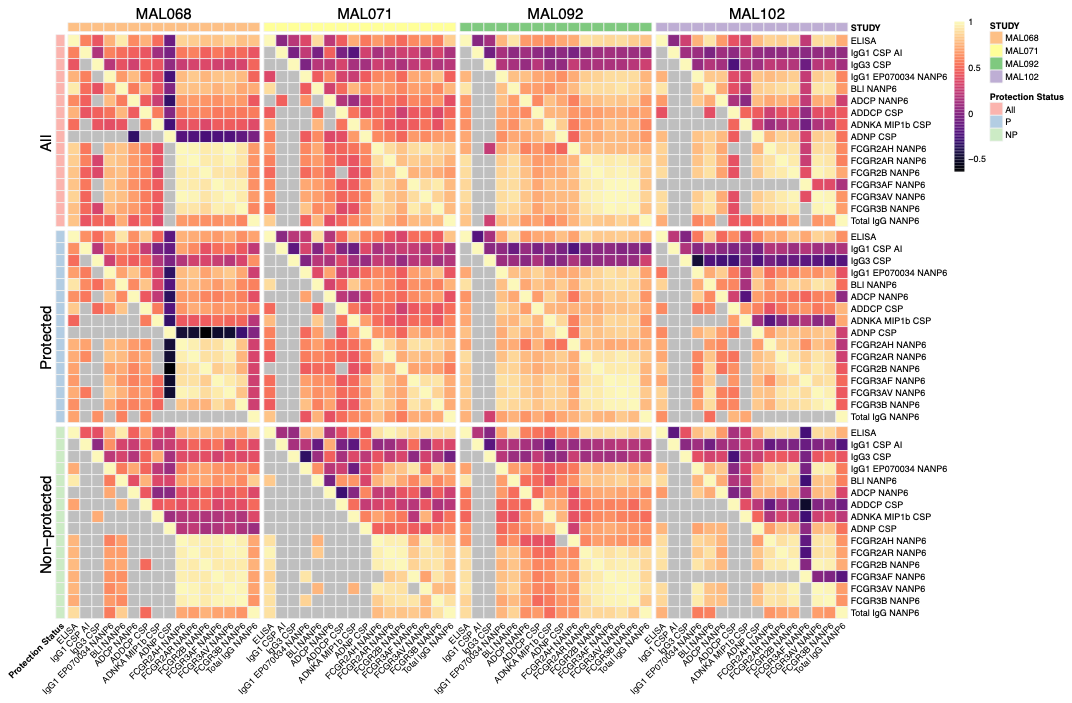

Supplementary Figure 5. Heatmaps of Spearman’s rank correlation coefficients measuring associations between day-of-challenge immune readouts. Each column represents one CHMI study (from left to right: MAL068, 071, 092, 102). The top row (All) shows correlation coefficients based on all participants from the corresponding study (i.e., regardless of protection status); the second and third rows show correlation coefficients computed using protected (second row) and non-protected (third row) study participants. Grey cells in the lower triangle of each heatmap indicate non-significant correlation coefficients (adjusted p > 0.05). The colors in the upper triangle represent the correlation coefficient values. For significant correlations, the lower triangle uses the same color as the corresponding upper triangle. If not significant, it remains grey in the lower triangle. ADCP, antibody-dependent cellular phagocytosis; ADDCP, antibody dependent dendritic cell phagocytosis; ADNKA, antibody-dependent natural killer cell activation; ADNP, antibody-dependent neutrophil phagocytosis; AI, avidity index; BLI, bio-layer interferometry; CSP, circumsporozoite protein; ELISA, enzyme-linked immunosorbent assay; IgG1, immunoglobulin G1; FCGR, Fc gamma receptor; IgG3, immunoglobulin G3. Biomarker nomenclature: BLI NANP6: BLI NANP6 Dissociation AUC. ELISA: CSP IgG ELISA.

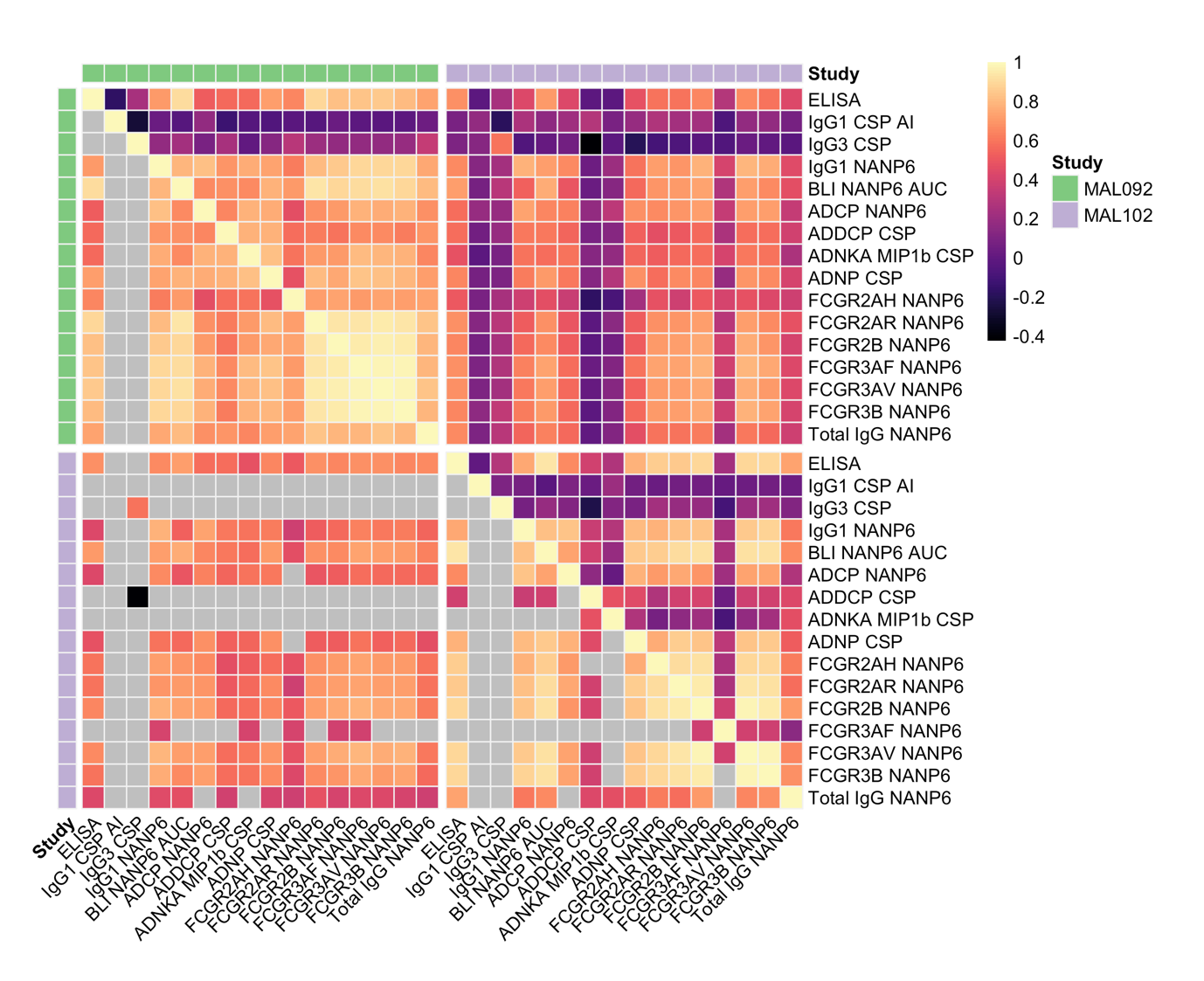

MAL092 Day 286

MAL102 Day 22

MAL102 Day 22

MAL092 Day 286

Day 22

Day 22

Supplementary Figure 6. Spearman’s correlation coefficients for biomarker levels at Day 286 (day of challenge) for MAL092 and Day 22 (day of challenge) for MAL102. Grey cells in the lower triangle of each heatmap indicate non-significant correlation coefficients (adjusted p > 0.05). The colors in the upper triangle represent the correlation coefficient values. For significant correlations, the lower triangle uses the same color as the corresponding upper triangle. If not significant, it remains grey in the lower triangle. ADCP, antibody-dependent cellular phagocytosis; ADDCP, antibody dependent dendritic cell phagocytosis; ADNKA, antibody-dependent natural killer cell activation; ADNP, antibody-dependent neutrophil phagocytosis; AI, avidity index; BLI, bio-layer interferometry; CSP, circumsporozoite protein; ELISA, enzyme-linked immunosorbent assay; IgG1, immunoglobulin G1; FCGR, Fc gamma receptor; IgG3, immunoglobulin G3. Biomarker nomenclature: BLI NANP6: BLI NANP6 Dissociation AUC. ELISA: CSP IgG ELISA.

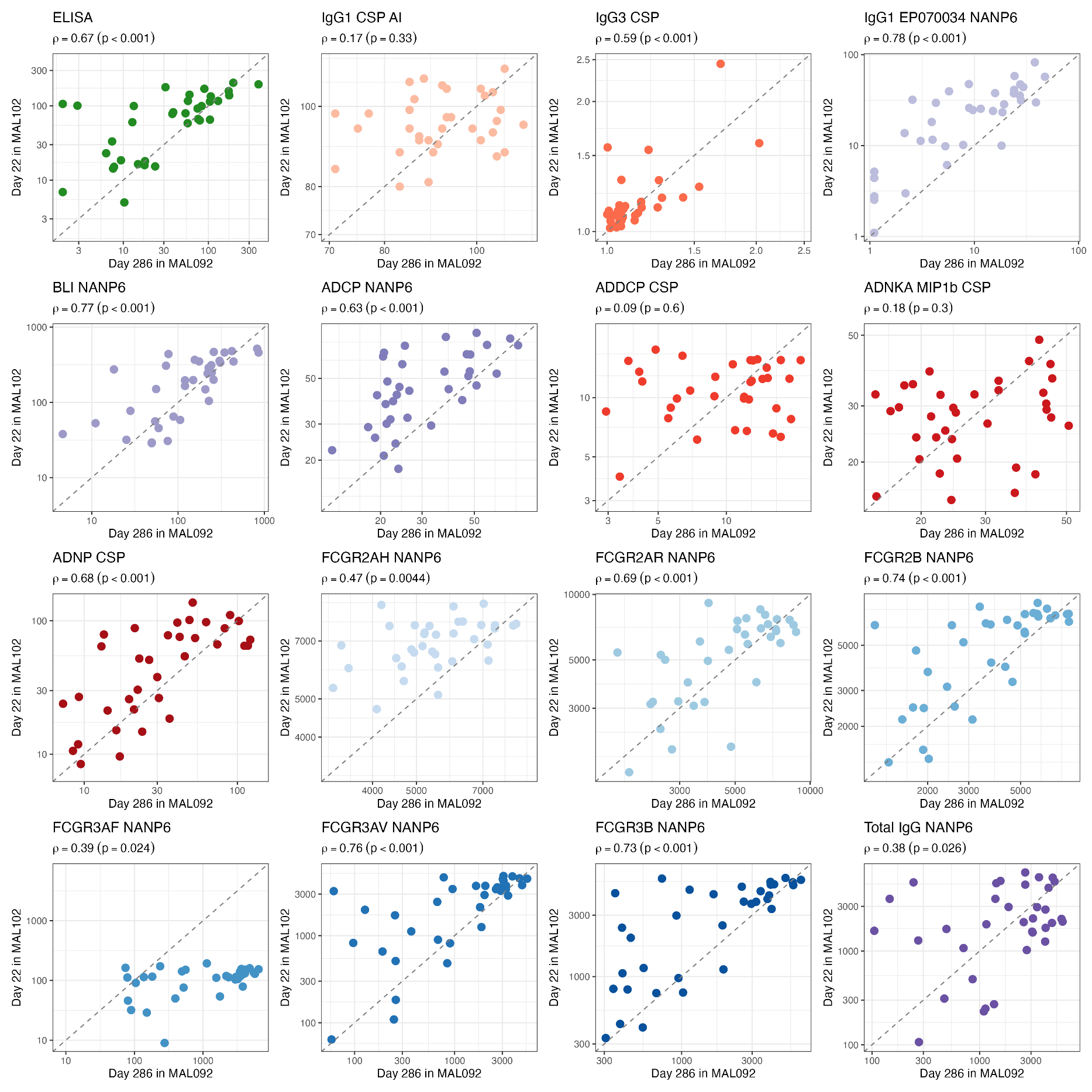

Supplementary Figure 7. Scatterplots of log-transformed biomarker levels at Day 22 in MAL102 versus at Day 286 in MAL092. The dashed diagonal line shows the line of identity (y=x). ADCP, antibody-dependent cellular phagocytosis; ADDCP, antibody dependent dendritic cell phagocytosis; ADNKA, antibody-dependent natural killer cell activation; ADNP, antibody-dependent neutrophil phagocytosis; AI, avidity index; BLI, bio-layer interferometry; CSP, circumsporozoite protein; ELISA, enzyme-linked immunosorbent assay; IgG1, immunoglobulin G1; FCGR, Fc gamma receptor; IgG3, immunoglobulin G3. Biomarker nomenclature: BLI NANP6: BLI NANP6 Dissociation AUC. ELISA: CSP IgG ELISA.

Supplementary Table 5. Counts and median fold-change from Day 226 (peak immunogenicity) to Day 286 (day of challenge) for each immune readout measured in the MAL092 study. Fold-change was calculated as the titer at Day 286 divided by the titer at Day 226 in MAL092 study (MAL092:Day 286/MAL092:Day 226). Related to Figure 3B.

|  | |  | Not Protected | | Protected | |
| --- | --- | --- | --- | --- | --- | --- |
| Biomarker | N | | n | Median  fold-change | n | Median  fold-change |
| CSP IgG ELISA | 94 | | 41 | 0.70 | 53 | 0.79 |
| IgG1 CSP Avidity Index | 75 | | 32 | 0.96 | 43 | 0.98 |
| IgG3 CSP | 91 | | 39 | 0.62 | 52 | 0.59 |
| IgG1 EP070034 NANP6 | 92 | | 40 | 0.58 | 52 | 0.75 |
| BLI NANP6 Dissociation AUC | 94 | | 41 | 0.66 | 53 | 0.76 |
| ADCP NANP6 | 92 | | 40 | 0.98 | 52 | 0.92 |
| ADDCP CSP | 94 | | 41 | 0.68 | 53 | 0.79 |
| ADNKA MIP1b CSP | 94 | | 41 | 0.78 | 53 | 0.83 |
| ADNP CSP | 94 | | 41 | 0.71 | 53 | 0.69 |
| FCGR2AH NANP6 | 94 | | 41 | 0.99 | 53 | 1.02 |
| FCGR2AR NANP6 | 94 | | 41 | 0.81 | 53 | 0.89 |
| FCGR2B NANP6 | 94 | | 41 | 0.78 | 53 | 0.84 |
| FCGR3AF NANP6 | 94 | | 41 | 0.63 | 53 | 0.80 |
| FCGR3AV NANP6 | 94 | | 41 | 0.61 | 53 | 0.83 |
| FCGR3B NANP6 | 90 | | 38 | 0.62 | 52 | 0.82 |
| Total IgG NANP6 | 94 | | 41 | 1.04 | 53 | 1.42 |

ADCP, antibody-dependent cellular phagocytosis; ADDCP, antibody dependent dendritic cell phagocytosis; ADNKA, antibody-dependent natural killer cell activation; ADNP, antibody-dependent neutrophil phagocytosis; AI, avidity index; BLI, bio-layer interferometry; CSP, circumsporozoite protein; ELISA, enzyme-linked immunosorbent assay; IgG1, immunoglobulin G1; FCGR, Fc gamma receptor; IgG3, immunoglobulin G3.

Supplementary Table 6. Counts and median fold-change from Day 286 (day of challenge in MAL092 study) to Day 22 (day of challenge in MAL102 study). Fold-change was calculated as the titer at Day 22 in MAL102 divided by the titer at Day 286 in MAL092 (fold-change = MAL102:Day 22/MAL092:Day 286). Column headers denote protection status at **(MAL092, MAL102)** respectively, where **P** = Protected and **NP** = Not Protected. For example, (NP_092_, NP_102_) indicates participants who were not protected in both MAL092 and MAL102 studies. n=number of participants; FC= fold-change. Related to Figure 3C.

|  | (NP_092_, NP_102_) | | (NP_092_, P_102_) | | (P_092_, NP_102_) | | (P_092_, P_102_) | |
| --- | --- | --- | --- | --- | --- | --- | --- | --- |
| Biomarker | n | Median  FC | n | Median  FC | n | Median  FC | n | Median  FC |
| CSP IgG ELISA | 8 | 1.59 | 9 | 2.08 | 9 | 0.88 | 9 | 1.10 |
| IgG1 CSP Avidity Index | 7 | 1.04 | 9 | 1.00 | 9 | 1.06 | 9 | 1.03 |
| IgG3 CSP | 8 | 1.47 | 9 | 0.86 | 9 | 0.87 | 9 | 2.55 |
| IgG1 EP070034 NANP6 | 8 | 3.98 | 9 | 4.99 | 9 | 1.51 | 9 | 1.63 |
| BLI NANP6 Dissociation AUC | 8 | 0.88 | 9 | 2.82 | 9 | 0.77 | 9 | 1.18 |
| ADCP NANP6 | 8 | 1.41 | 9 | 1.93 | 9 | 1.42 | 9 | 1.14 |
| ADDCP CSP | 8 | 1.47 | 9 | 0.88 | 9 | 0.80 | 9 | 0.93 |
| ADNKA MIP1b CSP | 8 | 1.09 | 9 | 1.15 | 9 | 0.81 | 9 | 1.05 |
| ADNP CSP | 8 | 1.31 | 9 | 1.41 | 9 | 0.97 | 9 | 1.53 |
| FCGR2AH NANP6 | 8 | 1.20 | 9 | 1.40 | 9 | 1.04 | 9 | 1.24 |
| FCGR2AR NANP6 | 8 | 0.93 | 9 | 1.36 | 9 | 0.92 | 9 | 1.08 |
| FCGR2B NANP6 | 8 | 0.97 | 9 | 1.52 | 9 | 1.00 | 9 | 1.21 |
| FCGR3AF NANP6 | 8 | 0.45 | 8 | 0.17 | 8 | 0.03 | 9 | 0.03 |
| FCGR3AV NANP6 | 8 | 1.21 | 9 | 3.47 | 9 | 0.91 | 9 | 1.27 |
| FCGR3B NANP6 | 8 | 1.10 | 8 | 3.25 | 9 | 1.09 | 9 | 1.29 |
| Total IgG NANP6 | 8 | 0.53 | 9 | 1.67 | 9 | 0.51 | 9 | 1.11 |

ADCP, antibody-dependent cellular phagocytosis; ADDCP, antibody dependent dendritic cell phagocytosis; ADNKA, antibody-dependent natural killer cell activation; ADNP, antibody-dependent neutrophil phagocytosis; AI, avidity index; BLI, bio-layer interferometry; CSP, circumsporozoite protein; ELISA, enzyme-linked immunosorbent assay; IgG1, immunoglobulin G1; FCGR, Fc gamma receptor; IgG3, immunoglobulin G3.

Supplementary Table 7. Results from linear regression analyses modeling fold-change, defined as the ratio of the Day 286 (day of challenge) and Day 226 (peak immunogenicity) titers [Y(286)/Y(226)], as a function of protection status. The first column (median FC) shows the median fold-change for each biomarker, regardless of protection status. Median fold-change of one indicates no decay in antibody levels between Day 226 and Day 286. A positive coefficient for protection status indicates a slower decay between Day 226 and Day 286 in protected individuals relative to non-protected individuals.

|  |  | **Intercept** | | | **Protection Status (Reference = NP)** | | |
| --- | --- | --- | --- | --- | --- | --- | --- |
| **Biomarker** | **Median Fold-Change** | **Intercept** | **P-value** | **Adj. P-value** | **Coefficient** | **P-value** | **Adj. P-value** |
| CSP IgG ELISA | 0.72 | 0.65 | <0.001 | <0.001 | 0.108 | **0.024** | 0.126 |
| IgG1 CSP AI | 0.97 | 0.96 | <0.001 | <0.001 | -0.004 | 0.913 | 0.913 |
| IgG3 CSP | 0.60 | 4.74 | 0.146 | 0.156 | -0.870 | 0.839 | 0.913 |
| IgG1 EP070034 NANP6 | 0.63 | 1.46 | 0.008 | 0.009 | -0.667 | 0.352 | 0.699 |
| BLI NANP6 Dissociation AUC | 0.73 | 0.83 | <0.001 | <0.001 | -0.020 | 0.876 | 0.913 |
| ADCP NANP6 | 0.95 | 0.98 | <0.001 | <0.001 | -0.047 | 0.327 | 0.699 |
| ADDCP CSP | 0.75 | 0.73 | <0.001 | <0.001 | 0.164 | **0.016** | 0.126 |
| ADNKA MIP1b CSP | 0.82 | 0.82 | <0.001 | <0.001 | 0.008 | 0.811 | 0.913 |
| ADNP CSP | 0.69 | 0.68 | <0.001 | <0.001 | 0.012 | 0.822 | 0.913 |
| FCGR2AH NANP6 | 1.00 | 0.99 | <0.001 | <0.001 | 0.026 | 0.393 | 0.699 |
| FCGR2AR NANP6 | 0.84 | 0.82 | <0.001 | <0.001 | 0.074 | **0.052** | 0.166 |
| FCGR2B NANP6 | 0.82 | 0.77 | <0.001 | <0.001 | 0.062 | **0.048** | 0.166 |
| FCGR3AF NANP6 | 0.70 | 0.62 | 0.628 | 0.628 | 1.642 | 0.338 | 0.699 |
| FCGR3AV NANP6 | 0.77 | 0.61 | <0.001 | <0.001 | 0.229 | **0.003** | 0.053 |
| FCGR3B NANP6 | 0.75 | 0.82 | <0.001 | <0.001 | -0.026 | 0.897 | 0.913 |
| Total IgG NANP6 | 1.26 | 1.49 | <0.001 | <0.001 | -0.071 | 0.762 | 0.913 |

ADCP, antibody-dependent cellular phagocytosis; ADDCP, antibody dependent dendritic cell phagocytosis; ADNKA, antibody-dependent natural killer cell activation; ADNP, antibody-dependent neutrophil phagocytosis; AI, avidity index; BLI, bio-layer interferometry; CSP, circumsporozoite protein; ELISA, enzyme-linked immunosorbent assay; IgG1, immunoglobulin G1; FCGR, Fc gamma receptor; IgG3, immunoglobulin G3.

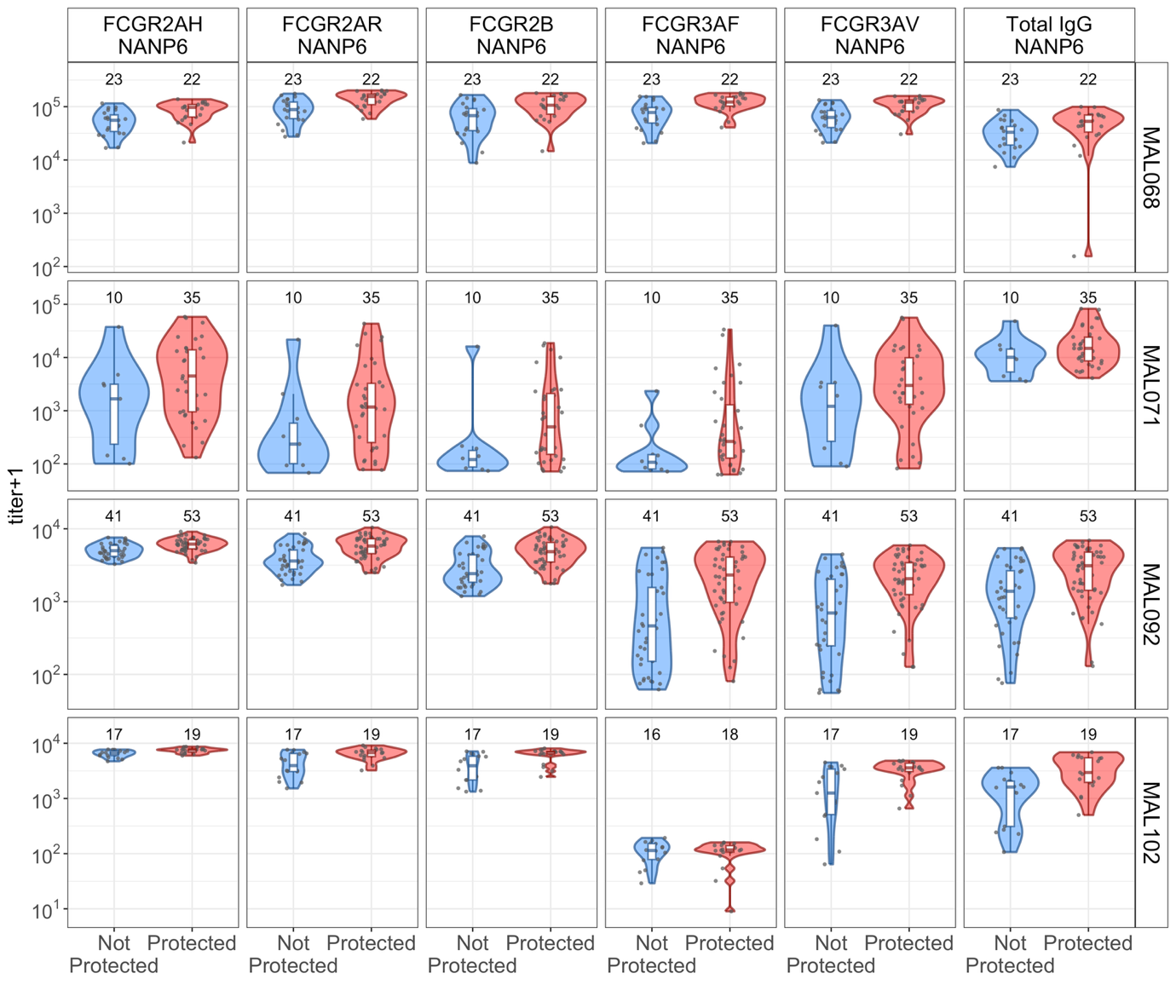
Supplementary Figure 8. Distributions of day-of-challenge levels of six of the remaining biomarkers not shown in Figure 5 (the other six are shown in Supplementary Figure 9), in MAL068, MAL071, MAL092 and MAL102, shown separately by protection status. Blue = not protected; red = protected. Each dot indicates one study participant. The y-axis of each plot represents the value of the measured titer. FCGR, Fc gamma receptor; IgG, immunoglobulin.

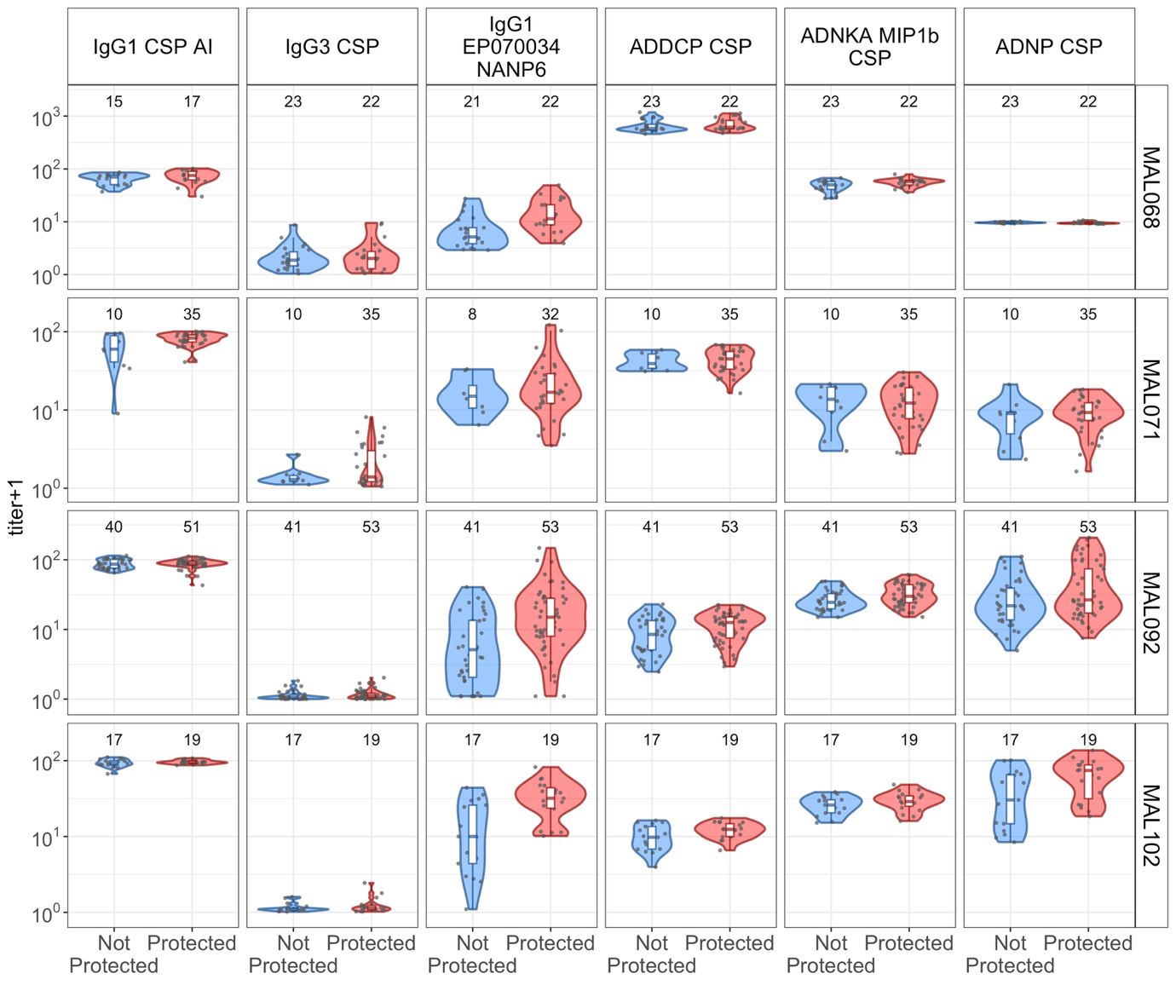
Supplementary Figure 9. Distributions of day-of-challenge levels of six of the remaining biomarkers not shown in Figure 5 (the other six are shown in Supplementary Figure 8), in MAL068, MAL071, MAL092 and MAL102, shown separately by protection status. Blue = not protected; red = protected. Each dot indicates one study participant. The y-axis of each plot represents the value of the measured titer. ADDCP, antibody dependent dendritic cell phagocytosis; ADNKA, antibody-dependent natural killer cell activation; ADNP, antibody-dependent neutrophil phagocytosis; AI, avidity index; CSP, circumsporozoite protein; IgG1, immunoglobulin G1; FCGR, Fc gamma receptor; IgG3, immunoglobulin G3.

| MAL068 | Not Protected | | | | Protected | | | |
| --- | --- | --- | --- | --- | --- | --- | --- | --- |
| Biomarker | **n** | **Median** | **Geometric mean** | **Geometric SD** | **n** | **Median** | **Geometric mean** | **Geometric SD** |
| CSP IgG ELISA | 24 | 50.2 | 51.8 | 2.2 | 22 | 116.9 | 120.8 | 2.4 |
| IgG1 CSP AI | 15 | 69.0 | 62.4 | 1.3 | 17 | 74.0 | 68.5 | 1.4 |
| IgG3 CSP | 23 | 0.9 | 0.8 | 3.1 | 22 | 1.0 | 0.8 | 4.0 |
| IgG1 EP070034 NANP6 | 21 | 4.2 | 5.1 | 2.1 | 22 | 10.4 | 11.3 | 2.1 |
| BLI NANP6 Dissociation AUC | 24 | 124.6 | 95.2 | 3.5 | 22 | 210.2 | 208.4 | 4.0 |
| ADCP NANP6 | 23 | 7.8 | 7.9 | 1.1 | 22 | 9.2 | 9.5 | 1.3 |
| ADDCP CSP | 23 | 583.0 | 645.0 | 1.3 | 22 | 615.0 | 686.8 | 1.3 |
| ADNKA MIP1b CSP | 23 | 49.1 | 45.9 | 1.3 | 22 | 56.6 | 54.2 | 1.2 |
| ADNP CSP | 23 | 8.6 | 8.6 | 1.0 | 22 | 8.4 | 8.6 | 1.1 |
| FCGR2AH NANP6 | 23 | 55620.0 | 49411.6 | 1.7 | 22 | 97120.0 | 82011.7 | 1.5 |
| FCGR2AR NANP6 | 23 | 88187.0 | 81795.0 | 1.7 | 22 | 154855.5 | 136237.6 | 1.4 |
| FCGR2B NANP6 | 23 | 67844.0 | 55609.7 | 2.1 | 22 | 106583.0 | 99116.8 | 1.8 |
| FCGR3AF NANP6 | 23 | 77746.0 | 68587.6 | 1.8 | 22 | 122541.0 | 116227.7 | 1.5 |
| FCGR3AV NANP6 | 23 | 62944.0 | 59819.9 | 1.7 | 22 | 121337.0 | 102947.0 | 1.5 |
| FCGR3B NANP6 | 23 | 57999.0 | 51838.7 | 2.1 | 22 | 111028.5 | 95470.2 | 1.6 |
| Total IgG NANP6 | 23 | 33087.0 | 29284.3 | 1.8 | 22 | 52955.0 | 37186.5 | 3.8 |

Supplementary Table 8. Median, geometric mean titer (GMT) and geometric standard deviation (GSD) of antibody titers on the day of challenge in MAL068 by biomarker and protection status.

ADCP, antibody-dependent cellular phagocytosis; ADDCP, antibody dependent dendritic cell phagocytosis; ADNKA, antibody-dependent natural killer cell activation; ADNP, antibody-dependent neutrophil phagocytosis; Adu, Adult; AI, avidity index; BLI, bio-layer interferometry; CSP, circumsporozoite protein; ELISA, enzyme-linked immunosorbent assay; Fx, fractional; IgG1, immunoglobulin G1; FCGR, Fc gamma receptor; IgG3, immunoglobulin G3; Ped, pediatric. Biomarker nomenclature: BLI NANP6: BLI NANP6 Dissociation AUC. ELISA: CSP IgG ELISA.

| MAL071 | Not protected | | | | Protected | | | |
| --- | --- | --- | --- | --- | --- | --- | --- | --- |
| Biomarker | **n** | **Median** | **Geometric mean** | **Geometric SD** | **n** | **Median** | **Geometric mean** | **Geometric SD** |
| CSP IgG ELISA | 10 | 67.1 | 57.8 | 2.5 | 35 | 105.0 | 90.8 | 2.0 |
| IgG1 CSP AI | 10 | 59.0 | 50.8 | 2.1 | 35 | 82.0 | 78.3 | 1.2 |
| IgG3 CSP | 10 | 0.3 | 0.3 | 2.1 | 35 | 0.4 | 0.6 | 3.8 |
| IgG1 EP070034 NANP6 | 8 | 14.1 | 14.0 | 1.8 | 32 | 15.9 | 16.9 | 2.4 |
| BLI NANP6 Dissociation AUC | 10 | 224.5 | 193.0 | 2.5 | 35 | 290.3 | 292.2 | 1.7 |
| ADCP NANP6 | 10 | 14.5 | 15.9 | 1.3 | 35 | 20.2 | 20.2 | 1.5 |
| ADDCP CSP | 10 | 38.5 | 40.3 | 1.3 | 35 | 44.0 | 40.7 | 1.4 |
| ADNKA MIP1b CSP | 10 | 12.7 | 10.0 | 2.2 | 35 | 11.3 | 9.8 | 2.1 |
| ADNP CSP | 10 | 8.0 | 5.9 | 2.3 | 35 | 8.3 | 7.2 | 2.0 |
| FCGR2AH NANP6 | 10 | 1670.5 | 1240.7 | 6.5 | 35 | 4487.0 | 3842.1 | 5.2 |
| FCGR2AR NANP6 | 10 | 235.0 | 364.3 | 5.9 | 35 | 1175.0 | 1239.9 | 6.3 |
| FCGR2B NANP6 | 10 | 120.0 | 190.8 | 5.0 | 35 | 496.0 | 620.5 | 5.5 |
| FCGR3AF NANP6 | 10 | 106.5 | 159.4 | 3.0 | 35 | 264.0 | 457.1 | 5.8 |
| FCGR3AV NANP6 | 10 | 1339.0 | 1079.8 | 6.6 | 35 | 2986.0 | 2820.4 | 5.5 |
| FCGR3B NANP6 | 10 | 1003.0 | 955.8 | 7.5 | 35 | 4189.0 | 3170.4 | 7.5 |
| Total IgG NANP6 | 10 | 10096.0 | 9991.7 | 2.2 | 35 | 14857.0 | 15614.3 | 2.2 |

Supplementary Table 9. Median, geometric mean titer (GMT) and geometric standard deviation (GSD) of antibody titers on the day of challenge in MAL071 by biomarker and protection status.

ADCP, antibody-dependent cellular phagocytosis; ADDCP, antibody dependent dendritic cell phagocytosis; ADNKA, antibody-dependent natural killer cell activation; ADNP, antibody-dependent neutrophil phagocytosis; Adu, Adult; AI, avidity index; BLI, bio-layer interferometry; CSP, circumsporozoite protein; ELISA, enzyme-linked immunosorbent assay; Fx, fractional; IgG1, immunoglobulin G1; FCGR, Fc gamma receptor; IgG3, immunoglobulin G3; Ped, pediatric. Biomarker nomenclature: BLI NANP6: BLI NANP6 Dissociation AUC. ELISA: CSP IgG ELISA.

| MAL092 | Not Protected | | | | Protected | | | |
| --- | --- | --- | --- | --- | --- | --- | --- | --- |
| Biomarker | **n** | **Median** | **Geometric mean** | **Geometric SD** | **n** | **Median** | **Geometric mean** | **Geometric SD** |
| CSP IgG ELISA | 41 | 19.0 | 18.9 | 4.0 | 53 | 65.7 | 55.9 | 3.0 |
| IgG1 CSP AI | 40 | 85.5 | 85.4 | 1.2 | 51 | 88.0 | 86.6 | 1.2 |
| IgG3 CSP | 41 | 0.07 | 0.04 | 6.8 | 53 | 0.1 | 0.07 | 6.0 |
| IgG1 EP070034 NANP6 | 41 | 4.1 | 2.7 | 6.7 | 53 | 14.0 | 11.4 | 4.2 |
| BLI NANP6 Dissociation AUC | 41 | 76.4 | 81.1 | 3.0 | 53 | 218.8 | 207.1 | 2.2 |
| ADCP NANP6 | 40 | 22.8 | 25.5 | 1.4 | 53 | 32.1 | 34.2 | 1.6 |
| ADDCP CSP | 41 | 7.5 | 6.8 | 2.0 | 53 | 11.6 | 9.4 | 1.8 |
| ADNKA MIP1b CSP | 41 | 23.5 | 24.8 | 1.4 | 53 | 29.0 | 30.1 | 1.4 |
| ADNP CSP | 41 | 20.9 | 23.3 | 2.2 | 53 | 25.6 | 34.4 | 2.5 |
| FCGR2AH NANP6 | 41 | 5000.0 | 5032.8 | 1.2 | 53 | 6158.0 | 6067.0 | 1.2 |
| FCGR2AR NANP6 | 41 | 3613.5 | 3704.0 | 1.5 | 53 | 5802.0 | 5522.8 | 1.4 |
| FCGR2B NANP6 | 41 | 2422.0 | 2751.7 | 1.7 | 53 | 4850.5 | 4657.0 | 1.6 |
| FCGR3AF NANP6 | 41 | 463.5 | 493.7 | 4.2 | 53 | 2322.0 | 1774.0 | 3.1 |
| FCGR3AV NANP6 | 41 | 697.5 | 573.4 | 3.9 | 53 | 2060.5 | 1846.1 | 2.5 |
| FCGR3B NANP6 | 38 | 743.8 | 877.0 | 2.5 | 53 | 2633.0 | 2310.2 | 2.2 |
| Total IgG NANP6 | 41 | 1386.0 | 1109.4 | 3.1 | 53 | 3110.0 | 2359.3 | 2.5 |

Supplementary Table 10. Median, geometric mean titer (GMT) and geometric standard deviation (GSD) of antibody titers on the day of challenge in MAL092 by biomarker and protection status.

ADCP, antibody-dependent cellular phagocytosis; ADDCP, antibody dependent dendritic cell phagocytosis; ADNKA, antibody-dependent natural killer cell activation; ADNP, antibody-dependent neutrophil phagocytosis; Adu, Adult; AI, avidity index; BLI, bio-layer interferometry; CSP, circumsporozoite protein; ELISA, enzyme-linked immunosorbent assay; Fx, fractional; IgG1, immunoglobulin G1; FCGR, Fc gamma receptor; IgG3, immunoglobulin G3; Ped, pediatric. Biomarker nomenclature: BLI NANP6: BLI NANP6 Dissociation AUC. ELISA: CSP IgG ELISA.

Supplementary Table 11. Median, geometric mean titer (GMT) and geometric standard deviation (GSD) of antibody titers on the day of challenge in MAL102 by biomarker and protection status.

| MAL102 | Not protected | | | | Protected | | | |
| --- | --- | --- | --- | --- | --- | --- | --- | --- |
| Biomarker | **n** | **Median** | **Geometric mean** | **Geometric SD** | **n** | **Median** | **Geometric mean** | **Geometric SD** |
| CSP IgG ELISA | 17 | 59.3 | 35.5 | 3.2 | 19 | 105.3 | 85.7 | 2.1 |
| IgG1 CSP AI | 17 | 91.0 | 91.2 | 1.1 | 19 | 94.0 | 95.0 | 1.1 |
| IgG3 CSP | 17 | 0.1 | 0.1 | 2.5 | 19 | 0.1 | 0.1 | 3.0 |
| IgG1 EP070034 NANP6 | 17 | 9.0 | 7.3 | 4.8 | 19 | 30.9 | 28.2 | 1.9 |
| BLI NANP6 Dissociation AUC | 17 | 103.6 | 100.6 | 2.8 | 19 | 306.3 | 241.7 | 2.0 |
| ADCP NANP6 | 17 | 30.6 | 36.2 | 1.6 | 19 | 45.1 | 50.7 | 1.3 |
| ADDCP CSP | 17 | 8.8 | 8.3 | 1.6 | 19 | 11.4 | 10.9 | 1.3 |
| ADNKA MIP1b CSP | 17 | 25.0 | 24.5 | 1.3 | 19 | 27.9 | 27.9 | 1.4 |
| ADNP CSP | 17 | 29.4 | 29.5 | 2.5 | 19 | 73.2 | 55.2 | 1.9 |
| FCGR2AH NANP6 | 17 | 6580.5 | 6393.6 | 1.2 | 19 | 7638.5 | 7400.9 | 1.1 |
| FCGR2AR NANP6 | 17 | 3950.0 | 3987.5 | 1.7 | 19 | 6787.5 | 6351.0 | 1.3 |
| FCGR2B NANP6 | 17 | 3922.0 | 3499.8 | 1.8 | 19 | 6785.0 | 5938.5 | 1.4 |
| FCGR3AF NANP6 | 16 | 113.0 | 100.5 | 1.7 | 18 | 115.5 | 94.1 | 2.1 |
| FCGR3AV NANP6 | 17 | 1252.0 | 1060.4 | 3.8 | 19 | 3581.5 | 3058.2 | 1.7 |
| FCGR3B NANP6 | 17 | 1135.5 | 1530.6 | 2.6 | 19 | 4604.5 | 3727.6 | 1.7 |
| Total IgG NANP6 | 17 | 1610.0 | 1043.5 | 3.0 | 19 | 2951.0 | 2950.9 | 2.1 |

ADCP, antibody-dependent cellular phagocytosis; ADDCP, antibody dependent dendritic cell phagocytosis; ADNKA, antibody-dependent natural killer cell activation; ADNP, antibody-dependent neutrophil phagocytosis; Adu, Adult; AI, avidity index; BLI, bio-layer interferometry; CSP, circumsporozoite protein; ELISA, enzyme-linked immunosorbent assay; Fx, fractional; IgG1, immunoglobulin G1; FCGR, Fc gamma receptor; IgG3, immunoglobulin G3; Ped, pediatric. Biomarker nomenclature: BLI NANP6: BLI NANP6 Dissociation AUC. ELISA: CSP IgG ELISA.

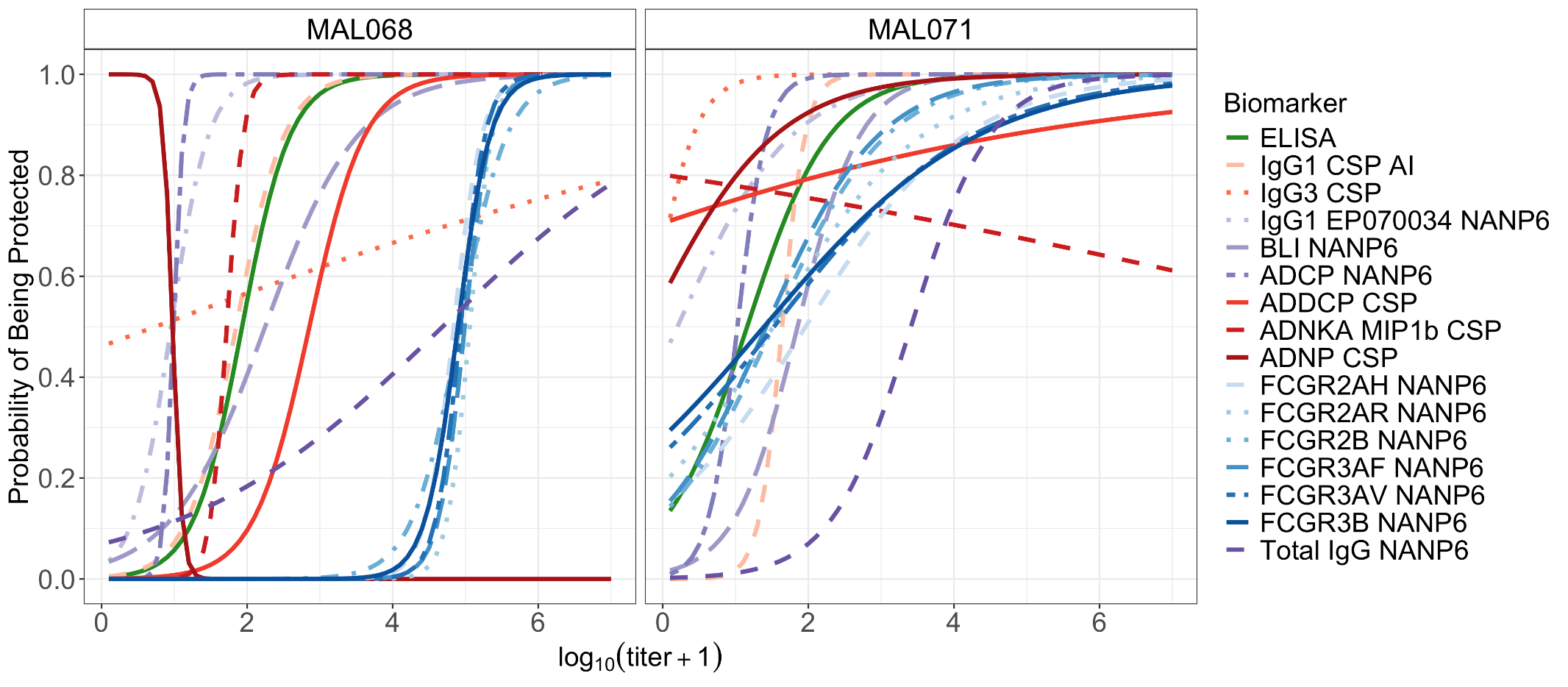

Supplementary Figure 10. Predicted probability of protection by day-of-challenge biomarker level in MAL068 and MAL071. Curves (one curve per biomarker) show probability of protection in MAL068 (left) and MAL071 (right), as predicted by the univariate logistic regression model. ADCP, antibody-dependent cellular phagocytosis; ADDCP, antibody dependent dendritic cell phagocytosis; ADNKA, antibody-dependent natural killer cell activation; ADNP, antibody-dependent neutrophil phagocytosis; AI, avidity index; BLI, bio-layer interferometry; CSP, circumsporozoite protein; ELISA, enzyme-linked immunosorbent assay; IgG1, immunoglobulin G1; FCGR, Fc gamma receptor; IgG3, immunoglobulin G3. Biomarker nomenclature: BLI NANP6: BLI NANP6 Dissociation AUC. ELISA: CSP IgG ELISA.

Supplementary Table 12. Multivariate logistic regression models of protection in MAL092 based on immune signatures containing CSP IgG ELISA.

|  | **ELISA** | | **Biomarker 2** | | **Biomarker 3** | | **BIC*** |
| --- | --- | --- | --- | --- | --- | --- | --- |
|  | Estimate | P-value | Estimate | P-value | Estimate | P-value |  |
| CSP IgG ELISA | 1.84 | **<0.001** |  |  |  |  | 120.79 |
| CSP IgG ELISA + BLI NANP6 Dissociation AUC | 0.09 | 0.93 | 2.5 | **0.051** |  |  | 121.23 |
| CSP IgG ELISA+ IgG1 CSP AI | 1.86 | **<0.001** | 2 | 0.55 |  |  | 124.98 |
| CSP IgG ELISA+ FCGR3AF NANP6 | 0.61 | 0.43 | 1.27 | **0.054** |  |  | 121.64 |
| CSP IgG ELISA+ FCGR3AV NANP6 | 0.38 | 0.65 | 1.69 | **0.04** |  |  | 120.97 |
| CSP IgG ELISA+ ADCP NANP6 | 1.52 | **0.011** | 1.42 | 0.37 |  |  | 124.54 |
| CSP IgG ELISA+ BLI NANP6 Dissociation AUC + FCGR3AF NANP6 | -0.32 | 0.78 | 1.84 | 0.2 | 0.88 | 0.24 | 124.37 |
| CSP IgG ELISA+ BLI NANP6 Dissociation AUC + FCGR3AV NANP6 | -0.52 | 0.68 | 1.78 | 0.22 | 1.24 | 0.18 | 123.87 |
| CSP IgG ELISA+ BLI NANP6 Dissociation AUC + ADCP NANP6 | 0.08 | 0.94 | 2.38 | 0.081 | 0.45 | 0.79 | 125.71 |
| CSP IgG ELISA+ IgG1 CSP AI + FCGR3AF NANP6 | 0.64 | 0.42 | 0.65 | 0.85 | 1.24 | **0.067** | 126.15 |
| CSP IgG ELISA+ IgG1 CSP AI + FCGR3AV NANP6 | 0.41 | 0.64 | 0.42 | 0.91 | 1.66 | **0.05** | 125.5 |
| CSP IgG ELISA+ IgG1 CSP AI + ADCP NANP6 | 1.56 | 0.098 | 1.71 | 0.61 | 1.33 | 0.4 | 128.83 |

*BIC = Bayes Information Criterion; smaller values suggest better models.

Multiple logistic regression was used to model the probability of being protected as a function of bivariate and trivariate immune signatures, all including the CSP IgG ELISA readout, in MAL092. The univariate logistic regression model including CSP IgG ELISA as predictor is also included. The table reports the estimated regression coefficient for each biomarker included in the model (Estimate) and their p-values.

ADCP, antibody-dependent cellular phagocytosis; ADDCP, antibody dependent dendritic cell phagocytosis; ADNKA, antibody-dependent natural killer cell activation; ADNP, antibody-dependent neutrophil phagocytosis; AI, avidity index; BLI, bio-layer interferometry; CSP, circumsporozoite protein; ELISA, enzyme-linked immunosorbent assay; IgG1, immunoglobulin G1; FCGR, Fc gamma receptor; IgG3, immunoglobulin G3.

Supplementary Table 13. Multivariate logistic regression models of protection in MAL102 based on immune signatures containing CSP IgG ELISA.

|  | **ELISA** | | **Biomarker 2** | | **Biomarker 3** | | **BIC*** |
| --- | --- | --- | --- | --- | --- | --- | --- |
|  | Estimate | P-value | Estimate | P-value | Estimate | P-value |  |
| CSP IgG ELISA | 2.28 | **0.019** |  |  |  |  | 50.01 |
| CSP IgG ELISA+ BLI NANP6 Dissociation AUC | -0.78 | 0.8 | 3.35 | 0.31 |  |  | 52.47 |
| CSP IgG ELISA+ IgG1 CSP AI | 2.35 | **0.021** | 12.82 | 0.25 |  |  | 52.15 |
| CSP IgG ELISA+ FCGR3AF NANP6 | 2.86 | **0.024** | -1.71 | 0.30 |  |  | 52.44 |
| CSP IgG ELISA+ FCGR3AV NANP6 | -2.09 | 0.43 | 5.44 | 0.11 |  |  | 49.68 |
| CSP IgG ELISA+ ADCP NANP6 | 1.51 | 0.2 | 2.91 | 0.31 |  |  | 52.55 |
| CSP IgG ELISA+ BLI NANP6 Dissociation AUC + FCGR3AF NANP6 | -0.89 | 0.79 | 4.22 | 0.27 | -1.96 | 0.26 | 54.6 |
| CSP IgG ELISA+ BLI NANP6 Dissociation AUC + FCGR3AV NANP6 | -5.64 | 0.26 | 3.86 | 0.36 | 5.02 | 0.12 | 52.37 |
| CSP IgG ELISA+ BLI NANP6 AUC + ADCP NANP6 | -0.43 | 0.89 | 2.4 | 0.51 | 1.94 | 0.54 | 55.68 |
| CSP IgG ELISA+ IgG1 CSP AI + FCGR3AF NANP6 | 3.37 | **0.026** | 18.29 | 0.16 | -2.49 | 0.19 | 53.78 |
| CSP IgG ELISA+ IgG1 CSP AI + FCGR3AV NANP6 | -1.82 | 0.51 | 9.55 | 0.42 | 5.13 | 0.14 | 52.59 |
| CSP IgG ELISA+ IgG1 CSP AI + ADCP NANP6 | 1.67 | 0.16 | 12.28 | 0.29 | 2.58 | 0.37 | 54.92 |

*BIC = Bayes Information Criterion; smaller values suggest better models

Multiple logistic regression was used to model the probability of being protected as a function of bivariate and trivariate immune signatures, all including the CSP IgG ELISA readout, in MAL102. The univariate logistic regression including ELISA as predictor is also included. The table reports the estimated regression coefficient for each biomarker included in the model (Estimate) and their p-values.

ADCP, antibody-dependent cellular phagocytosis; ADDCP, antibody dependent dendritic cell phagocytosis; ADNKA, antibody-dependent natural killer cell activation; ADNP, antibody-dependent neutrophil phagocytosis; AI, avidity index; BLI, bio-layer interferometry; CSP, circumsporozoite protein; ELISA, enzyme-linked immunosorbent assay; IgG1, immunoglobulin G1; FCGR, Fc gamma receptor; IgG3, immunoglobulin G3.

**B**

**A**

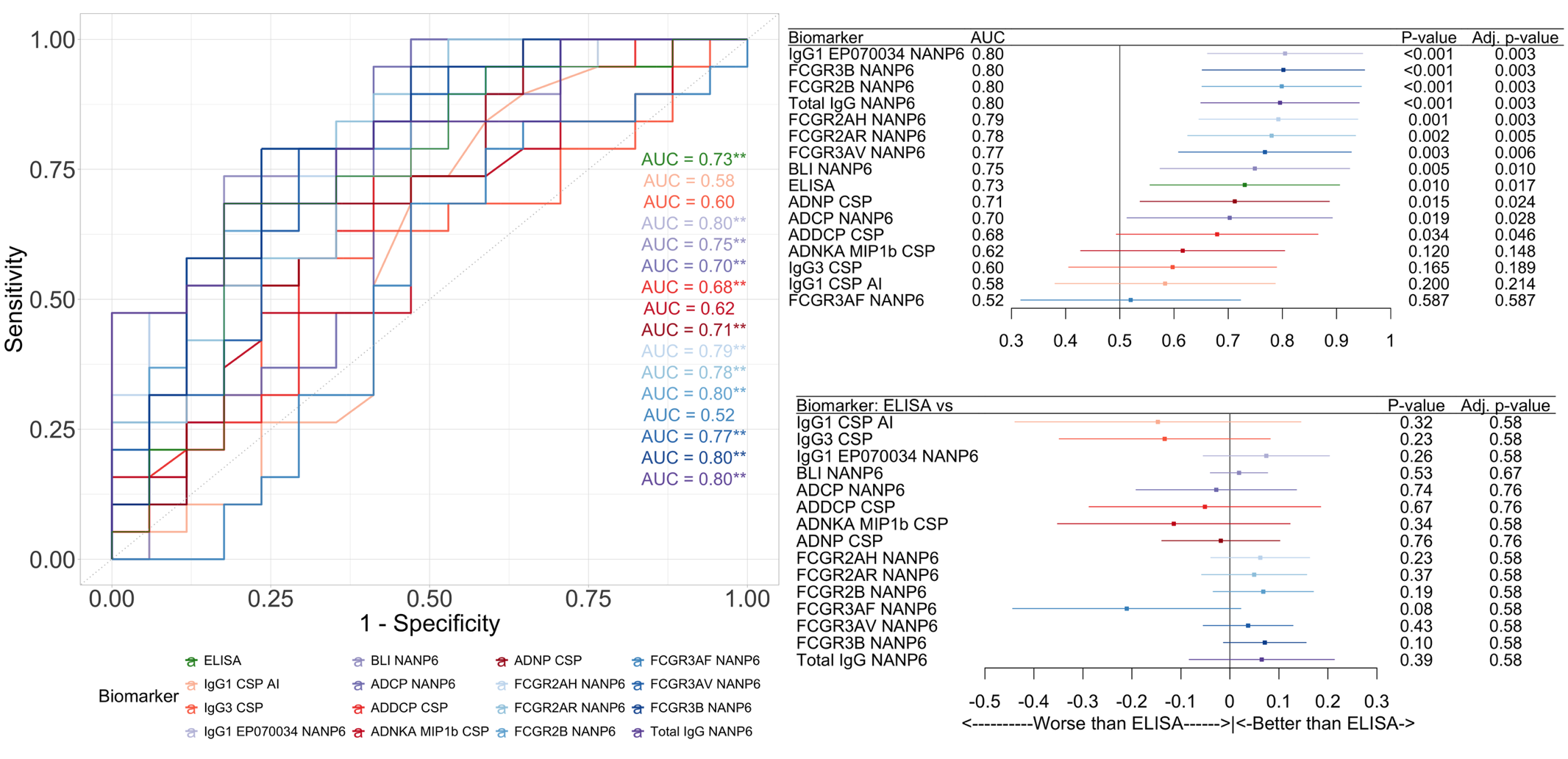

**C**

Supplementary Figure 11. Predictive performance of each biomarker in MAL102. (A) Receiver operating characteristic (ROC) curve of each individual biomarker, and their area under the curve (AUC). (B) AUCs and 95% CIs of each biomarker. A value of 0.5 for the AUC indicates no predictive power, whereas a value in (0.5; 1.0] indicates predictive performances, and perfect predictions with a value of 1.0. The horizontal line extending on either side of each dot represents the 95% confidence interval of the AUC. The p-value assesses the significance of AUC (i.e., AUC = 0.5 or AUC > 0.5). Adjusted p-values that control the false discovery rate for multiple comparisons are provided in the right column. (C) Differences between the AUC from each biomarker and the AUC from CSP IgG ELISA. A value of 0 indicates no difference in AUC, whereas a positive or negative value indicates better or worse performances for the corresponding biomarker relative to CSP IgG ELISA. The horizontal line extending on either side of each dot represents the 95% confidence interval of the difference between the AUC. The p-value assesses the significance of the difference between the AUC. Adjusted p-values that control the false discovery rate for multiple comparisons are provided in the right column of the figure. In (A-C), the color scheme distinguishes the class of antibodies targeted by the readout, with orange representing CSP, blue representing NANP6, and green indicating the CSP IgG ELISA readout. ADCP, antibody-dependent cellular phagocytosis; ADDCP, antibody dependent dendritic cell phagocytosis; ADNKA, antibody-dependent natural killer cell activation; ADNP, antibody-dependent neutrophil phagocytosis; AI, avidity index; BLI, bio-layer interferometry; CSP, circumsporozoite protein; ELISA, enzyme-linked immunosorbent assay; IgG1, immunoglobulin G1; FCGR, Fc gamma receptor; IgG3, immunoglobulin G3. Biomarker nomenclature: BLI NANP6: BLI NANP6 Dissociation AUC. ELISA: CSP IgG ELISA.

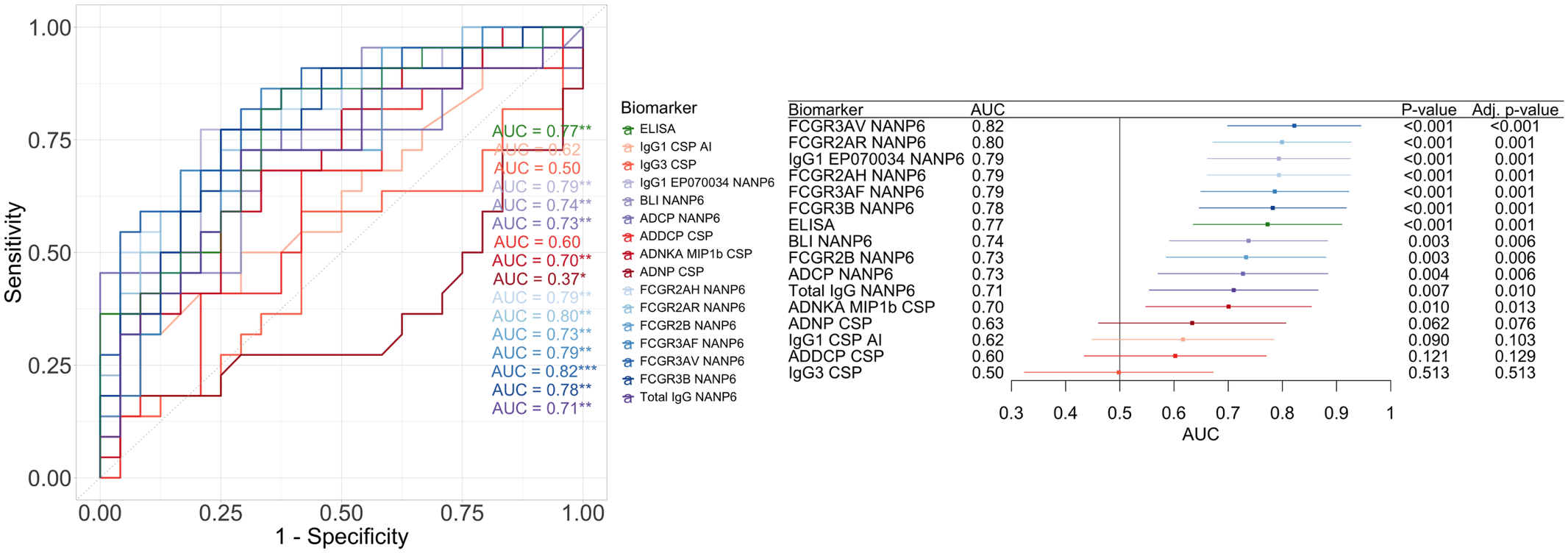

**A**

**B**

Supplementary Figure 12. Predictive performance of each biomarker in MAL068. (A) Receiver operating characteristic (ROC) curve of each individual biomarker, and their area under the curve (AUC). (B) AUCs and 95% CIs of each biomarker. A value of 0.5 for the AUC indicates no predictive power, whereas a value in (0.5; 1.0] indicates predictive performances, and perfect predictions with a value of 1.0. The horizontal line extending on either side of each dot represents the 95% confidence interval of the AUC. The p-value assesses the significance of AUC (i.e., AUC = 0.5 or AUC > 0.5). Adjusted p-values that control the false discovery rate for multiple comparisons are provided in the right column. In (A-B), the color scheme distinguishes the class of antibodies targeted by the readout, with orange representing CSP, blue representing NANP6, and green indicating the CSP IgG ELISA readout. ADCP, antibody-dependent cellular phagocytosis; ADDCP, antibody dependent dendritic cell phagocytosis; ADNKA, antibody-dependent natural killer cell activation; ADNP, antibody-dependent neutrophil phagocytosis; AI, avidity index; BLI, bio-layer interferometry; CSP, circumsporozoite protein; ELISA, enzyme-linked immunosorbent assay; IgG1, immunoglobulin G1; FCGR, Fc gamma receptor; IgG3, immunoglobulin G3. Biomarker nomenclature: BLI NANP6: BLI NANP6 Dissociation AUC. ELISA: CSP IgG ELISA.

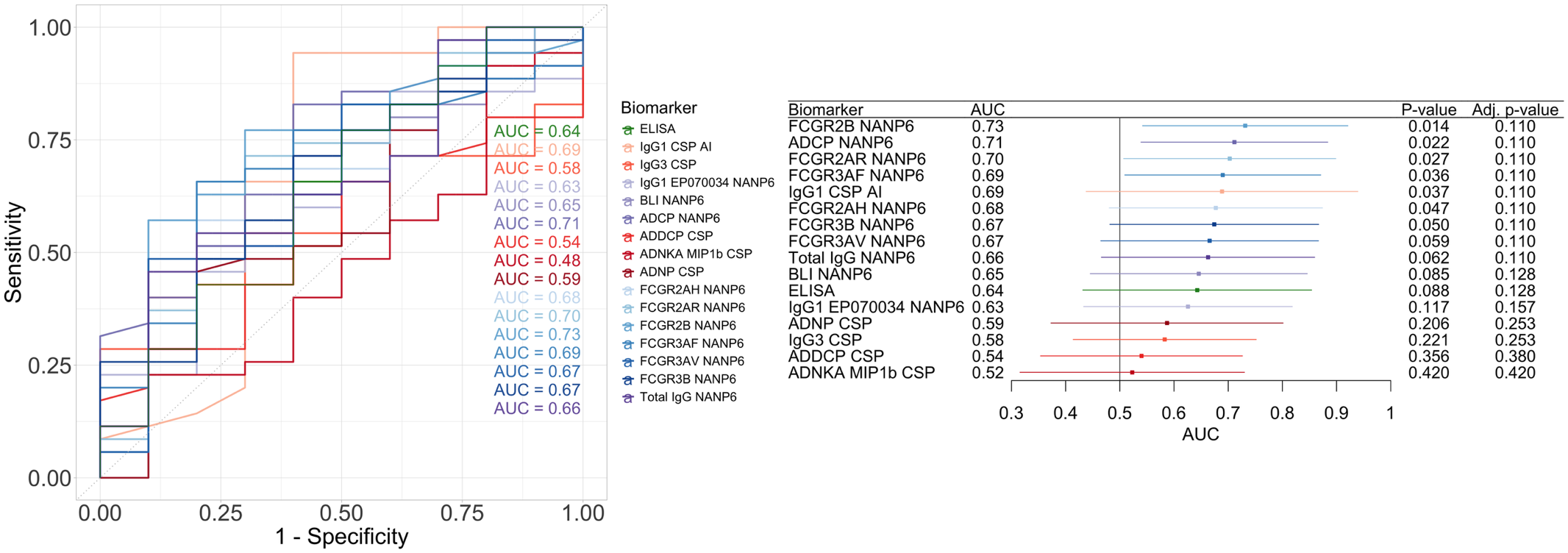

**B**

**A**

Supplementary Figure 13. Predictive performance of each biomarker in MAL071. (A) Receiver operating characteristic (ROC) curve of each individual biomarker, and their area under the curve (AUC). (B) AUCs and 95% CIs of each biomarker. A value of 0.5 for the AUC indicates no predictive power, whereas a value in (0.5; 1.0] indicates predictive performances, and perfect predictions with a value of 1.0. The horizontal line extending on either side of each dot represents the 95% confidence interval of the AUC. The p-value assesses the significance of AUC (i.e., AUC = 0.5 or AUC > 0.5). Adjusted p-values that control the false discovery rate for multiple comparisons are provided in the right column. In (A-B), the color scheme distinguishes the class of antibodies targeted by the readout, with orange representing CSP, blue representing NANP6, and green indicating the CSP IgG ELISA readout. ADCP, antibody-dependent cellular phagocytosis; ADDCP, antibody dependent dendritic cell phagocytosis; ADNKA, antibody-dependent natural killer cell activation; ADNP, antibody-dependent neutrophil phagocytosis; AI, avidity index; BLI, bio-layer interferometry; CSP, circumsporozoite protein; ELISA, enzyme-linked immunosorbent assay; IgG1, immunoglobulin G1; FCGR, Fc gamma receptor; IgG3, immunoglobulin G3. Biomarker nomenclature: BLI NANP6: BLI NANP6 Dissociation AUC. ELISA: CSP IgG ELISA.

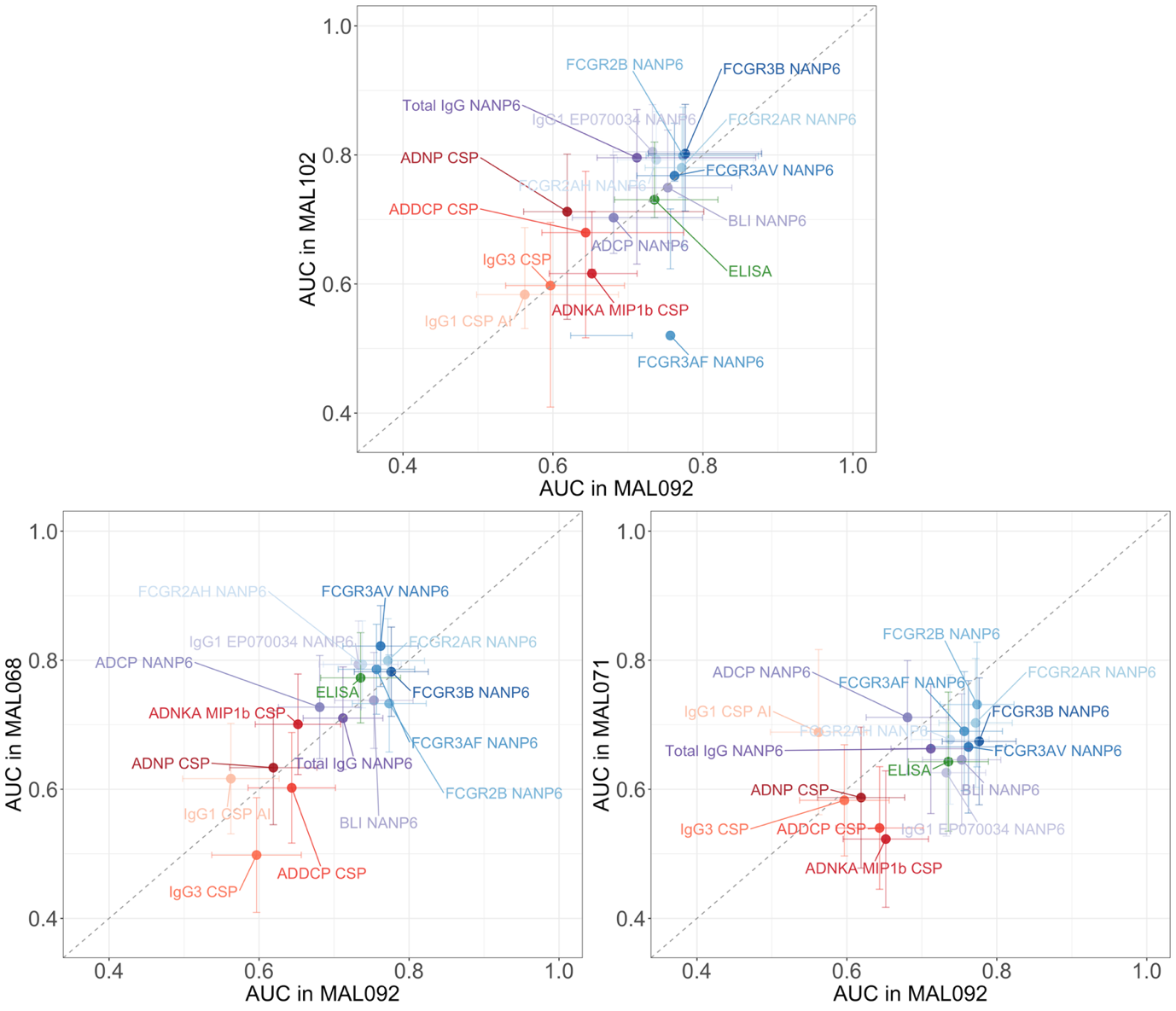
Supplementary Figure 14. Comparison of the predictive power of individual biomarkers between studies. (A) MAL092 versus MAL102, (B) MAL092 versus MAL102, (C) MAL068 versus MAL071. Each dot signifies the AUC attained by the corresponding readout in the two studies. The horizontal and vertical bars extending from the dots denote the 95% confidence interval of the AUC in the respective study. The color scheme distinguishes the class of antibodies targeted by the readout, with orange representing CSP, blue representing NANP6, and green indicating the CSP IgG ELISA readout. ADCP, antibody-dependent cellular phagocytosis; ADDCP, antibody dependent dendritic cell phagocytosis; ADNKA, antibody-dependent natural killer cell activation; ADNP, antibody-dependent neutrophil phagocytosis; AI, avidity index; BLI, bio-layer interferometry; CSP, circumsporozoite protein; ELISA, enzyme-linked immunosorbent assay; IgG1, immunoglobulin G1; FCGR, Fc gamma receptor; IgG3, immunoglobulin G3. Biomarker nomenclature: BLI NANP6: BLI NANP6 Dissociation AUC. ELISA: CSP IgG ELISA.

**C**

**B**

**A**

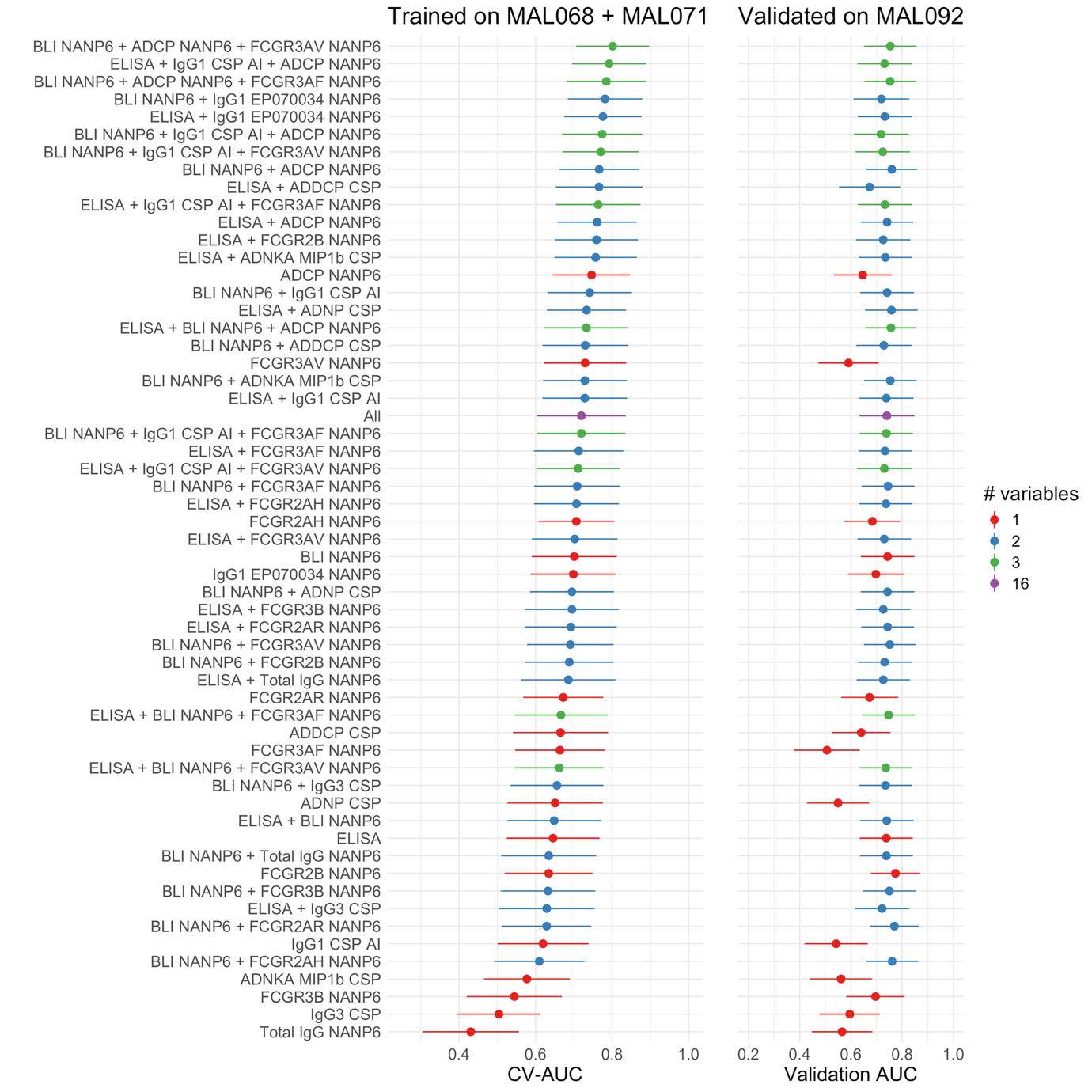

Supplementary Figure 15. Predictive performances of multivariate immune signatures in MAL092. The predictive signatures were trained on data from the MAL068 and MAL071 trials and independently validated in MAL092. The forest plot of the left panel shows the cross-validated AUC from the predictive models trained on MAL068 and MAL071, along with their 95% confidence intervals, by immune signature. The forest plot of the right panel shows the corresponding AUC and their 95% confidence intervals, when the immune signatures are applied to predict protection in MAL092. For simplicity and ease of visualization, the subset of bivariate models included in the figure was restricted to those that included CSP IgG ELISA and/or BLI NANP6 as predictors, whereas the subset of trivariate models was mostly restricted to those that were pre-specified in the Statistical Analysis Plan. ADCP, antibody-dependent cellular phagocytosis; ADDCP, antibody dependent dendritic cell phagocytosis; ADNKA, antibody-dependent natural killer cell activation; ADNP, antibody-dependent neutrophil phagocytosis; AI, avidity index; BLI, bio-layer interferometry; CSP, circumsporozoite protein; ELISA, enzyme-linked immunosorbent assay; IgG1, immunoglobulin G1; FCGR, Fc gamma receptor; IgG3, immunoglobulin G3. Biomarker nomenclature: BLI NANP6: BLI NANP6 Dissociation AUC. ELISA: CSP IgG ELISA.

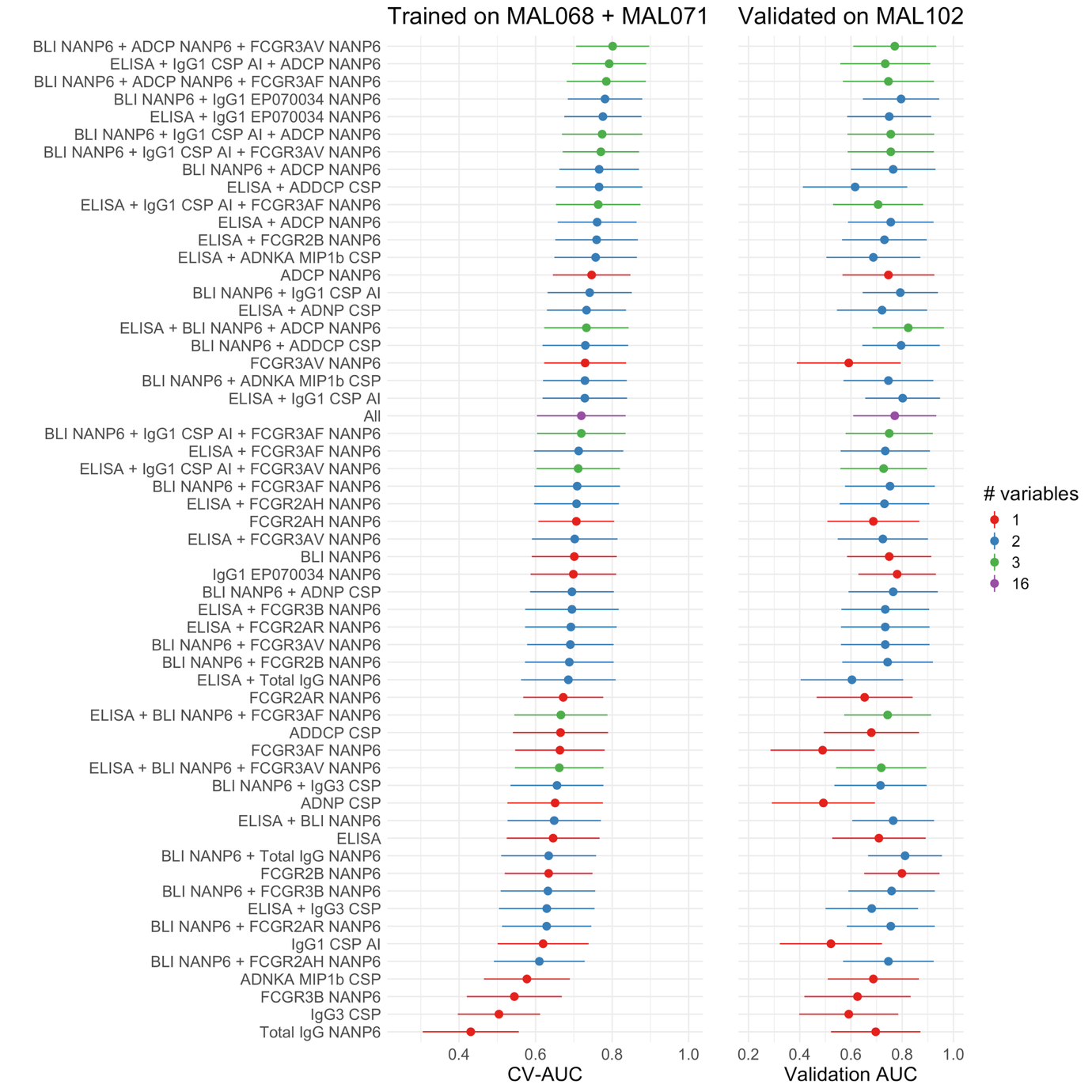

Supplementary Figure 16. Predictive performances of multivariate immune signatures in MAL102. The predictive signatures were trained on data from the MAL068 and MAL071 trials and independently validated in MAL102. The forest plot of the left panel shows the cross-validated AUC from the predictive models trained on MAL068 and MAL071, along with their 95% confidence intervals, by immune signature. The forest plot of the right panel shows the corresponding AUC and their 95% confidence intervals, when the immune signatures are applied to predict protection in MAL102. For simplicity and ease of visualization, the subset of bivariate models included in the figure was restricted to those that included CSP IgG ELISA and/or BLI NANP6 as predictors, whereas the subset of trivariate models was mostly restricted to those that were pre-specified in the Statistical Analysis Plan. ADCP, antibody-dependent cellular phagocytosis; ADDCP, antibody dependent dendritic cell phagocytosis; ADNKA, antibody-dependent natural killer cell activation; ADNP, antibody-dependent neutrophil phagocytosis; AI, avidity index; BLI, bio-layer interferometry; CSP, circumsporozoite protein; ELISA, enzyme-linked immunosorbent assay; IgG1, immunoglobulin G1; FCGR, Fc gamma receptor; IgG3, immunoglobulin G3. Biomarker nomenclature: BLI NANP6: BLI NANP6 Dissociation AUC. ELISA: CSP IgG ELISA.
